# Garetosmab, an inhibitor of activin A, reduces heterotopic ossification and flare-ups in adults with fibrodysplasia ossificans progressiva: a randomized, double-blind, placebo-controlled phase 2 trial

**DOI:** 10.1101/2023.01.11.23284254

**Authors:** Maja Di Rocco, Eduardo Forleo-Neto, Robert Pignolo, Richard Keen, Philippe Orcel, Thomas Funck-Brentano, Christian Roux, Sami Kolta, Annalisa Madeo, Judith S Bubbear, Jacek Tabarkiewicz, Małgorzata Szczepanek, Javier Bachiller-Corral, Angela M Cheung, Kathryn M Dahir, Esmée Botman, Pieter G Raijmakers, Mona Al Mukaddam, Lianne Tile, Cynthia Portal-Celhay, Neena Sarkar, Peijie Hou, Bret Musser, Anita Boyapati, Kusha Mohammadi, Scott Mellis, Andrew J. Rankin, Aris N. Economides, Dinko Gonzalez Trotter, Gary Herman, Sarah J. O’Meara, Richard DelGizzi, David M. Weinreich, George D. Yancopolous, E. Marelise W. Eekhoff, Frederick S. Kaplan, the LUMINA-1 Investigators

## Abstract

**Background:** Fibrodysplasia ossificans progressiva (FOP), an ultra-rare disorder caused by mutations in the gene encoding activin A receptor type 1 (*ACVR1)*, is characterized by painful flare-ups and cumulative heterotopic ossification (HO). Garetosmab, a fully-human monoclonal antibody blocking activin A, prevents HO in FOP mice.

**Methods:** LUMINA-1 (NCT03188666) was a phase 2, multi-center, randomized, double-blind, placebo-controlled study evaluating the safety, tolerability, and effects on HO of intravenous (IV) garetosmab 10 mg/kg every 4 weeks (Q4W). Adult patients with FOP were randomized to garetosmab or placebo for 28 weeks (Period_1), followed by an open-label period (Period_2). After Period_2, patients were allowed to stay on garetosmab in an open-label extension. For Period_1, primary endpoints were HO total lesion activity (HO-TLA) by ^18^F-sodium fluoride positron emission tomography (^18^F-NaF PET) and HO total lesion volume by computed tomography (CT). The Period_2 primary endpoint compared the number of new lesions in Period_2 versus Period_1. The safety primary endpoint was incidence and severity of TEAEs through the end of the Period 1 at week 28.

**Findings:** Patients (n=44) were randomized to garetosmab (n=20) or placebo (n=24). In Period_1, there was a trend for garetosmab to decrease HO-TLA versus placebo (24.6%; *P*=0.07), primarily driven by near complete prevention of new lesions (97% decrease by ^18^F-NaF PET, post-hoc *P*=0.009; 90% relative reduction by CT, post-hoc *P*=0.017); flare-ups were significantly reduced (*P*=0.0005). For placebo patients transitioning to garetosmab in Period_2, no patients developed new HO lesions (0% in Period_2 versus 40.9% in Period_1; *P*=0.0027) by CT. All 44 patients met primary safety endpoint of at least one TEAE during Period 1. Garetosmab was associated with more adverse events than placebo: mild recurrent epistaxis, madarosis, and skin/soft tissue infections. Overall, the AEs were predominantly mild in severity, with no effect on patients’ ability to receive garetosmab. Five deaths (5/44; 11.4%) occurred either in Period_2 or the open-label extension. The deaths were associated with baseline disease severity in some, preexisting comorbidities in others and occurred following 8-16 doses (median: 15) of garetosmab in the open label/follow-up periods.

**Interpretation:** Garetosmab reduced flare-ups and prevented new HO lesions in FOP patients. Although side effects were mild to moderate, there were a relatively high number of deaths for a small study; the deaths were not related to epistaxis and considered unlikely to be related to garetosmab.

**Funding:** Regeneron Pharmaceuticals, Inc.

## Introduction

Fibrodysplasia ossificans progressiva (FOP; MIM#135100) is an ultra-rare, autosomal dominant disorder^1^ with an estimated prevalence of 0.36–1.36 per million.^2-5^ The most common and debilitating manifestation of FOP is the heterotopic ossification (HO) of connective tissues, such as tendons, ligaments, and skeletal muscles excepting the diaphragm, extraocular muscles and the tongue.^6^ HO typically presents during the first decade of life and may be accompanied by painful, soft tissue inflammatory swellings, known as flare-ups, however, this feature of the disease is variable.^7^ A diagnostic hallmark of FOP at birth is bilateral malformation of the great toes, which is often missed and can lead to a delay in diagnosis.^8^

HO in FOP is seemingly episodic, and may be precipitated by trauma.^9^ The resulting HO lesions accumulate, leading to joint immobility, skeletal deformity, severe pain, disability, and early mortality.^6,10,11^ Patients with FOP experience significant disease burden including restriction of activity as well as hearing loss which worsens social isolation leading to decreased quality of life. Surgical removal of bone is ineffective as it causes new HO and is associated with high anesthetic challenges.^12^ The degree of immobility can be captured by the cumulative analog joint involvement scale (CAJIS) score, measuring mobility across 15 anatomic locations, and ranging from 0 to 30 (higher scores indicating more severe limitations in mobility and other function).^13^ On the basis of FOP features related to joint dysfunction (flare-up activity, body regions affected, thoracic insufficiency, and other complications) as well as its consequences (assistance in activities of daily living, ambulation, and CAJIS score), 5 clinical stages of FOP have been defined: early, moderate, severe, profound, and end of life.^14^ Estimated median age of survival for patients with FOP is 56 years.^15^ Mortality is correlated with disease severity and CAJIS score and is primarily due to cardiorespiratory failure from thoracic insufficiency syndrome, pneumonia, and complications of falls.^15^ With no globally-approved disease-modifying therapies, clinical management is mostly limited to symptomatic flare-up treatment with corticosteroids or non-steroidal anti-inflammatory agents, evidence of which is anecdotal.^7^

FOP is caused by heterozygous missense mutations in *ACVR1*, the gene encoding activin A receptor type 1, a bone morphogenetic protein (BMP) type I receptor.^16^ A significant breakthrough in understanding the molecular mechanism of HO in FOP was the discovery that FOP-causing variants of ACVR1 recognize activin A as an agonist (**Figure S1**).^17-20^ In contrast, activin A normally inhibits BMP signalling via wild-type ACVR1.^17,21^ Garetosmab, a fully-human monoclonal antibody (generated using *VelocImmune™ technology)*,^1,2^ binds activin A and blocks its ability to activate FOP-mutant ACVR1. Inhibition of activin A with garetosmab blocked new HO lesions and resulted in stasis or partial reduction in the volume of pre-existing lesions in a genetically humanized murine FOP model in which FOP-mutant ACVR1 was introduced.^17,18^ As such, it was hypothesized that garetosmab may beneficially impact FOP disease progression in humans. LUMINA-1 was designed to assess the safety and efficacy of garetosmab as a potential disease-modifying treatment for FOP, and consequently also validate the role of activin A as a key driver of HO in FOP.

## Methods

### Study design

LUMINA-1 (NCT03188666) was a phase 2, randomized, double-blind, placebo-controlled study evaluating the safety, tolerability, and effects on HO of intravenous (IV) garetosmab 10 mg/kg every 4 weeks (Q4W). The study was conducted at 11 sites in 8 countries across North America and Europe, between February 26, 2018 and September 16, 2021. ^18^F-NaF PET and whole-body, low-dose X-ray computed tomography (CT)^22^ were used to measure and track HO. The study consisted of a 28-day screening/baseline period followed by a 28-week randomized, double-blind, placebo-controlled period (Period_1), a 28-week open-label treatment period (Period_2), and a subsequent open-label extension from week 56 to end of study (**Figure S2**). The primary analysis was conducted when all patients completed Period_1. Further analyses were conducted at the end of Period_2. Amendments to the trial protocol are detailed in **Table S1**. Patient safety and welfare were monitored by an Independent Data Monitoring Committee. This study was conducted in accordance with the 2013 Declaration of Helsinki and the International Council for Harmonization guidelines for GCP. All patients provided written, informed consent. Detailed information and a full list of the Institutional Review Boards are in the **Supplementary Methods**.

### Patients

Adult patients (aged 18–60 years) with a clinical diagnosis of FOP (based on congenital great toe malformation, episodic soft tissue swelling, and/or progressive HO) were included. Clinical diagnosis was confirmed with documentation of any FOP-causing *ACVR1* variant and disease activity (defined as pain, swelling, stiffness, and other signs/symptoms associated with FOP flare-ups; or worsening of joint function, or radiographic progression of HO) within 1 year of screening. Key inclusion/exclusion criteria are in **Table S2** and **Supplementary Methods**.

### Randomization and masking

Enrolled patients were randomized (1:1) to receive garetosmab 10 mg/kg Q4W as previously assessed,^23^ or placebo in Period_1, according to a central randomization scheme. Block randomization was done using an interactive response technology (IRT) provided to the designated study pharmacist or qualified designee. Randomization was stratified by presence/absence of baseline active HO lesions, gender, and mutation type. All PET/CT scans were reviewed by 2 independent readers and an adjudicator; all 3 were blinded to treatment assignment.

### Procedures

Patients were assigned to receive garetosmab 10 mg/kg or placebo Q4W for 28 weeks (Period_1). After week 28, all patients continued or were switched to receive garetosmab 10 mg/kg Q4W (Period_2). At the conclusion of Period_2, patients were given the option to continue on garetosmab in an open-label extension. Whole-body PET/CT scans were acquired to identify pre-existing target lesions at baseline, identify new HO lesions, and measure volume changes in pre-existing target and new HO lesions. Baseline imaging with PET/CT was performed at most 7 days prior to study drug administration and at Weeks 8, 28, 56, and 76 (**Figure S2**). Detailed information on imaging acquisition and read procedures are in the **Supplementary Methods**.

### Outcomes

#### Period_1

The pre-specified primary endpoint for efficacy was the effect of garetosmab versus placebo on time-weighted average (TWA) of the percentage change from baseline in total lesion activity (TLA) by PET; TLA is considered proportional to the deposition rate of bone mineral into actively forming HO lesions (definition in supplementary methods section). The next endpoint in the hierarchy was to assess percentage change of the total volume of HO lesions by CT in Period_1 relative to baseline. Other key secondary and exploratory endpoints are listed in **Table S3**. Exploratory endpoints in Period_1 included the percentage of patients with flare-ups as assessed by patient diary and investigator report.

#### Period_2

Based on the outcomes of Period_1 (see Results), the primary endpoint for efficacy for Period_2 was prospectively changed to the number of new lesions in patients transitioning from placebo to garetosmab as assessed by CT.

#### Safety

The primary safety endpoint for Period_1 and a primary endpoint for Period_2 was the incidence and severity of adverse events, which included those not present at baseline or an exacerbation of a pre-existing condition.

### Statistical analyses

Testing of primary and key secondary efficacy outcomes followed a pre-specified hierarchical testing procedure to address multiplicity at an overall 2-sided alpha=0.05 significance for Period_1. Other secondary outcomes were tested at the nominal 2-sided alpha=0.05 significance without multiplicity adjustment. In Period_2, a 2-sided 10% significant level was applied for the primary and key secondary endpoints. For the

Period_1 primary analysis, the TWA percent change from baseline in TLA ^18^F-NaF PET over 28 weeks was analyzed in AHO (baseline active HO analysis set) and AHOC (baseline active HO classic ACVR1[R206H] mutation analysis set) populations using an analysis of covariance model. The difference in least squares mean change from baseline, 95% confidence interval, and *P*-value were provided from the model to compare garetosmab against placebo group. For Period_2 primary analysis, the Wilcoxon signed-rank test was used to compare the number of new HO lesions as assessed by CT at week 56 (relative to week 28 scan) with the number of new HO lesions at week 28 (relative to baseline scan) in patients who crossed over from placebo to garetosmab. The estimate and 95% confidence interval of the rate of new HO lesions at week 56 and that of the rate ratio (comparing Period_2 versus Period_1) were based on a negative binomial model with repeated measures and using a generalized estimating equation. Safety outcomes were analyzed using descriptive statistics. Further details are provided in the **Supplementary Methods** and amendments to the statistical methodology are summarized in **Table S1**.

## Results

LUMINA-1 enrolled 44 patients with FOP, all of whom had active HO (AHO) at baseline (see full definition in the **Supplementary Methods**). Overall, 42/44 (95.5%) of patients had the “classic” FOP-causing variant of *ACVR1* (c.617G>A, p.R206H) (AHOC population). Patients were randomized to garetosmab 10 mg/kg IV Q4W (n=20) or placebo (n=24) (**Figure S2; Figure S3**). Baseline demographics and disease characteristics were similar between groups (**Table S4**). The mean (SD) age was 27.6 (8.5) years. The mean CAJIS score at baseline was 15.7 (SD: 6.6; range, 6–30; Median [Quartile1, Quartile3]: 15.0 [11, 19]). Data analyses presented are for the AHO population and were similar to the AHOC population (**Table S5A**). The study was initiated in February 2018; primary data cut-off was September 17, 2019 (week 28); additional data cut-off dates for efficacy analyses were August 11, 2020 (week 56), and October 30, 2020 (week 76); safety was reported until last patient last visit on September 16, 2021.

### Period_1 Results

Forty-three (98%) patients completed Period_1. During Period_1, garetosmab reduced TLA (new and existing lesions) from baseline versus placebo by 24.6% (95% confidence interval [CI]: −51.8%–2.5%; *P*=0.07; **Figure 1A**; **Supplementary Section: Results**). Although this did not meet the pre-specified statistical threshold of *P*≤0.05, patients treated with garetosmab had a ∼97% relative reduction in new lesion activity, a ∼90% relative reduction in new HO lesion volume compared with the placebo arm (**Figure 1B**,**C; Table S6**). Additionally, a reduction in both the number of new HO lesions per patient as assessed by PET (mean of 0.15 new lesions/patient for garetosmab versus 1.19 for placebo; post-hoc analysis, nominal *P*=0.006) and CT (0.15 versus 1.13, respectively; post-hoc analysis, nominal *P*=0.009; **Table S6**) were observed. The total number of new HO lesions in Period_1 for placebo was 27 by CT and 29 by PET compared with 3 by CT and PET for garetosmab (**Figure 1B**). The percentage of patients who developed new lesions in Period_1 was lower with garetosmab (15% by CT; 15% by PET) versus placebo (45.8% by CT and PET; nominal *P=*0.05; **Figure S4**).

**Figure 1.**
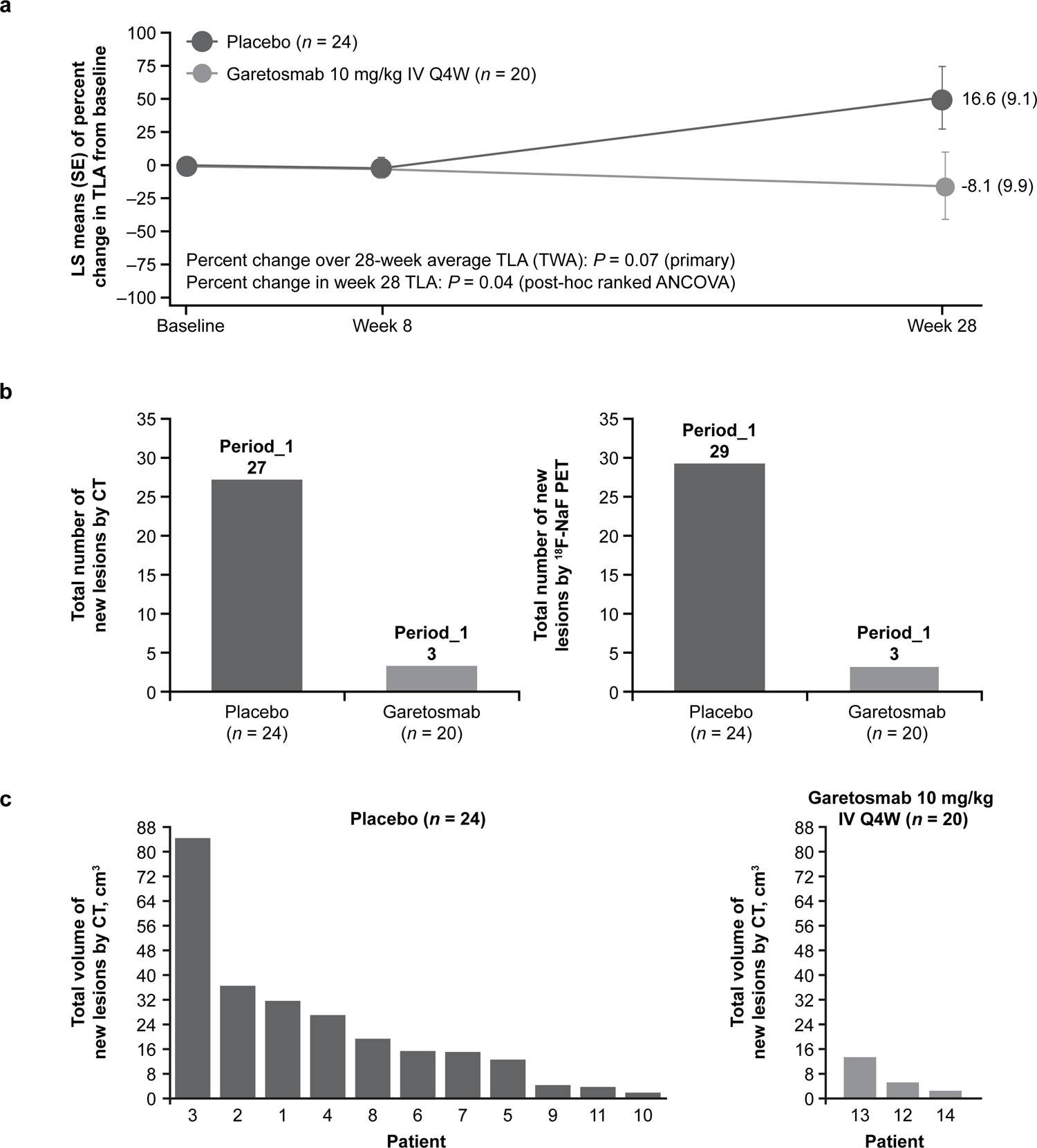
Effect of garetosmab on the (A) change from baseline in TLA compared with placebo, (B) total number of new HO lesions assessed by quantitative imaging, and (C) total volume of new HO lesions by CT per patient in Period_1. **(A)** Time-weighted average of the percent change from baseline in total lesion activity as assessed by ^18^F-NaF PET over 28 weeks (active HO analysis set). **(B)** Total number of new lesions in all patients per group (combined) by CT and ^18^F-NaF PET during Period_1 relative to baseline (active HO analysis set). **(C)** Total volume of new lesions per patient as assessed by CT in Period_1 (active HO analysis set). ^18^F-NaF PET=fluorine-18-labelled sodium fluoride positron emission tomography; HO=heterotopic ossification; IV=intravenous; LS=least squares; Q4W=every 4 weeks; SE=standard error; TLA=total lesion activity; TWA=time-weighted average.

Given that garetosmab appears to block the formation of new HO in patients with FOP, its effects on normal bone were also examined as reference using PET. Assessment of percent change from baseline of the mean [standard error] SUVmean of selected normotopic bones (as defined in the protocol), showed no differences between treatments at week 8 (garetosmab: 42.0% [9.9]; placebo: 20.3% [9.2]; nominal *P*=0.12) or week 28 (garetosmab: 24.6% [8.4]; placebo: 28.9% [7.7]; nominal *P*=0.71; data not shown), indicating that garetosmab does not affect bone turnover.

In addition to the reduction in the number of new HO lesions, an effect of garetosmab was also observed on reported flare-ups. A marked reduction was observed during Period_1 in patients reporting flare-ups on garetosmab versus placebo based on patient diary entries (35.0% versus 70.8%, respectively; relative risk [RR]=0.49; nominal *P*=0.03) and investigator adverse event reports (10.0% versus 41.7%, respectively; RR=0.24; post-hoc analysis, nominal *P*=0.039; **Table 1A; Figure S5A, B**).

**Table 1.** Percent of patients with flare-ups by patient diary and by investigator adverse event (AE) verbatim term in (A) Period_1 and (B) Period_2 (mITT)

| Incidence of flare-up events |  |  |  |
| --- | --- | --- | --- |
| A<br>Double-blind Period_1 (P1)*<br>Inter-group comparison – n (%) |  |  |  |
| Patient diary | Placebo (n=24) | Garetosmab (n=20) | P value |
|  | 17 (70.8) | 7 (35.0) | 0.03 |
| Investigator AE verbatim term | 10 (41.7) | 2 (10.0) | 0.04 |
| B<br>Period_2 (P2) compared to Period_1 (P1)**<br>Intra-group comparison – n (%) |  |  |  |
|  | Placebo (P1) vs Garetosmab (P2) (n=22)# |  | P value |
| Patient diary | 15 (68.2) | 3 (13.6) | 0.0005 |
| Investigator AE verbatim term | 10 (45.5) | 3 (13.6) | 0.019 |

\*Active HO analysis set; \*\* mITT analysis set; #No comparison performed since the intent is to evaluate persistency of garetosmab effect. mITT=modified intent-to-treat. \*Of the 24 patients on placebo in Period\_1, 22 went on to garetosmab in Period\_2. Shown here are only those 22 patients who transitioned from placebo to garetosmab.

Since garetosmab’s most pronounced effect on HO during Period_1 was the suppression of new HO lesions formation, the study hypothesis for Period_2 was redefined to be that garetosmab prevents formation of new HO lesions.

### Period_2 Results

Of the 43 patients who completed Period_1, all entered Period_2 and 42 patients completed Period_2. Patients randomized in Period_1 to placebo crossed-over to receive garetosmab. In Period_2, the total number of new lesions measured by CT for patients who crossed over from placebo to garetosmab was reduced 100% relative to Period_1, (0 versus 22, respectively; *P*=0.0039) and 95% by PET (1 versus 23, respectively; *P*=0.0039; **Figure 2A, B; Table S5B**). Mean new lesion volume and activity were significantly lower for Period_2 versus Period_1 (volume: 0.05 cm^3^ versus 9.29 cm^3^, respectively, *P*=0.0039; activity: 13.20 versus 204.45, respectively; *P*=0.0273; **Figure 2C, D**). This reduction was also reflected in the percentage of patients who developed new lesions, which was significantly lower during Period_2 compared with Period_1 by CT (0% versus 40.9%, respectively; *P*=0.0027) and PET (4.5% versus 40.9%, respectively; *P*=0.0047; **Figure 2E, F)**.

**Figure 2.**
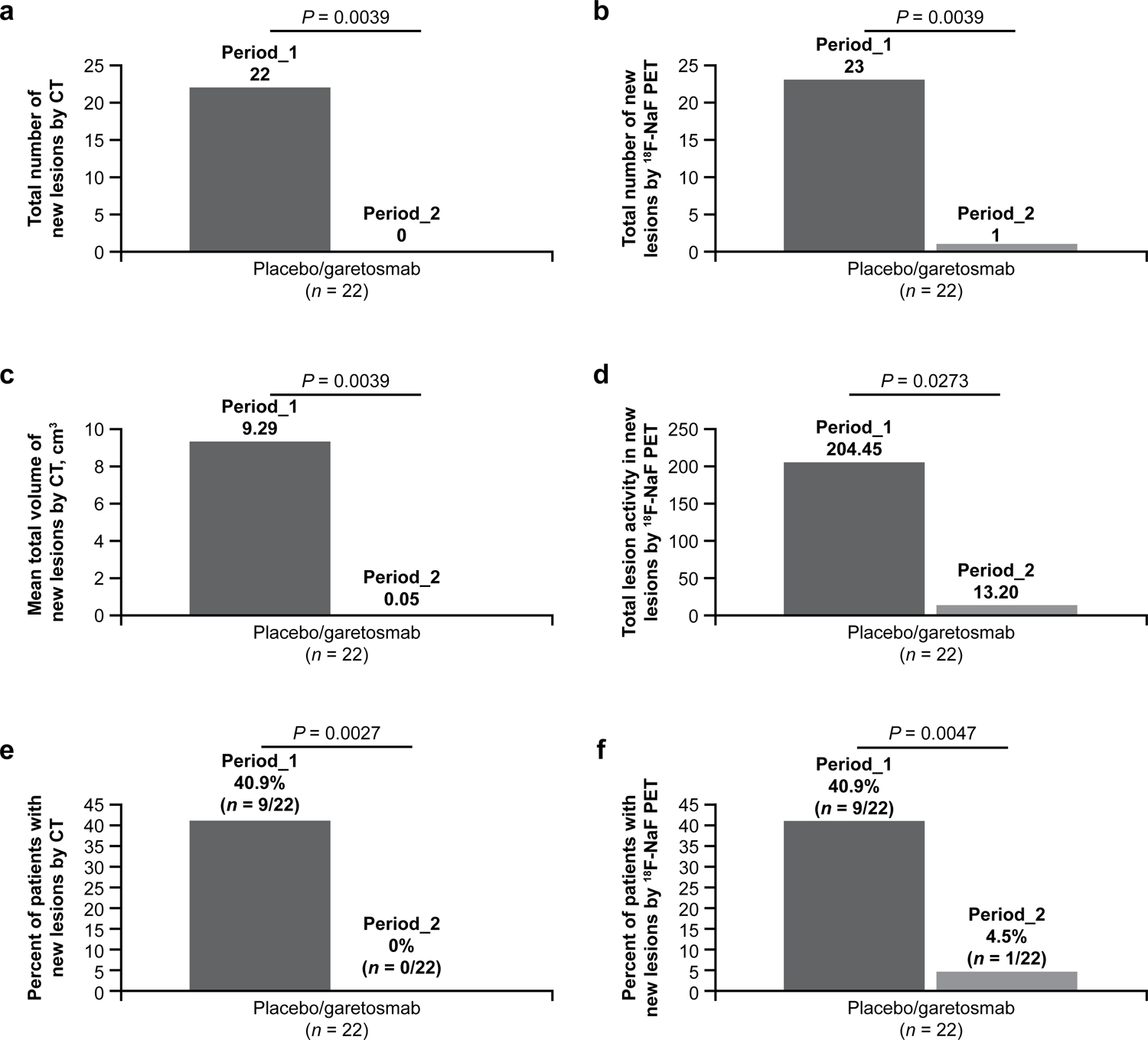
Effect of garetosmab in Period_2 relative to Period_1 on the (A, B) total number of new HO lesions, (C) mean total volume of new HO lesions (D) TLA in new HO lesions, and (E, F) percent of patients with new lesions assessed by quantitative imaging in patients originally randomized to placebo. **(A)** Total number of new lesions by CT during Period_2 relative to Period_1 (mITT analysis set). **(B)** Total number of new lesions by ^18^F-NaF PET during Period_2 relative to Period_1 (mITT analysis set). **(C)** Mean total volume of new lesions as assessed by CT in Period_2 relative to Period_1 (mITT analysis set). **(D)** TLA of new lesions by 18F-NaF PET in Period_2 relative to Period_1 (mITT analysis set). **(E)** Percent of patients with new lesions by CT during Period_2 relative to Period_1 (mITT analysis set). **(F)** Percent of patients with new lesions by ^18^F-NaF PET during Period_2 relative to Period_1 (mITT analysis set).^18^F-NaF PET=fluorine-18-labelled sodium fluoride positron emission tomography; CT=computed tomography; HO=heterotopic ossification; mITT=modified intent-to-treat; TLA=total lesion activity. A new flare-up was defined as a flare-up event starting in the corresponding period. Investigator-defined flare-ups were defined as TEAEs with the verbatim term containing “flare”. HO=heterotopic ossification; mITT=modified intent-to-treat; TEAE=treatment-emergent adverse event.

For patients originally randomized to garetosmab and who remained on garetosmab in Period_2, garetosmab’s efficacy in preventing new HO lesions was maintained (0 new lesions by CT and PET in Period_2 versus 2 by CT and 1 by PET in Period_1; **Figure S6**. Additionally, the percentage of patients with new lesions was 0% for Period_2 by PET and CT (**Figure S6**) for patients remaining on garetosmab.

### Open-Label Extension Results

During the subsequent open-label extension, 0 new lesions at week 76 were observed in patients crossing to garetosmab after week 28 (n=17) while one new lesion was observed by both PET and CT at 76 weeks among patients continuing garetosmab since baseline (n=15). Overall, treatment with garetosmab resulted in a sustained and pronounced effect in preventing new HO lesions from forming up to 76 weeks (the time of last imaging scan assessments in the study). Thirty-four (77%) patients continued in the study, after the open-label extension.

### Flare-Ups and CAJIS

For patients switching to garetosmab after week 28, the proportion with new flare-ups was significantly lower in Period_2 compared to Period_1 as reported by both patients (13.6% versus 68.2%, respectively; *P*=0.0005) and investigators (13.6% versus 45.5%, respectively; *P*=0.0196; **Table 1B; Figure S5C, D**). The total number of new flare-ups by patient diary was 11 in Period_2 compared with 31 in Period_1 and by investigator report was 4 and 22, respectively (**Figure S5E, F**). The relationship between baseline age and CAJIS score is depicted in **Figure 3A**.

**Figure 3.**
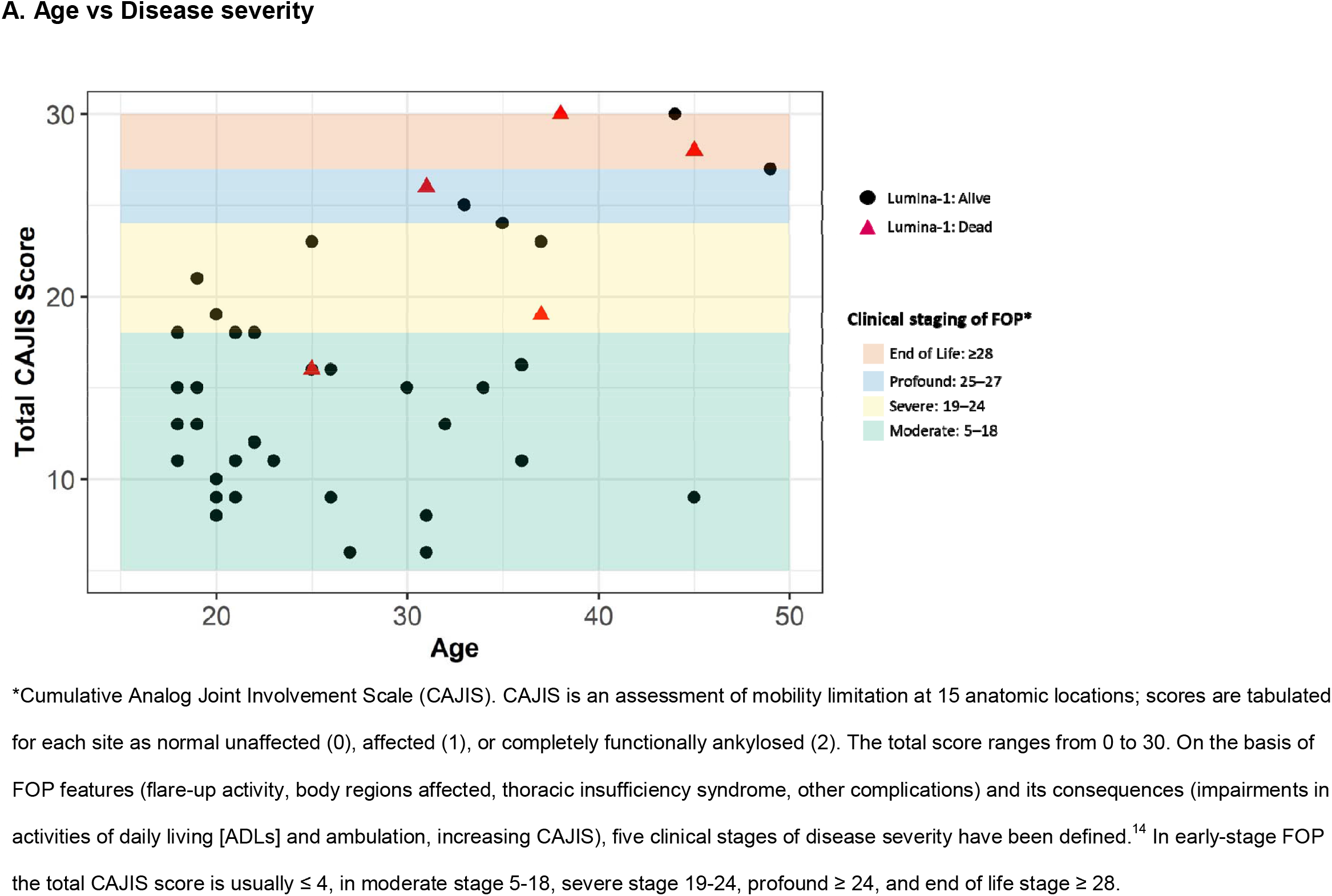
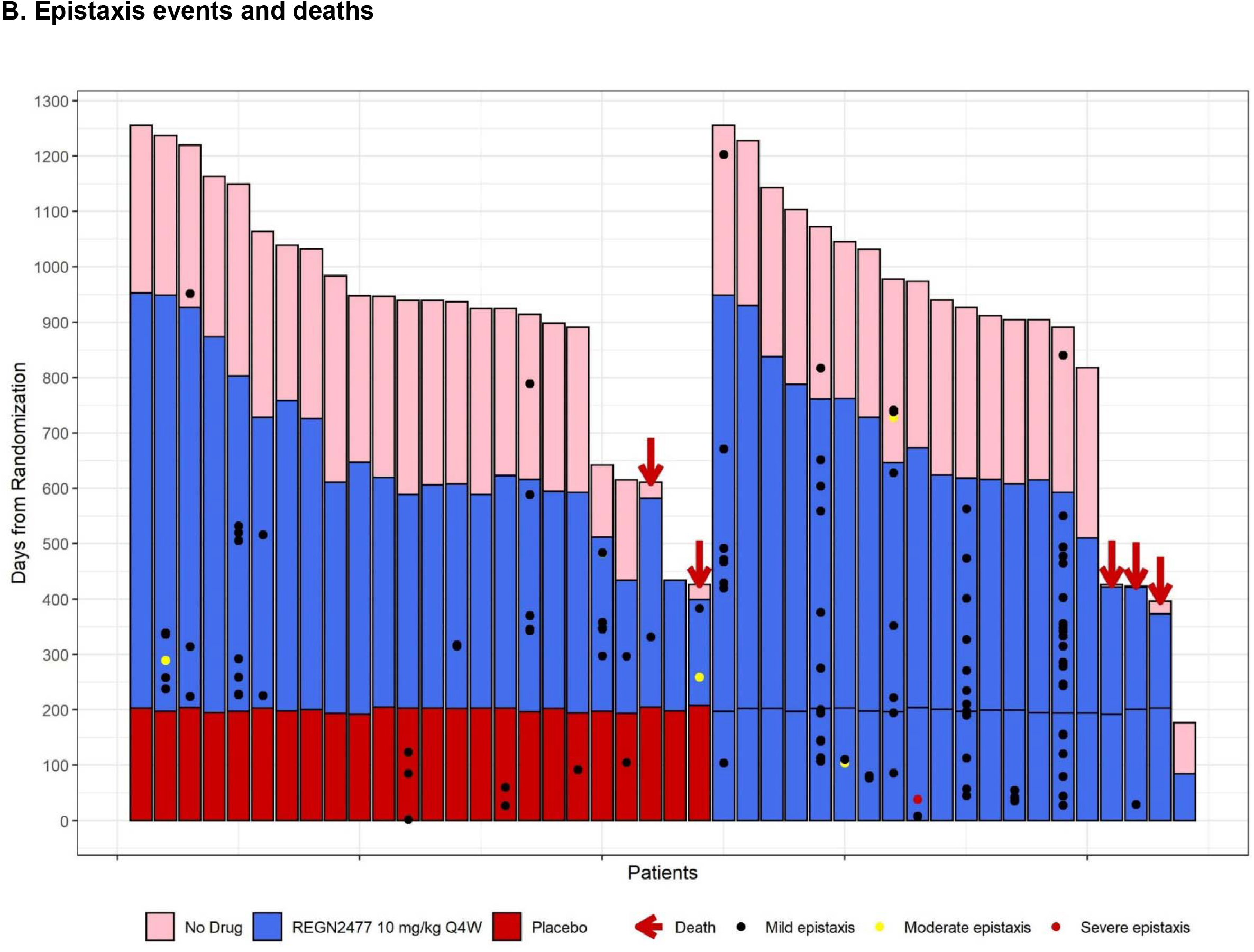
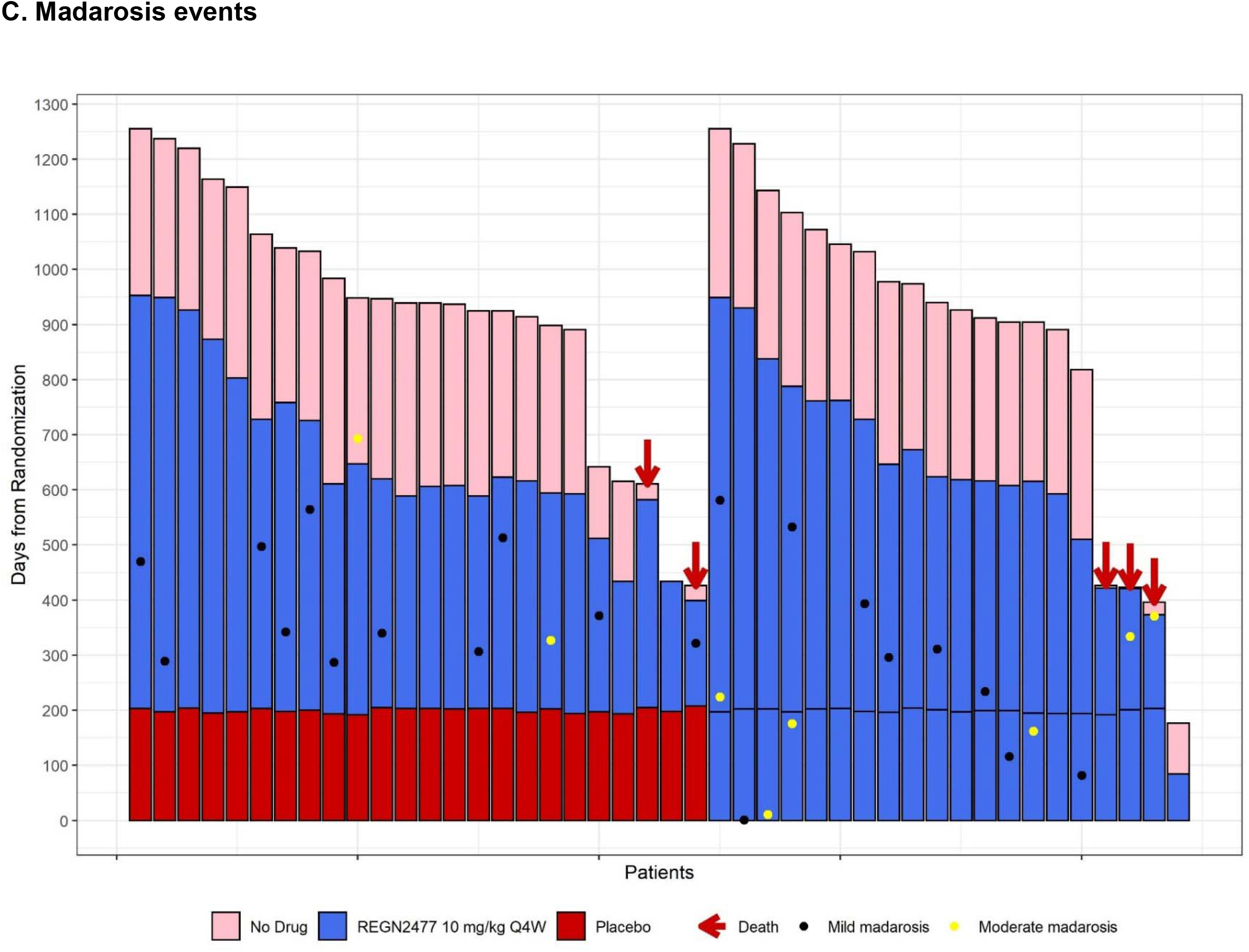
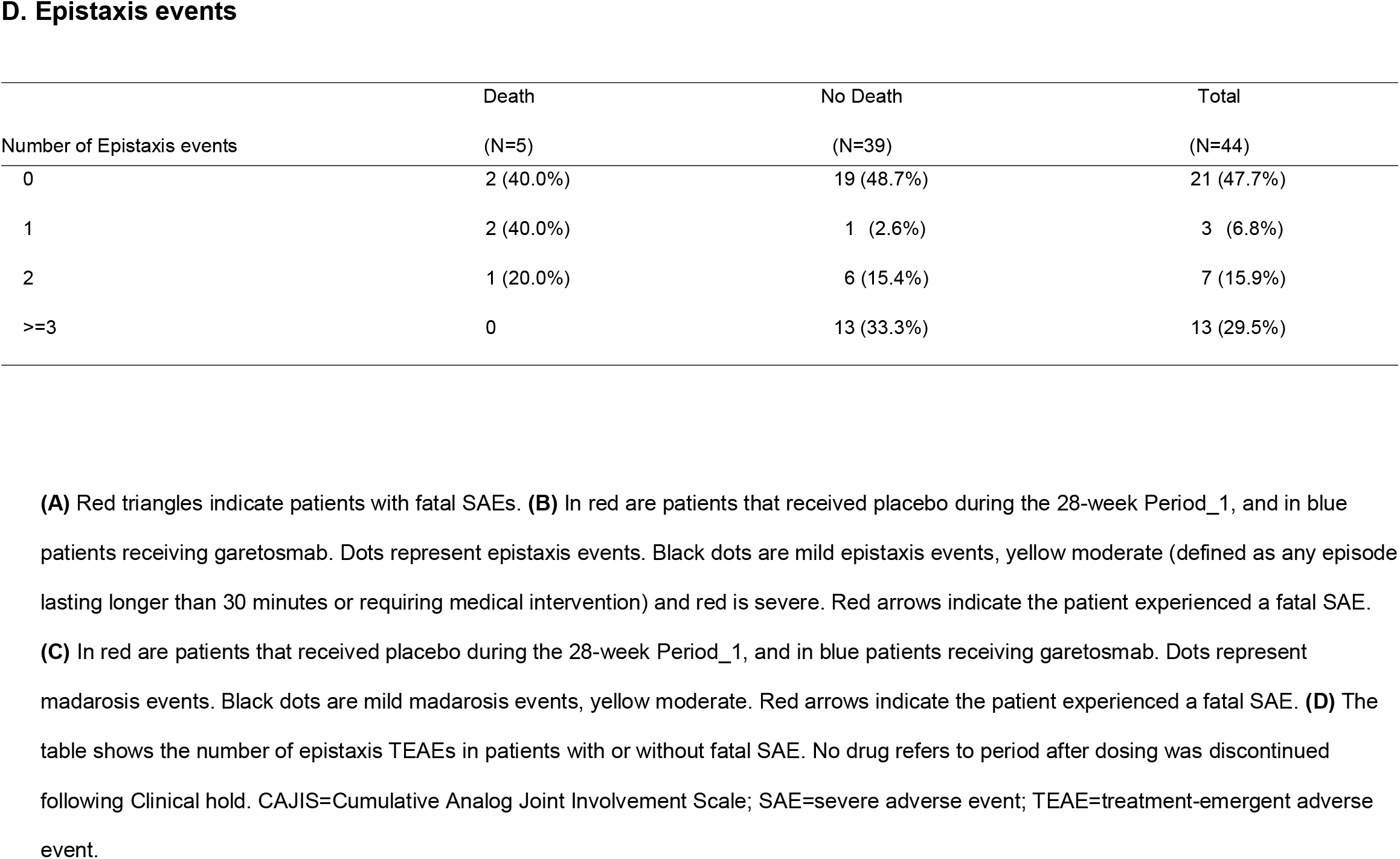
(A) Relationship between patient baseline age and disease severity as measured by the clinical staging of FOP (B) Lack of association of epistaxis events and deaths in LUMINA-1 study (C) madarosis events in the LUMINA-1 study (D) number of epistaxis TEAEs in patients with or without fatal SAE.

### Safety

All patients in the LUMINA-1 trial reported ≥1 AE during Period_1 (**Table 2**). One patient with a medical history of restrictive lung disease, pulmonary congestion, and mucus plugging discontinued during Period_1 due to an TEAE of mild pyrexia that followed recurrent episodes of pneumonia and hemoptysis. Most AEs were considered by investigators to be mild to moderate in severity. In Period_1, notable imbalances in Aes with garetosmab versus placebo included epistaxis (50.0% versus 16.7%), madarosis (loss of eyebrows/eyelashes; 30.0% versus 0%), and a composite of skin and soft tissue infections that included acne (65% versus 12.5%; **Table S7; Table S8**). Nine serious adverse events (SAEs) in Period_1 occurred in 6 (13.6%) patients; of these, 4 (20.0%) patients received garetosmab versus 2 (8.3%) patients who received placebo (**Table S9**). SAEs in the garetosmab group included 3 (15.0%) patients with infections and infestations, 1 (5.0%) patient with epistaxis (5.0%), and 1 (5.0%) with intestinal obstruction. The SAE of epistaxis resulted in hospitalization for nasal packing and was assessed as related to garetosmab by the investigator, but patient required no transfusions and had no drop in hemoglobin. The patient fully recovered and continued in the study without recurrence. This SAE of epistaxis led to a protocol amendment to include additional exclusion criteria (**Table S2**) and baseline (if applicable) and post-treatment laboratory measures of coagulation parameters and platelet effector function to exclude patients who may have existing propensity for bleeding or mitigate the potential risk for epistaxis.

**Table 2.**
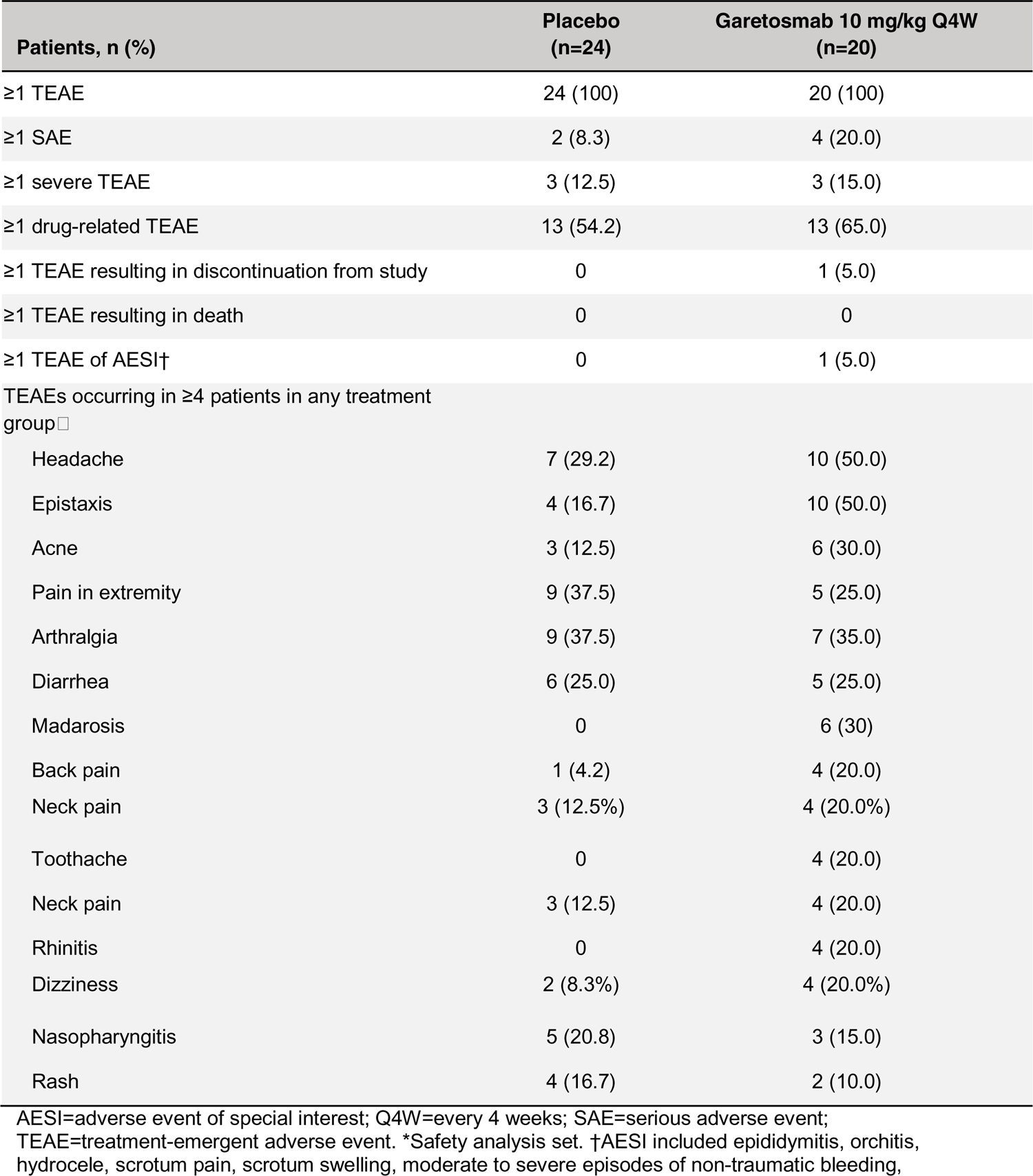
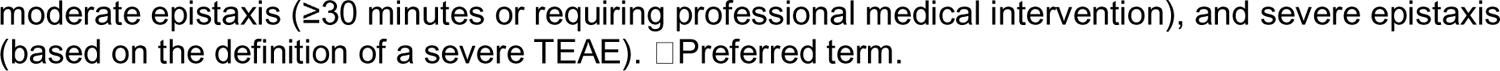
Summary of TEAEs during the double-blind period of the study (Period_1)*

| Patients, n (%) | Placebo<br>(n=24) | Garetosmab 10 mg/kg Q4W<br>(n=20) |
| --- | --- | --- |
| ≥1 TEAE | 24 (100) | 20 (100) |
| ≥1 SAE | 2 (8.3) | 4 (20.0) |
| ≥1 severe TEAE | 3 (12.5) | 3 (15.0) |
| ≥1 drug-related TEAE | 13 (54.2) | 13 (65.0) |
| ≥1 TEAE resulting in discontinuation from study | 0 | 1 (5.0) |
| ≥1 TEAE resulting in death | 0 | 0 |
| ≥1 TEAE of AESI† | 0 | 1 (5.0) |
| TEAEs occurring in ≥4 patients in any treatment group□ |  |  |
| Headache | 7 (29.2) | 10 (50.0) |
| Epistaxis | 4 (16.7) | 10 (50.0) |
| Acne | 3 (12.5) | 6 (30.0) |
| Pain in extremity | 9 (37.5) | 5 (25.0) |
| Arthralgia | 9 (37.5) | 7 (35.0) |
| Diarrhea | 6 (25.0) | 5 (25.0) |
| Madarosis | 0 | 6 (30) |
| Back pain | 1 (4.2) | 4 (20.0) |
| Neck pain | 3 (12.5%) | 4 (20.0%) |
| Toothache | 0 | 4 (20.0) |
| Neck pain | 3 (12.5) | 4 (20.0) |
| Rhinitis | 0 | 4 (20.0) |
| Dizziness | 2 (8.3%) | 4 (20.0%) |
| Nasopharyngitis | 5 (20.8) | 3 (15.0) |
| Rash | 4 (16.7) | 2 (10.0) |

AESI=adverse event of special interest; Q4W=every 4 weeks; SAE=serious adverse event; TEAE=treatment-emergent adverse event. \*Safety analysis set. †AESI included epididymitis, orchitis, hydrocele, scrotum pain, scrotum swelling, moderate to severe episodes of non-traumatic bleeding,
moderate epistaxis ( $\geq 30$ minutes or requiring professional medical intervention), and severe epistaxis (based on the definition of a severe TEAE). ☐ Preferred term.

Coagulation tests and platelet functional assays including activated prothrombin time (aPTT), prothrombin time (PTT), and prothrombin international normalized ratio (INR), measured in a subset of patients at baseline and post treatment, were in the normal range at baseline and fluctuations observed in placebo or in the garetosmab treatment group remained within the normal ranges (**Figure S7**). In Period_1, 13 (65.0%) patients in the garetosmab treatment group experienced a bleeding event compared to 9 (37.5%) patients in the placebo treatment group (**Table S10**).The only bleeding event reported in more than two patients in the garetosmab group was epistaxis. Except for the single epistaxis SAE, other events of epistaxis were non-serious and the majority mild in severity. Other non-epistaxis bleeding events were balanced between garetosmab and placebo recipients and non-serious. None of the patients discontinued therapy due to a bleeding AE.

All patients reported ≥1 AE during Period_2 and subsequent open label extension, which involved patients receiving garetosmab. The most frequently reported AEs were consistent with those reported in Period_1, comprising of skin and soft tissue infections (81.4%; acne (32.6%), madarosis (46.5%)) and epistaxis (34.9%), and were mostly mild/moderate in severity (**Table S8; Table S11; Figure 3B and 3C**). During Period_2 and subsequent treatment until end of study, epistaxis was reported in 15 patients (34.9%). Most events were mild, and no patients discontinued treatment with garetosmab due to epistaxis. Twenty SAEs in 13 (30.2%) patients were reported during Period_2 until end of study, including infections and infestations (16.3%) and gastrointestinal disorders (4.7%; **Table S12**). Five (11.6%) patients experienced 6 SAEs of abscess requiring a hospital admission or ER visit for incision and drainage. These events resolved and patients continued garetosmab after a temporary interruption of study drug; 5 of these 6 SAEs were deemed related to garetosmab by the investigator.

In order to determine if epistaxis events were related to changes in BMP/TGFß family members other than activin A known to regulate angiogenesis and vascular endothelium homeostasis, levels of BMP9 were measured.^24,25^ Minor fluctuations in serum BMP-9 were observed in both the placebo and in the garetosmab treatment group. The onset of epistaxis occurred in patients with stable and fluctuating serum BMP-9 concentrations. (**Figure S8**).

The percentage of patients with infusion reactions was balanced between garetosmab (5/20 [25%]) and placebo (6/24 [25%]). Except for two infusion reactions of moderate severity that occurred in patients who received garetosmab during Period_1, the other infusion reactions were reported as mild. Some reactions required a temporary infusion interruption or antihistamine (loratadine) pre-medication (n=2), but all infusions were completed. No patient discontinued treatment due to infusion reactions and none of the infusion reactions were associated with signs or symptoms of anaphylaxis or development of antidrug antibodies.

There were five deaths (5/44; 11.4%) during the study, all of which occurred in Period_2 (Open label) or in the open label extension between 8-16 (median-15) doses of garetosmab; none occurred during the double-blind period (Period_1). The deaths were reported by investigators as unrelated to garetosmab. The case summaries can be found in Supplement **Table S13**. As can be seen in **Figure 3B and 3D**, there was no relationship between subjects who died and the frequency or severity of epistaxis. The causes of death were head and brain trauma due to a fall in the setting of severe motor disability; hemorragic stroke in the setting of poorly controlled hypertension and previous strokes; fatal intestinal obstruction in the setting of a previous episode of intestinal obstruction; cardiac arrest during surgery procedure for traumatic spleen rupture due to fall; sudden cardiac death in a patient with lung granulomatous inflammation most likely attributable to chronic pulmonary aspiration.

Of the five deaths, three patients had a CAJIS score ≥24 and either profound or end-of-life disease severity as measured by the clinical staging of FOP developed by Pignolo & Kaplan.^14^ Among the fatal events in study participants with lower CAJIS scores, one occurred in a 26 to 30-year-old patient with a CAJIS score of 16 (moderate disease staging) who died from a fall resulting in severe head and brain trauma. This individual had significant rigidity and walking disability at baseline. Another was a 36 to 40-year-old patient with a CAJIS score of 19 (severe disease staging) who died from apparent sudden cardiac arrest with extensive granulomatous formation in the lungs consistent with chronic aspiration of foreign material.

## Discussion

This phase 2 study was conducted to assess the safety and efficacy of garetosmab in patients with FOP. The initial hypothesis for Period_1 was that garetosmab prevents formation of new lesions and progression of pre-existing lesions. This was based on preclinical data demonstrating that activin A is required for the formation of new heterotopic bone lesions, as well as continued growth and expansion of pre-existing, yet nascent lesions.^17,18^ Data from Period_1 demonstrated that the most pronounced effect of garetosmab was the suppression of the formation of new HO lesions. Garetosmab did not significantly affect the continued growth and expansion of pre-exisiting HO lesions. Contrary to our apriori hypothesis, these established lesions had already achieved maturity prior to the administration of garetosmab as evidenced by no significant growth in the placebo arm (**Table S14**). This finding led to prospective redefining of the study hypothesis for Period_2 to be that garetosmab prevents formation of new HO lesions in patients switching from placebo to garetosmab. The hypothesis was agreed upon with the US Food and Drug Administration, along with concordance that patients exposed to placebo during Period_1 could serve as the control group when they transitioned to receive garetosmab during Period_2 given the ultra-rare nature of FOP and striking results in Period_1. Notably, no new lesions were detected in Period_2 by CT in patients transitioning to garetosmab, while efficacy persisted in those who received garetosmab during Period_1 and continued garetosmab in Period_2. This study also demonstrated that PET and CT scans were similarly well correlated in the detection of new lesions.

In addition to the significant reduction in the number of new HO lesions, treatment with garetosmab demonstrated a significant effect on the incidence and frequency of soft-tissue inflammatory flare-ups based upon both patient diary and investigator adverse event reporting. Flare-ups are a significant burden for patients with FOP, often are painful and debilitating and require the use of corticosteroids in high doses or during prolonged periods of time. The reduction in flare ups by garetosmab suggests that flare-ups are also associated with the activin A pathway and highlights an additional potential utility of garetosmab as a disease-modifying and steroid-sparing treatment for patients with FOP.

Data from patients treated with garetosmab in both Period_1 and Period_2 highlight the durability of garetosmab in preventing the formation of new HO lesions, as evident from both CT and PET scan results. This effect by garetosmab was sustained through week 76.

The clinical program for garetosmab included two previous phase 1 first-in-human studies conducted in healthy volunteers (Clinicaltrials.gov: NCT02870400, NCT02943239) that showed garetosmab in multiple doses up to 10 mg/kg (single dose) and 10 mg/kg Q4W for 4 doses were well-tolerated. In one study of 30 healthy females treated with garetosmab and 10 randomized to placebo, there were no occurrences of epistaxis or skin and soft tissue infections, suggesting that these effects of garetosmab maybe specific to FOP.^23^ In another phase 1 study (NCT02943239) combining garetosmab with an anti-myostatin, GDF8 mAb (n=82), there was a single epistaxis event and 2 episodes of postmenopausal bleeding events. Overall there were 3 (3.75%) bleeding event AEs (1 epistaxis and 2 episodes of postmenopausal bleeding) amongst 122 volunteers which were all non-serious and mild in severity. The review of the healthy volunteer data from Phase I clinical trials did not suggest a signal between garetosmab and epistaxis although the exposure to garetosmab was brief.

Overall, in this study, the AEs were predominantly mild in severity, with no effect on patients’ ability to receive garetosmab. Notable AE imbalances with garetosmab during Period_1 included epistaxis, madarosis, and a composite of skin and soft tissue infections (which included skin abscesses, folliculitis, cellulitis, furuncles and carbuncles). Based on results from Period_1, higher incidences of epistaxis, madarosis, and skin and soft tissue infections were determined to be associated with garetosmab.

Further investigation shows that while there was an apparent link between garetosmab and epistaxis, there is no apparent link between garetosmab and bleeding events in other organs or systems; these events were balanced during Period_1. The great majority of epistaxis events throughout the study were mild in severity, with only 4 moderate events and one severe event. The biological mechanism behind inhibition of activin A with garetosmab and the occurrence of epistaxis in a subset of FOP patients is not understood. While activin A utilizes the same type II receptors as BMP9, which is associated with angiogenesis and vascular endothelium homeostasis,^24,25^ treatment-related changes or epistaxis events did not correlate with fluctuations in serum BMP-9 levels. Furthermore, there was no difference in serum BMP9 levels among patients with a fatal outcome. Coagulation tests and platelet functional assays were also in the normal ranges. Based on available data, no abnormalities were reported suggesting that the patients enrolled did not have coagulation defects at study entry or post garetosmab treatment.

Five fatal events were observed during Period_2 and subsequent open-label extension; in both periods patients received only garetosmab. Three of the 5 patients who died experienced epistaxis and all events were mild to moderate in severity. Two of these patients experienced one event (one of them had history of epistaxis prior to study enrollment, and the other had medical history of brain cavernoma hemorrhage as well as pigmented purpuric dermatosis, which is an inflammatory dermatitis with vascular fragility); the third patient reported 2 epistaxis events (**Figure 3B**).

Of the five fatalites, two of the deaths were due to complications resulting from falls, which is common in patients with FOP. One death was due to intestinal obstruction which has been previously reported as a complication of FOP.^15^ One death occurred as a result of a hemorrhagic stroke to the deep structures of the brain leading to acute respiratory failure. This patient presented with difficult-to-control arterial hypertension and interpretation of the brain CT by two independent neuroradiologists concluded evidence of chronic hypertensive effects and the findings suggested death was likely a consequence of hypertension. One death was initially considered to be related to a hemorrhagic event in the lung, which generated concern around the possible association between bleeding and deaths, given that early in the course of the study epistaxis had already been identified as a risk associated with garetosmab. As a result, a temporary pause in patient dosing was instituted to allow the sponsor time to conduct thorough investigations into the deaths, evaluate the autopsy reports performed in 2 of the 5 patients with a fatal outcome, and further assess a potential association between garetosmab and bleeding risk. The cause of death of the patient initially thought to have experienced lung hemorrhage was ultimately determined to be sudden cardiac arrest secondary to recurrent aspiration. There was no evidence of any bleeding which was further confirmed by a second independent autopsy report from a pathologist at a separate institution.

While there are published reports of mortality in FOP, there are no published reports of annualized rates of death in this community.^13^ With this in mind, post-hoc assessment of safety data by all the authors concluded: 1) While epistaxis events were related to garetosmab 2) there was no apparent association between epistaxis and patient deaths; 3) causes of death appeared consistent with known causes of death and life expectancy for patients with FOP of similar age and disease severity as measured by clinical staging.^15^ Four of the 5 patients who died on study had advanced CAJIS scores. The one relatively young patient with a lower CAJIS score was known to have significant gait difficulties due to FOP, and died from a fall down a flight of stairs not believed to be precipitated by an antecedent bleed. Garetosmab was considered unlikely to be the cause of death in these cases, which appeared more likely to have been related to the underlying severity of their FOP disease. Although a definitive link to garetosmab could not be established in this population with advanced disease, the number of deaths was relatively high for this small study. As these deaths were in the open label period, although a contribution by the drug can not be excluded, no common pathogenetic mechanism of deaths related to garetosmab has been identified.

The major impact of garetosmab in reducing new HO suggests that the greatest utility of garetosmab may be early in the disease course before substantial HO has occurred and before permanent disability ensues. Together with previously published preclinical data in murine models of FOP, these data indicate that activin A is also required for the continued growth and progression of new lesions. The reduction in flare-ups provides an additional and important benefit by significantly reducing disease burden. Together, the data generated during LUMINA-1 demonstrate that activin A is a required ligand for HO in FOP, and provides strong evidence that inhibition of activin A using garetosmab is a promising disease-modifying therapy with the ability to not only block HO but also reduce the number and severity of flare-ups. In the context of this ultra-rare disease setting, the patient population was small with limited patient exposure. A phase 3 adult study and a pediatric study are planned to continue to assess the benefits and risks of garetosmab intervention in people with FOP, and further validate the results from this small phase 2 study.

## Conclusions

Garetosmab treatment has demonstrated substantial and persistent reduction of new heterotopic bone lesion formation and flare-ups in adults with FOP. Although a definitive link between garetosmab and the five deaths was not established in this population with advanced disease, the number of deaths was relatively high for this small study, therefore the benefit-risk of garetosmab will be further evaluated in Phase 3 clinical studies. Garetosmab may provide a therapeutic option in this ultra-rare, severely debilitating, life-threatening disease and further studies are planned.

## Supporting information

CONSORT checklist

## Data Availability

Qualified researchers may request access to study documents that support the methods and findings reported in this manuscript. Individual anonymized patient data will be considered for sharing once the product and indication has been approved by major health authorities (eg, FDA, EMA, PMDA, etc), if there is legal authority to share the data and there is not a reasonable likelihood of patient re-identification. Submit requests to https://vivli.org/.

## Consortium Author Information

### Disclosures

Maja Di Rocco: PI of Regeneron Pharmaceuticals Inc. and Ipsen trials.

Eduardo Forleo-Neto: employee of and holds stocks and shares in Regeneron Pharmaceuticals Inc.

Robert J. Pignolo: PI of the Regeneron Pharmaceuticals Inc. LUMINA-1 and Clementia/Ipsen MOVE trials, founding member and immediate past president of the International Clinical Council (ICC) on FOP, and chair of the ICC Publications Committee.

Richard Keen: PI of clinical trials sponsored by Clementia/Ipsen and Regeneron Pharmaceuticals Inc; non-paid member of the International Clinical Council on FOP and IFOPA registry advisory board.

Philippe Orcel: PI of clinical trial sponsored by Regeneron Pharmaceuticals Inc.

Thomas Funck-Brentano: is a sub-investigator of the Regeneron Pharmaceuticals Inc. LUMINA-1 trial and PI of IPSEN FALKON trial

Christian Roux: research grants, for the institution, from Regeneron Pharmaceuticals Inc.

Sami Kolta: sub-investigator of the Regeneron Pharmaceuticals Inc. LUMINA-1 trial.

Annalisa Madeo: sub-Investigator in clinical trials sponsored by Regeneron Pharmaceuticals Inc. and Clementia-Ipsen.

Judith S. Bubbear: sub-investigator of the Regeneron Pharmaceuticals Inc. LUMINA-1 and Clementia/Ipsen MOVE trials.

Jacek Tabarkiewicz: grants to institution (UR), speaker for Merck, Novartis; hired scientific expert for SoftSystem.

Malgorzata Szczepanek: speaker for Roche.

Javier Bachiller-Corral: investigator of clinical trial sponsored by Regeneron Pharmaceuticals Inc.

Angela M. Cheung: grant to institution (UHN) for clinical trial, no personal COI.

Kathryn M. Dahir: PI on the Regeneron Pharmaceuticals Inc. Trial. This project described was supported by CTSA award No. UL1 TR002243 from the National Center for Advancing Translational Sciences. Its contents are solely the responsibility of the authors/sponsor and do not necessarily represent official views of the National Center for Advancing Translational Sciences or the National Institutes of Health.

Esmée Botman: is a sub-investigator of the Regeneron Pharmaceuticals Inc. LUMINA-1 trial at the Amsterdam UMC, the Netherlands.

Pieter G Raijmakers: is a sub-investigator of the Regeneron Pharmaceuticals Inc. LUMINA-1 trial at the Amsterdam UMC, the Netherlands.

Mona Al Mukaddam: PI of clinical trials sponsored by Clementia/Ipsen, Regeneron Pharmaceuticals Inc. and Incyte; non-paid member of the International Clinical Council on FOP and IFOPA registry advisory board.

Lianne Tile: grant to institution (UHN) for clinical trial (sub-investigator), no personal COI.

Cynthia Portal-Celhay: employee of and holds stocks and shares in Regeneron Pharmaceuticals Inc.

Melissa Simek-Lemos: employee of and holds stocks and shares in Regeneron Pharmaceuticals Inc.

Neena Sarkar: employee of and holds stocks and shares in Regeneron Pharmaceuticals Inc.

Peijie Hou: employee of and holds stocks and shares in Regeneron Pharmaceuticals Inc.

Bret Musser: employee of and holds stocks and shares in Regeneron Pharmaceuticals Inc.

Anita Boyapati: employee of and holds stocks and shares in Regeneron Pharmaceuticals Inc.

Scott Mellis: employee of and holds stocks and shares in Regeneron Pharmaceuticals Inc.

Andrew J. Rankin: employee of and holds stocks and shares in Regeneron Pharmaceuticals Inc.

Aris N. Economides: employee of and holds stocks and shares in Regeneron Pharmaceuticals Inc.

Dinko Gonzalez Trotter: employee of and holds stocks and shares in Regeneron Pharmaceuticals Inc.

Gary Herman: employee of and holds stocks and shares in Regeneron Pharmaceuticals Inc.

David M. Weinreich: employee of and holds stocks and shares in Regeneron Pharmaceuticals Inc.

George D. Yancopolous: employee of and holds stocks and shares in Regeneron Pharmaceuticals Inc.

E. Marelise W. Eekhoff: subsidies/financing FOP research: Dutch FOP Patient Foundation, IFOPA, Regeneron Pharmaceuticals Inc., EU-IMI (AZ), Clementia/Ipsen (i.e., general lecture). Non-paid Board memberships: the International Clinical Council on FOP, IFOPA registry advisory board, Dutch Society for Endocrinology (NVE) BoNe; Representative: Amsterdam Bone Center and Rare Bone Expert Center, European FOP consortium. Member of ERN BOND and of an ASBMR committee.

Frederick S. Kaplan: founding member and past president of the International Clinical Council on FOP, member of the Medical Advisory Board of the IFOPA Global Registry and global principal investigator on the Regeneron LUMINA-1 and the Clementia/Ipsen MOVE trials.

## Acknowledgements

We thank the patients and their families for their participation in this study and the healthcare professionals and investigators who treated these patients and made this study possible.

We acknowledge the contributions of Kevin Niswender, MD, PhD; Edward Sherwood. MD; Lana Howard, BSN,RN, CCRP; Margo Black, MSN, RN, CCRP, at Vanderbilt University Medical Center.

We acknowledge the contributions of Staci Kallish DO, Katherine S.Toder CCRC, and Renee Jurek CRC at the University of Pennsylvania.

We thank Donal O’Donovan Ph.D. for the safety figure concept and Richa Attre Ph.D. from Regeneron Pharmaceuticals, Inc. for assistance with development of the manuscript. Medical writing support under the direction of the authors was provided by Caroline Ridley, PhD, Prime Global (Knutsford, UK) according to Good Publication Practice guidelines (<u>Link</u>) and funded by Regeneron Pharmaceuticals, Inc.

The authors were involved in the study design and collection, analysis, and interpretation of data. All authors had full access to all the data in this study and take complete responsibility for the integrity of the data and the accuracy of the data analysis.

## Notes

### Clinical Trial

NCT03188666

### Funding Statement

The study was funded by Regeneron Pharmaceuticals, Inc.

### Author Declarations

The trial was approved by the following Institutional Review Boards: University Health Network 700 University Ave.10th Floor, Suite 1056, Toronto Ontario, M5G1Z5, Canada; Comité de Protection des Personnes (CPP) Ile-de-F, 78 rue du Général Leclerc, Le Kremlin Bicentre, Paris France, 94275; Comitato Etico Regione Liguria, IRCCS Ospedale Policlinico, San Martino, Largo Rosanna Benzi, 10, Genova Italy, 16132; Science committee AMS, Attn. Dr. R.T.de Jongh, VUmc, internal medicine, room 4A35, De Boelelaan 1117, 1081 HV Amsterdam, The Netherlands; METC VUmc BS7, Kamer H-443, Postbus 7057, Amsterdam Netherlands, 1007 MD; Komisja Bioetyczna Uniwersytetu Rzeszowskiego ul. Warszawska 26A, 35-205, Rzeszow Poland; Comité de Ética de la Investigación con medicamentos del Hospital Universitario Ramón y Cajal. Ctra. Colmenar, km. 9,100, Madrid Spain, 28034; London - Central Research Ethics Committee, 3rd Floor, Barlow House, 4 Minshull Street, Manchester UK, M13DZ; University of Pennsylvania, Office of Regulatory, 3624 Market Street, Suite 301 S, Philadelphia PA, 19104, United States; Mayo Clinic Institutional Review Board, 200 First Street SW, Rochester Minnesota, 55905, United States; Vanderbilt University 1313 21st Ave., South, Suite 505, Nashville Tennessee, 37232, United States.

### Summary of Updates

Minor edit

