## Supplement for "Garetosmab, an inhibitor of activin A, reduces heterotopic ossification and flare-ups in adults with fibrodysplasia ossificans progressiva: a randomized, double-blind, placebo-controlled phase 2 trial"

### DiRocco LUMINA1 manuscript Supplemental

#### Figure S1. Activin A activates signaling by FOP-mutant ACVR1 but inhibits signaling by wild type ACVR1

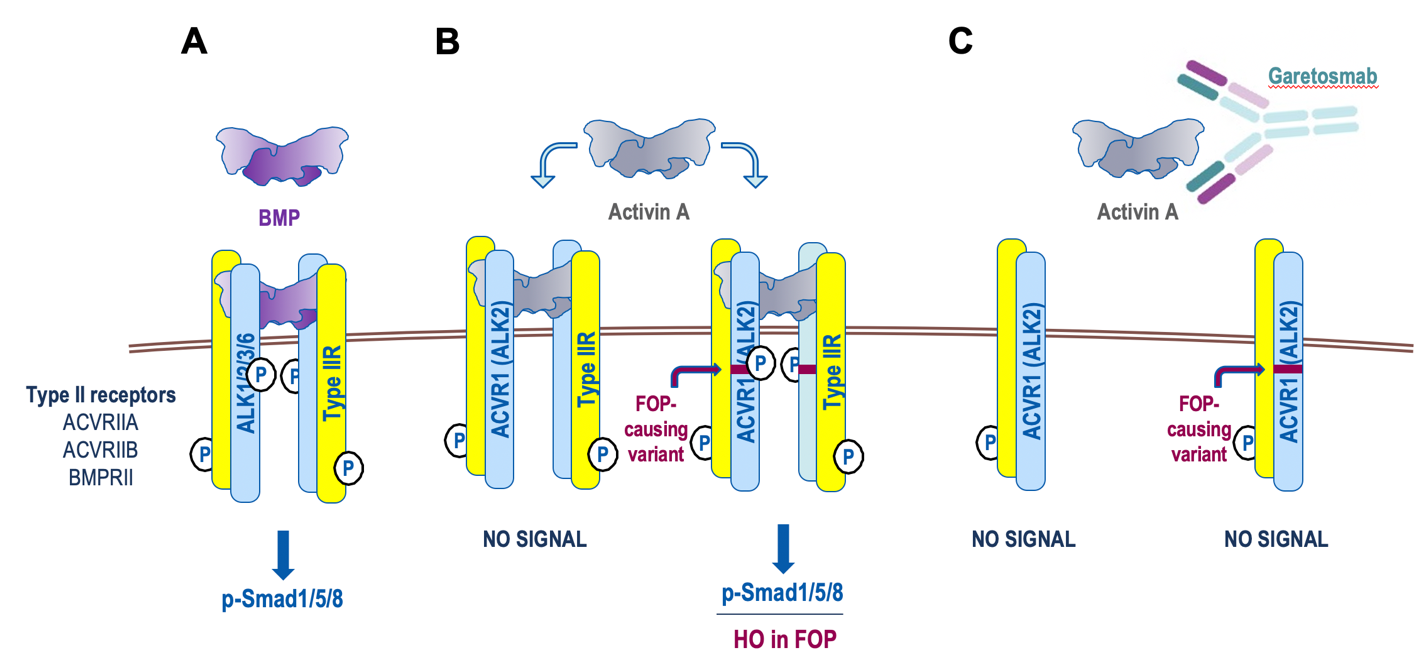

(**A**) In the presence of BMPs, ACVR1 forms a signaling complex together with its cognate type II receptors, ACVR2A, ACVR2B, and BMPR2. This complex activates signaling, mainly via phosphorylation of Smad1/5/8. (**B**) In the presence of Activin A, wild type ACVR1 forms a non-signaling complex that antagonizes signaling by BMPs. When ACVR1 is an FOP-causing variant, the non-signaling complex is converted into a signaling complex and transduces signal much like the complex formed by BMPs. (**C**) Garetosmab blocks the interaction of Activin A with its cognate type I receptors ACVR1 (ALK2) and ACVR1B (ALK4, not depicted), and hence inhibits the formation of both signaling and non-signaling complexes that would normally be generated by Activin A and the corresponding type I and type II receptor pairs. BMP=bone morphogenetic protein; FOP=fibrodysplasia ossificans progressiva; HO=heterotopic ossification; R=receptor.

#### Figure S2. Schematic overview of the study design

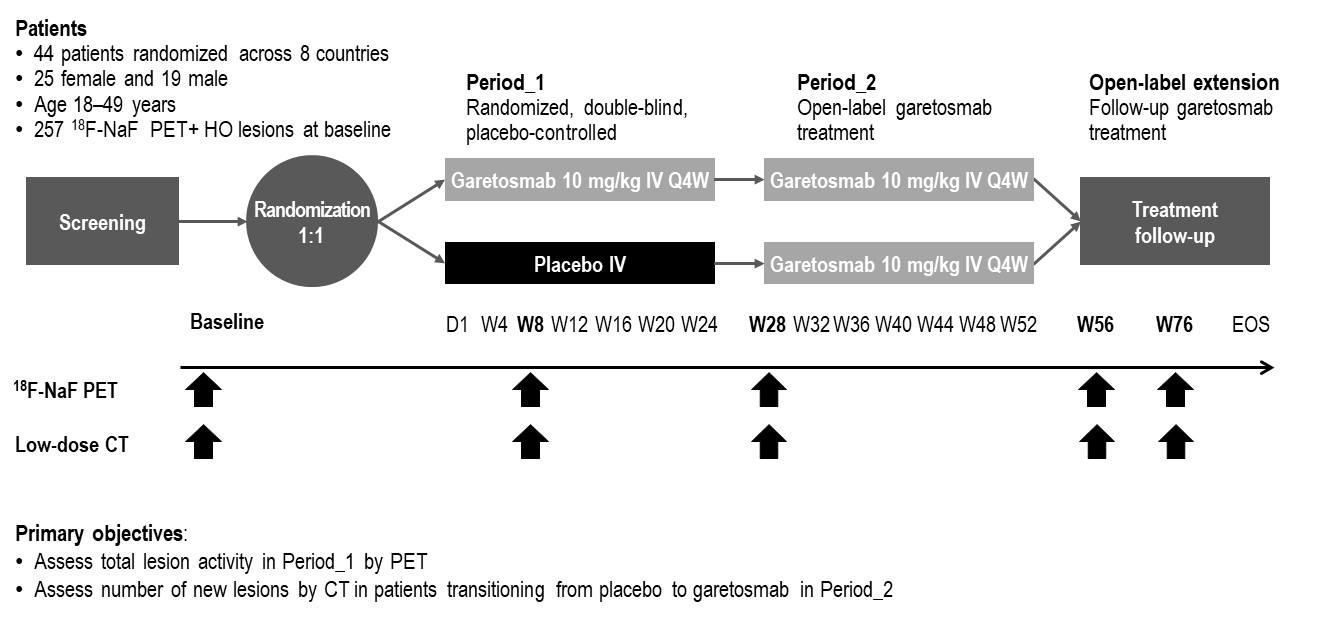

LUMINA-1 is a phase 2, randomized, double-blind, placebo-controlled study in adult patients with FOP. This study consists of a 4-week screening/baseline period, a 28-week randomized double-blind placebo-controlled treatment period (Period_1), a 28-week open-label garetosmab treatment period (Period_2), and a follow-up treatment period with open-label garetosmab (open-label extension). Following confirmed eligibility during the screening/baseline period (Day -28 to Day 1), patients were randomized 1:1 to 10 mg/kg garetosmab IV dosed Q4W or placebo. Efficacy is assessed by ^18^F-NaF PET and low-dose CT imaging analysis of Heterotopic Ossification (HO)at weeks 8, 28, 56, and 76. The primary analysis was conducted when all the patients completed the double-blind treatment (Period_1). ^18^F-NaF PET=fluorine-18-labelled sodium fluoride positron emission tomography; CT=computed tomography; D=day; EOS=end of study; FOP=fibrodysplasia ossificans progressiva; HO=heterotopic ossification; IV=intravenous; Q4W=every 4 weeks; W=week.

#### Figure S3. Trial profile

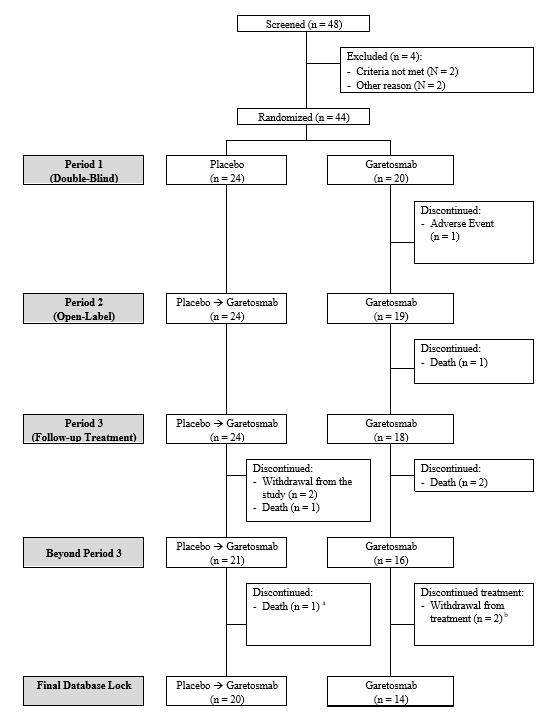

A total of 48 patients were screened and 44 patients were randomized (20 patients to garetosmab and 24 patients to placebo). One patient from the garetosmab group discontinued from the study in Period_1 due to a TEAE of pyrexia and four patients discontinued during Periods_2/3 due to potential risks associated with the COVID-19 pandemic (n=1) and a lack of perceived benefit (n=3). As of the final database lock (20 Oct 2021), 5 patients died; TEAE=treatment-emergent adverse event.

#### Figure S4. Percent of patients with new lesions by quantitative imaging in Period_1

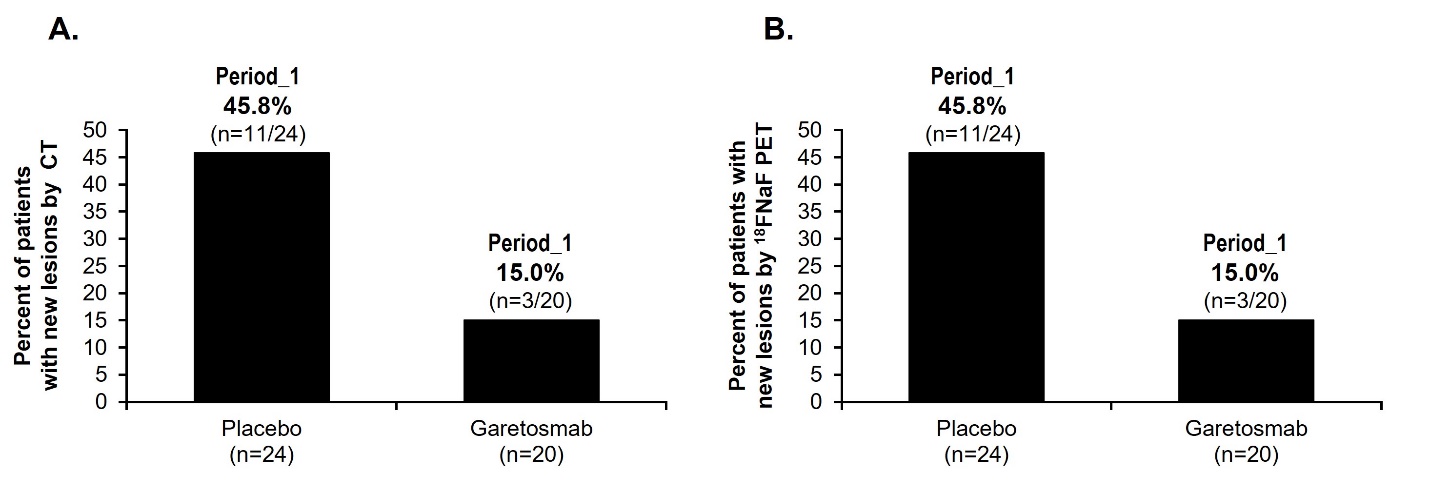

**(A)** Percent of patients with new lesions by CT during Period_1 relative to baseline (active HO analysis set). **(B)** Percent of patients with new lesions by ^18^F-NaF PET during Period_1 relative to baseline (active HO analysis set). ^18^F-NaF PET=fluorine-18-labelled sodium fluoride positron emission tomography; CT=computed tomography; mITT=modified intent-to-treat.

Figure S5. Effect of garetosmab on flare-up events
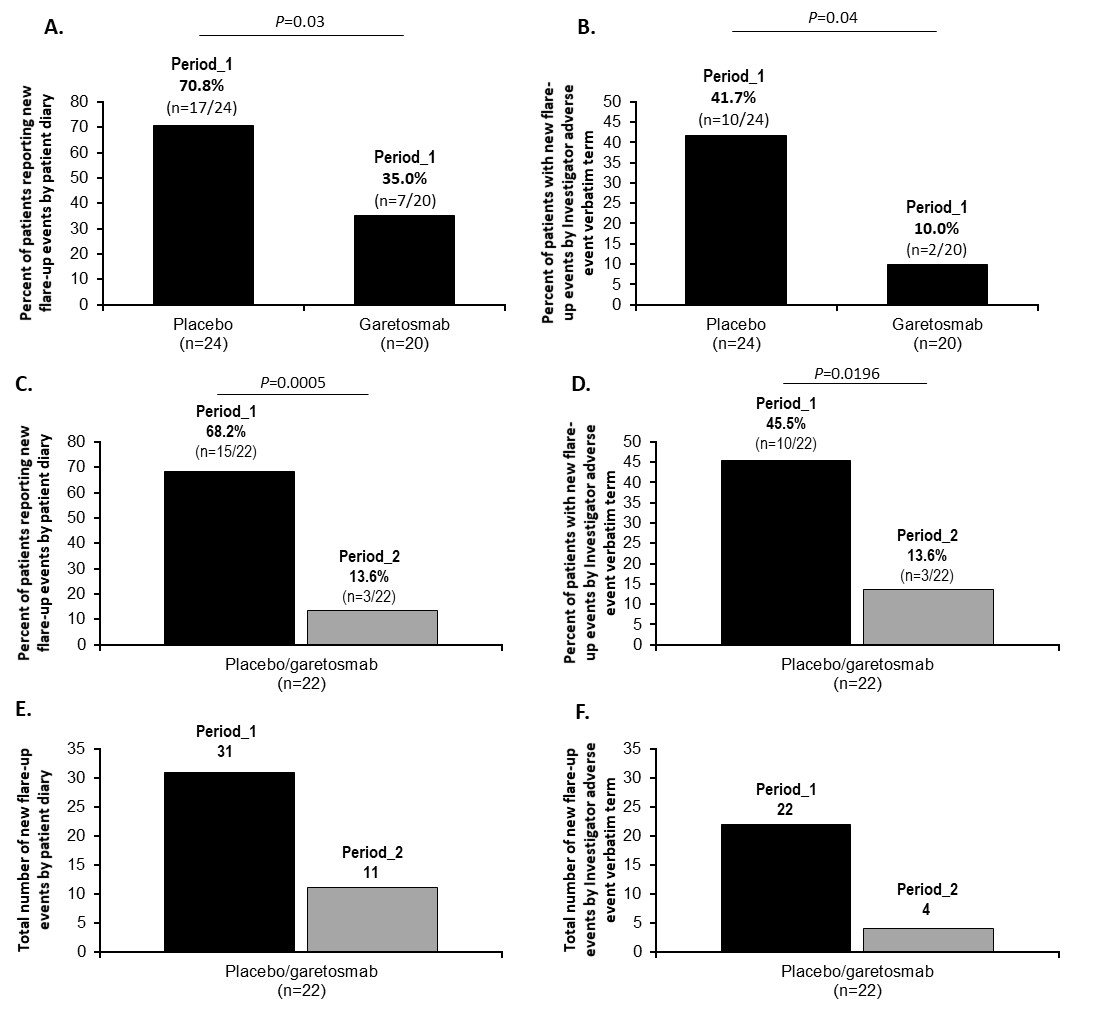

Percent of patients with new flare-up events **(A)** as reported by patient diary and **(B**) by investigator adverse event verbatim term in Period_1 (active HO analysis set). Percent of patients with new flare-up events **(C)** as reported by patient diary and **(D)** by investigator adverse event verbatim term for patients originally assigned to placebo (mITT analysis set). Total number of new flare-up events **(E)** as reported by patient diary and **(F)** by investigator adverse event verbatim term for patients originally assigned to placebo (mITT analysis set). A new flare-up was defined as a flare-up event starting in the corresponding period. Investigator-defined flare-ups were defined as TEAEs with the verbatim term containing “flare”. HO=heterotopic ossification; mITT=modified intent-to-treat; TEAE=treatment-emergent adverse event.

#### Figure S6. Effect of garetosmab in Period_2 relative to Period_1

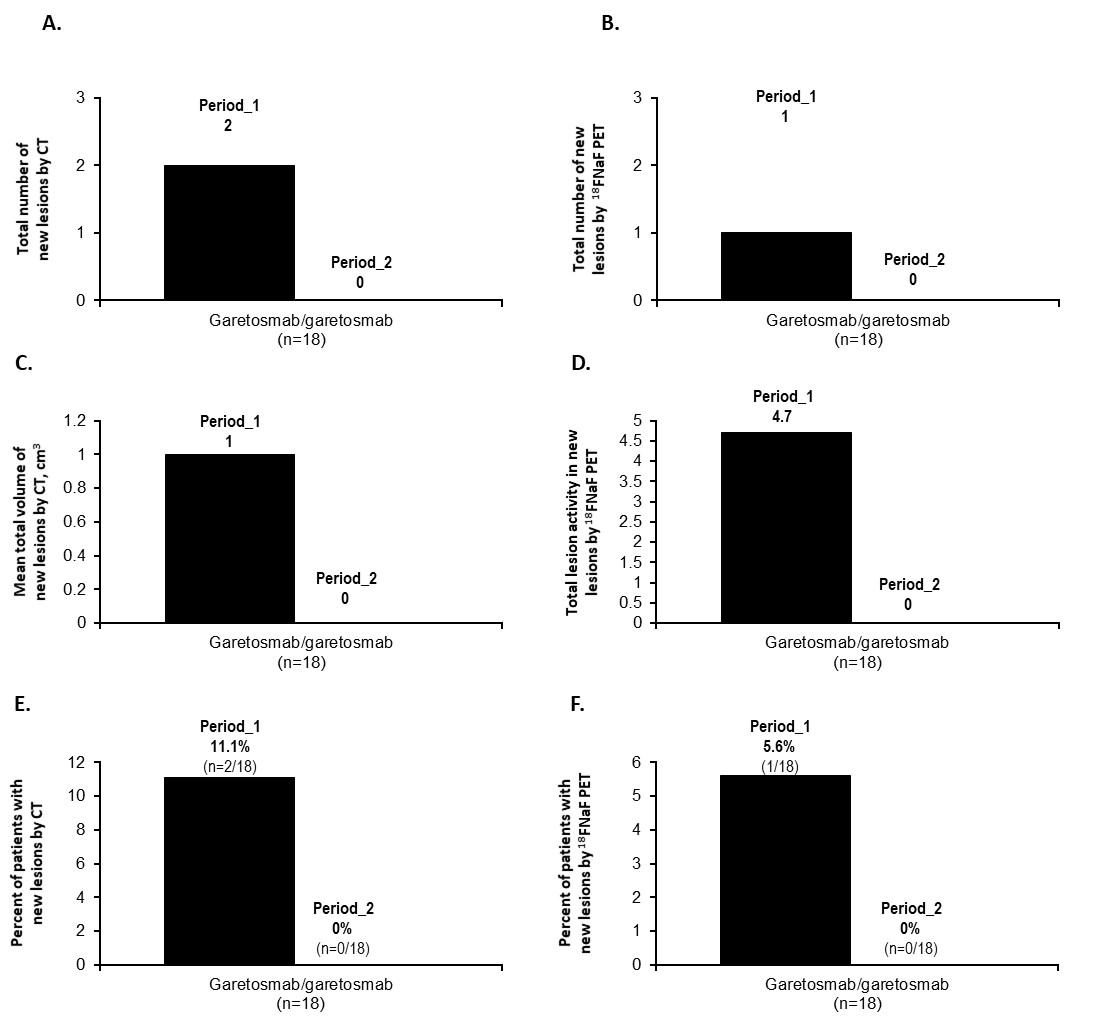

Effect of garetosmab in Period_2 relative to Period_1 on the (A, B) total number of new HO lesions, (C) mean total volume of new HO lesions (D) TLA in new HO lesions, and (E, F) percent of patients with new lesions, as assessed by quantitative imaging in patients originally randomized to garetosmab in Period_1. CT= computerized tomography; PET= positron emission tomography.

#### Figure S7. Coagulation and platelet functional assays at baseline and post treatment in LUMINA-1 patients

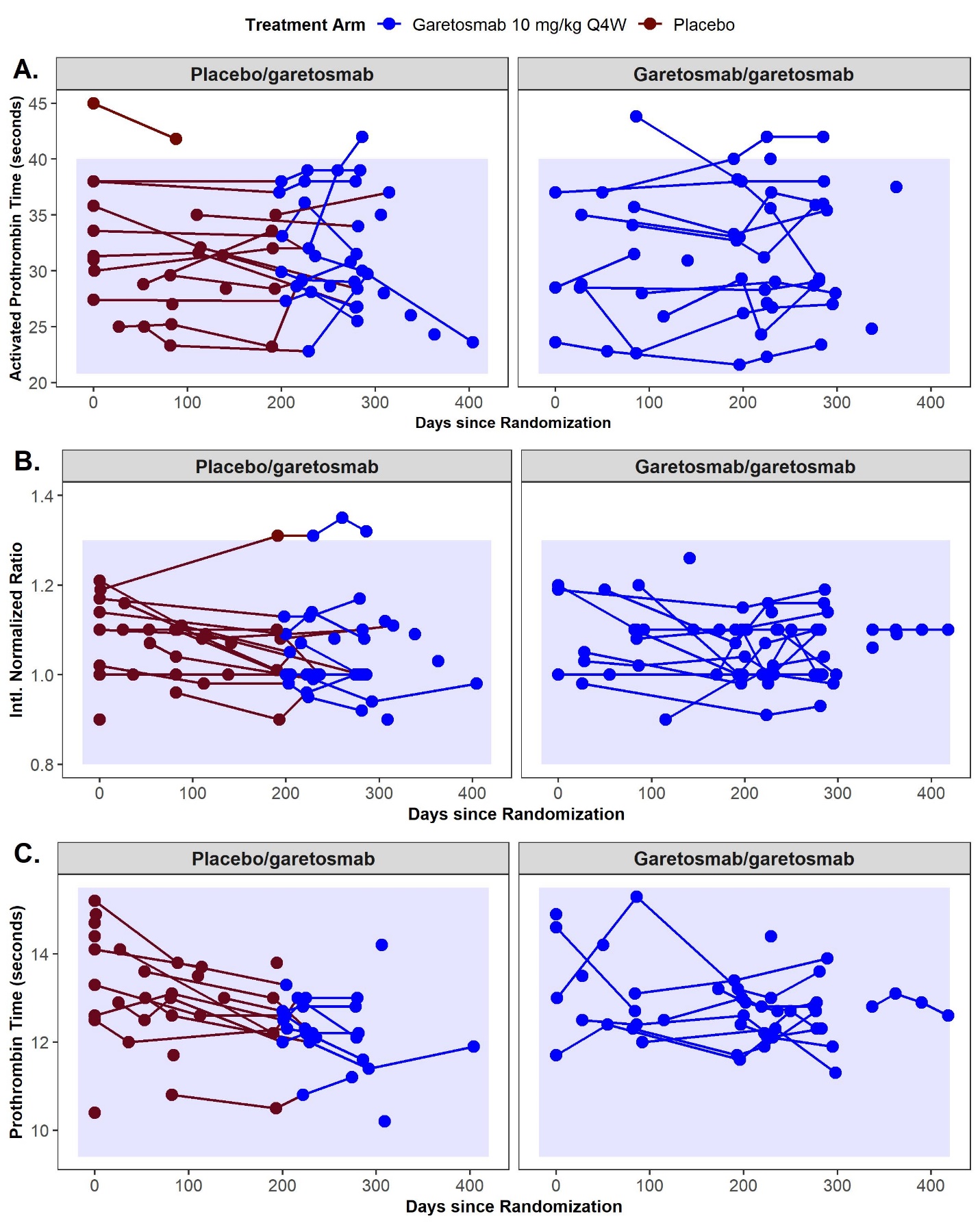

**D.**

| Treatment Assignment | Parameter | n | Mean (SD) | Median | Q1:Q3 | Min:Max |
| --- | --- | --- | --- | --- | --- | --- |
| Placebo | Activated Partial Thromboplastin Time (sec) | 9 | 34.5 (5.4) | 33.6 | 31:38:00 | 27.4 : 45 |
| Placebo | Prothrombin Intl. Normalized Ratio | 8 | 1.1 (0.1) | 1.1 | 01:01.2 | 0.9 : 1.2 |
| Placebo | Prothrombin Time (sec) | 9 | 13.6 (1.5) | 14.1 | 12.6 : 14.7 | 10.4 : 15.2 |
| Garetosmab 10 mg/kg Q4W | Activated Partial Thromboplastin Time (sec) | 3 | 29.7 (6.8) | 28.5 | 26:32.8 | 23.6 : 37 |
| Garetosmab 10 mg/kg Q4W | Prothrombin Intl. Normalized Ratio | 3 | 1.1 (0.1) | 1.2 | 1.1 : 1.2 | 01:01.2 |
| Garetosmab 10 mg/kg Q4W | Prothrombin Time (sec) | 4 | 13.6 (1.5) | 13.8 | 12.7 : 14.7 | 11.7 : 14.9 |

Coagulation tests and platelet functional assays including **(A)** activated prothrombin time (aPTT)**, (B)** prothrombin international normalized ratio (INR), and **(C)** prothrombin time were collected after protocol amendment 3 at local laboratories to explore mechanism of epistaxis and garetosmab mechanism of action. For patients already enrolled in the study, the blood samples for these assessments were collected at their next visit unless the assessment was performed in the last year. The blue box indicates the normal range across the local labs for the individual sites. **(D)** The table shows the number of measurements and summary statistics for the coagulation tests and platelet functional assays at baseline.

#### Figure S8. Changes in BMP9 levels are minor and not associated with episodes of epistaxis

**(No correlation of epistaxis with BMP-9; only 2 colors; bigger panels figures being redrawn)**

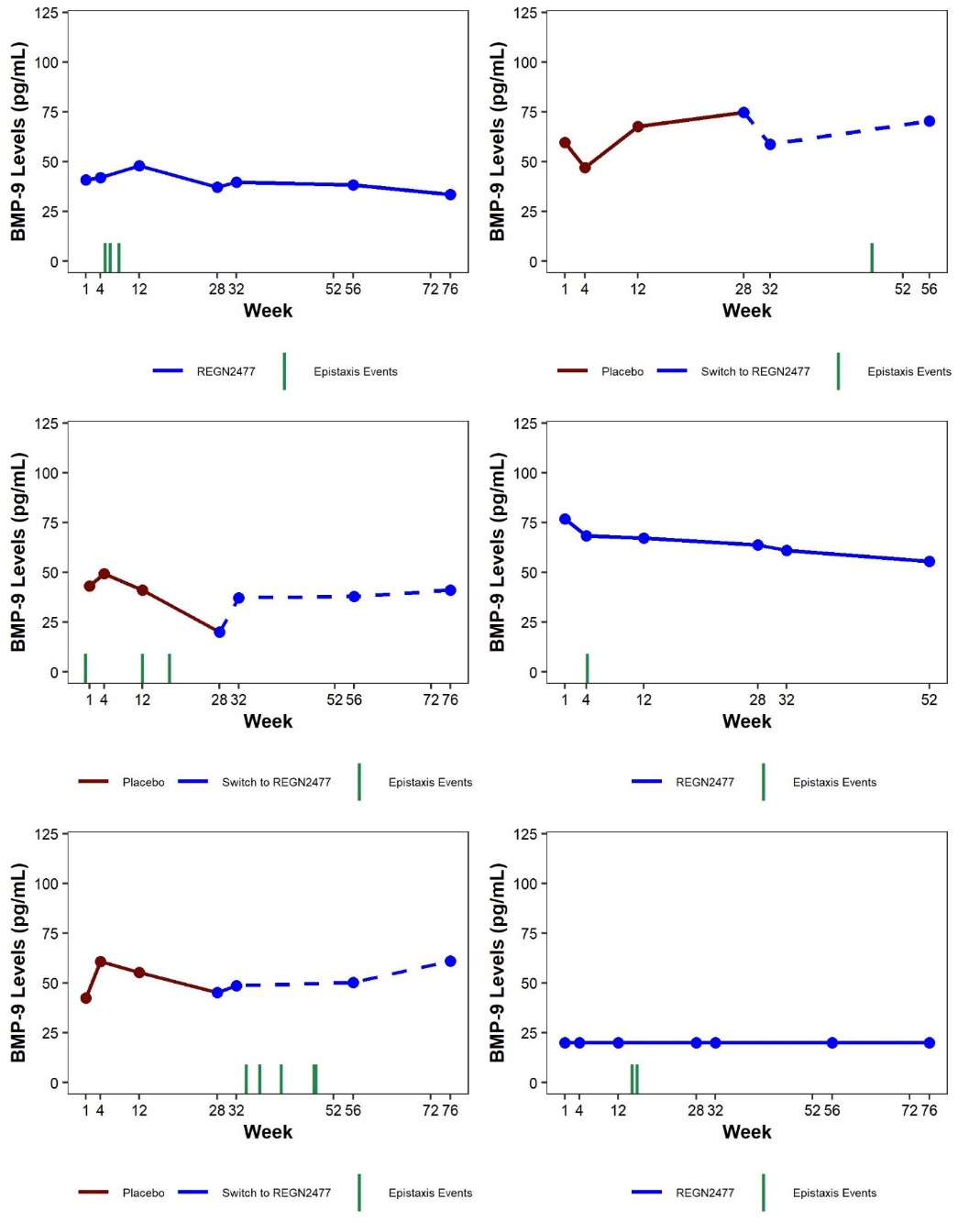

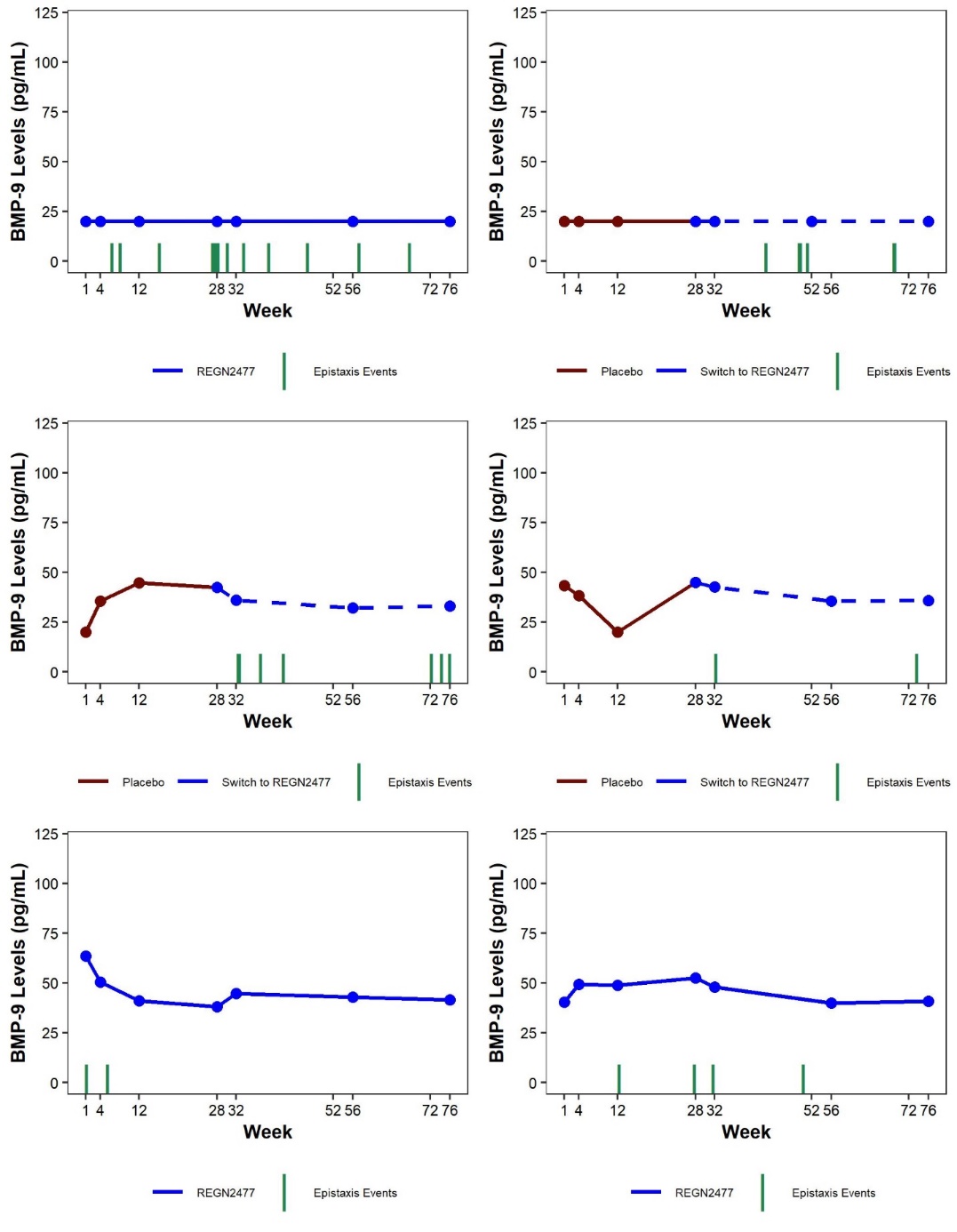

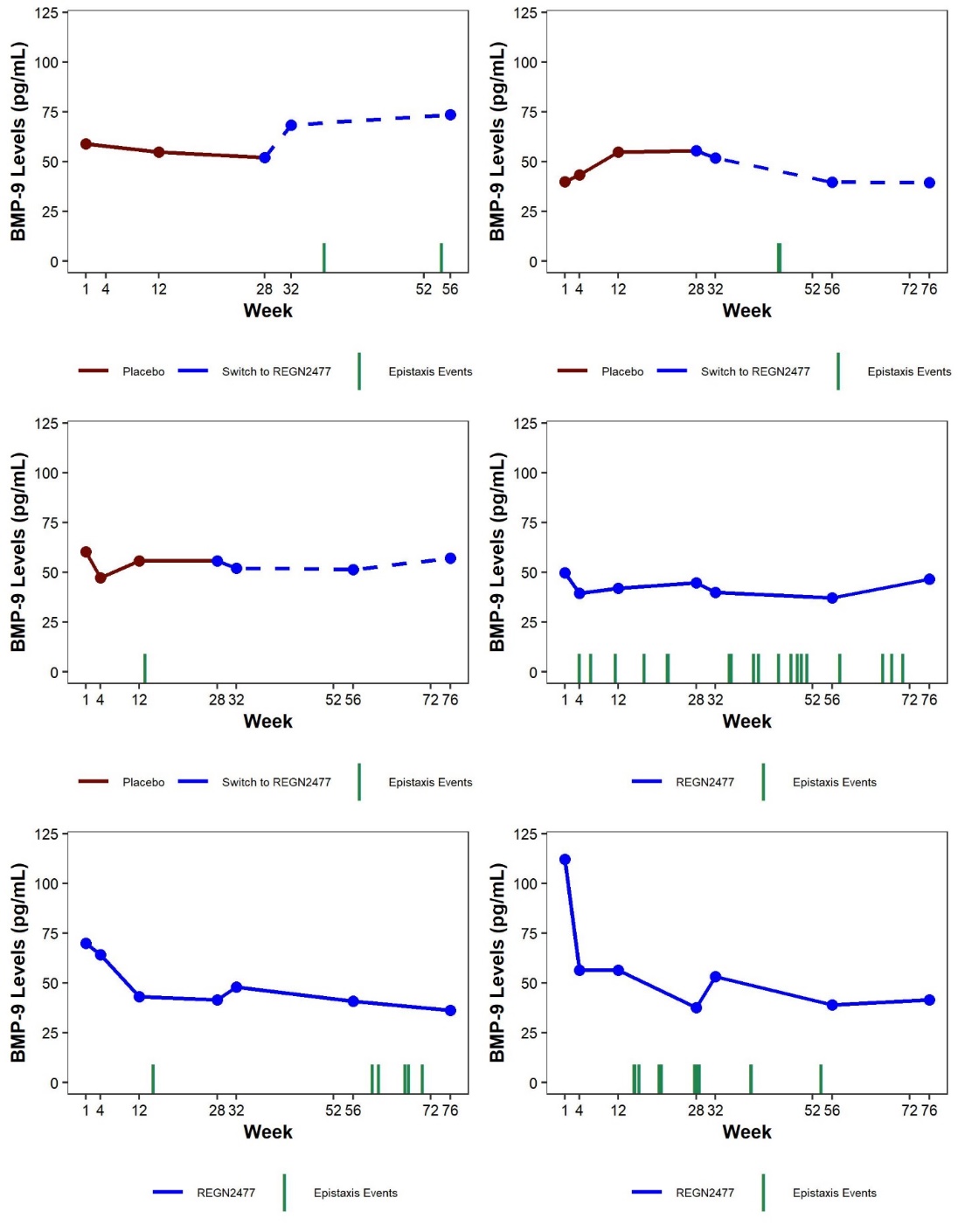

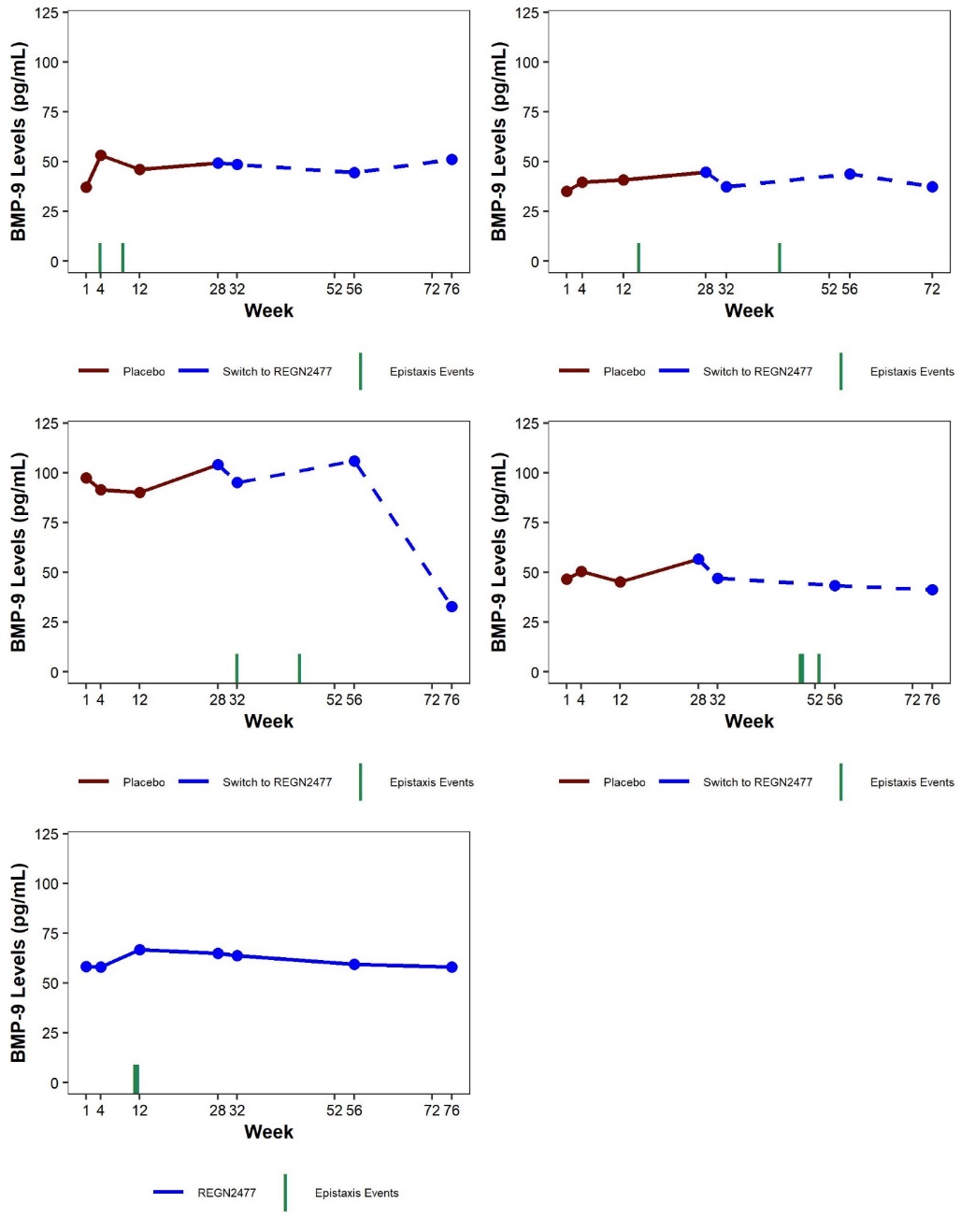

BMP-9 levels in patients who experienced TEAEs of epistaxis. Red lines indicate an epistaxis event. BL=baseline; BMP-9=bone morphogenetic protein-9; TEAE=treatment-emergent adverse event.

**Table S1. Overview of protocol and statistical methodology amendments**

|  | **Amendments to statistical methodology** | |
| --- | --- | --- |
| 1. | In Jan 2019, an urgent safety measure was implemented due to a serious event of epistaxis (resulting in hospitalization; Section 5.2.1.5) and the totality of data regarding the occurrence of epistaxis in the ongoing study at that time. This resulted in the implementation of Protocol Amendment 3 (Appendix 16.1.1), which included the following updates:  Inclusion of new exclusion criteria for patients on concomitant antiplatelet therapy, history of severe non‑traumatic bleeding, and history of bleeding diathesis (see Section 3.3.1).  Inclusion of moderate to severe episodes of non-traumatic bleeding and moderate to severe epistaxis as adverse events of special interest (AESIs; see Section 5.2.1.7).  The use of anti-inflammatory drugs that inhibited platelet function or were otherwise associated with an increased risk of bleeding was discouraged to the extent possible.  The low dose of acetylsalicylic acid was clarified (≤100 mg/day).  Baseline and post-treatment laboratory measures of coagulation parameters and platelet effector function were added.  A baseline exploratory research sample was added to explore mechanism of epistaxis and garetosmab mechanism of action.  Antiplatelet therapy, anticoagulants, and herbal supplements (that inhibit platelet function or are associated with increased risk of bleeding) were added to the list of prohibited medications. | |
| 2. | In Oct 2020, a second urgent safety measure was issued to halt garetosmab dosing after a fatal SAE occurred during treatment with garetosmab. Preliminary findings from a macroscopic post‑mortem examination suggested gross hemorrhage in the right lung. However, the final cause of death was considered sudden cardiac arrest, with additional findings of extensive granulomatous inflammation most likely attributable to chronic ongoing aspiration of foreign material (more information in Section 5.2.1.4). Four other deaths had been previously reported during the open-label period of the study (Section 5.2.1.4). Before garetosmab dosing was halted, all subjects had completed Period 3 (ie, all patients had reached 76 weeks) and all efficacy assessments had been completed. In Sep 2021, the sponsor decided to close the study (final cutoff, see Section 1.4), after all participants had performed all efficacy and other relevant assessments described in the study protocol and open‑label administration of garetosmab was not planned to be reinitiated. | |
| 3. | By the time COVID-19 was declared as a global pandemic by the World Health Organization (11 Mar 2020), all patients had completed Period 1 and most patients had completed Period 2 of the study. The sponsor decided to continue the study based on the assessment the it could be conducted without jeopardizing patient safety, data integrity, or compliance with relevant Regulatory Guidance.  Because of the public health emergency related to the COVID-19 pandemic, study procedures were adapted, and are reflected in Appendix 16.1.1, Protocol Amendment 6. Study procedures, including study drug administration, were allowed at alternative sites, which were established to minimize study patient(s)’ exposure to the SARS‑CoV‑2. It was acceptable to perform the study visit 18 (week 56) and/or visit 23 (week 76) imaging exams employing only whole-body low‑dose CT scans because of potential unavailability of the ^18^F-NaF tracer necessary for the PET scan, though this was not ultimately needed. In addition, the time window for study visit 18 (week 56) was made flexible, as long as any changes in the schedule were documented. A new dataset was also defined in Protocol Amendment 6 to mitigate the effects of the public health emergency on study outcomes, applicable to Periods 2 and 3 (COVID-19 mITT, see Section 3.7.2.1).  Formal documentation of deviations to Standard Operating Procedures or other controlled procedural documents is in Appendix 16.1.18.  An assessment of protocol deviation trending during the pandemic is in Appendix 16.1.18.  All temporary mechanisms used and deviations from planned study procedures were documented as being related to COVID-19. | |
| 4. | Upon Protocol Amendment 1, the scope of patient eligibility was expanded by opening enrollment to FOP patients with any ACVR1 mutation known to cause the disease, not only the classic ACVR1[R206H] mutation. This expansion was based on in vitro and in vivo studies that demonstrated that all the FOP-causing mutations in ACVR1 render this receptor responsive to Activin A, much like the prototypical ACVR1[R206H] variant (see Section 1.2). These results indicated that garetosmab is likely to be effective in blocking HO irrespective of the specific FOP-causing variants of ACVR1 present in any given patient (see Appendix 16.1.1, Protocol Amendment 1). To support the analyses of study results, the ACVR1 genotype initially documented by investigators at enrollment was confirmed retrospectively in a central laboratory for all patients. Efficacy of garetosmab was assessed irrespective of ACVR1 mutation and separately for the patients with ACVR1[R206H] mutation. | |
| 5. | Observations from Period 1 led to a redefinition of pre-specified hypotheses for Period 2. In Period 1, non-statistically significant reductions were observed in both HO activity (as measured by ^18^F-NaF PET) and in HO volume (as measured by CT) (see Section 5.1.1.1). Further examination of these data revealed that changes in these measures were entirely driven by activity and growth associated with new HO lesions. These observations led to redefinition of endpoints for Period 2 with focus upon the growth and activity of new HO lesions. The study hypothesis for Period 2 was defined in Protocol Amendment 6 (Appendix 16.1.1) as follows:  Treatment with garetosmab prevents the formation of new HO lesions in patients with FOP.  This hypothesis was tested primarily in the group of patients who crossed over from placebo to garetosmab in Period 2. Analysis in patients who were treated with garetosmab in both Periods 1 and 2 aimed to provide insight into the durability of pharmacological effect. | |
| 6. | No formal hypotheses were formulated for Period 3. The primary goals of Period 3 were to assess the persistence of efficacy and to assess the long-term safety of garetosmab. | |
| **Protocol amendment 6** | | |
| **Change** | | **Rationale for Change** |
| Added statements to address the impact of the COVID-19 pandemic:  That temporary, alternative mechanisms and flexibility in the visit schedule that may be used to ensure the continuity of clinical study conduct and oversight in light of the public health emergency related to COVID-19.  Defined the COVID-19 modified intent-to-treat principle that will be used for the week 56 primary analysis of efficacy endpoints related to imaging | | To explain the plan for ensuring continuity of clinical study activities and study oversight activities during the COVID-19 public health emergency.  To address the potential impact of the COVID-19 pandemic on the collection of imaging scans and/or missed or delayed study drug administration |
| Added a summary of the primary analysis results for efficacy and safety in Period 1 (week 28)  Added a summary of risks for epistaxis and skin and soft tissue infection | | To provide a high-level summary of the study results for efficacy and safety for Period 1(week 28) |
| Added a new hypothesis for Period 2 (week 56; open-label treatment period) based on the current understanding of the biology of FOP | | To update the planned efficacy analysis for Period 2 (week 56; open-label treatment period) based on the primary efficacy analysis results from Period 1 (week 28; double-blind, placebo- controlled period) |
| Added a rationale for the analyses to confirm the new hypothesis (week 56) with additional alpha of 0.1 specified for control for Type I multiplicity for Period 2 (week 56).  Revised the description of the Clinical Endpoint Measures | | To specify new analyses which can handle the possibility of patients with missing week 56 imaging assessment (due to COVID-19 disruption for the study) and are consistent with regulatory guidance for analysis for rare disease patient populations. |
| Added new endpoints for Period 2 (week 56) to confirm the understanding of the biology of new HO bone lesions:  primary efficacy endpoint  key secondary efficacy endpoints  other secondary efficacy endpoints | | To describe the added endpoints or analysis in Period 2 (open label treatment period; Week 56) |
| Added a new primary endpoint (week 56) based on low-dose CT assessment  Reorganized the Study Variables section to present endpoints by Period 1 (week 28), Period 2 (week 56), and Period 3 (week 76) | | To specify a primary endpoint using an imaging assessment that may be easier to perform than the PET assessment during the COVID-19 |
| Added description of the new statistical analyses for efficacy in Period 2 (week 56) | | To specify the new statistical analyses in Period 2 (open-label treatment period; week 56) based on the primary efficacy analysis results from Period 1 (week 28; double-blind, placebo-controlled period) |
| Updated the description of the analysis of efficacy endpoints in Period 1 (Week 28) was updated (amendment 6) to match the final SAP for Period 1 | | To be consistent with the final SAP for Period 1 (double-blind treatment period; week 28) issued before the database lock |
| Clarified the description of drug (functional REGN2477) and target (total activin A) assessed | | To be consistent with the drug and target assays |
| Added a description of the criteria for continued access to REGN2477 for patients who completed Period 3 (week 76) | | To be consistent with our corporate policy governing access to investigational drugs in confirmatory clinical studies. |
| Removed the requirement for a safety discussion with investigators prior to each administration of study drug after week 56. Therefore, after week 56 the need for such interactions will be decided on a case by case basis. | | The safety results for Period 1 (week 28) have not identified significant safety concerns that require continued safety discussions or frequent safety laboratory assessments. |
| Reduced the frequency of blood sample collection to every 3 months after week 56 for assessments of safety and ADA | | The reduction in assessments for safety and ADA after week 56 reduces the burden on patient. |
| Added assessments for FOP I-ADL, EQ-5D-3L, CAJIS and spirometry and collection of blood samples for measurement of drug and target, and ADA every 24 weeks after week 76. | | To support assessment of safety, PK, target, and ADA. |
| Will ask patients who permanently discontinue study treatment but do not withdraw from the study to return to the clinic for blood sample collection for   - measurement of serum drug and target concentration, at 4 weeks, 8 weeks, and 16 weeks, and 28 weeks after the patient's last dose. - assessment of ADA at the patients last visit. | | The request will support assessment of   - drug elimination phase - safety assessments for follow-up of patients - assessment for potential disease rebound - assessment of ADA at the end of the study |
| Updated Pharmacovigilance and Risk Management to Global Patient Safety in various sections | | Text change reflects the new organization name |
| **Protocol amendment 5** | | |
| **Change** | | **Rationale for change** |
| The option for subcutaneous (SC) administration is removed as the concentration of the current formulation is not suitable for SC use. | | The purpose of this amendment is to revise the description of study treatment based on a new liquid formulation. |
| **Protocol amendment 4** | | |
| **Change** | | **Rationale for change** |
| The order of statistical testing of the primary efficacy and key secondary endpoints is: Baseline-Active heterotopic ossification (AHO) analysis set first and Baseline-Active HO Classic ACVR1[R206H] Mutation (AHOC) analysis set second. | | This is a non-substantial amendment to change the hierarchy order of statistical testing for the primary efficacy and key secondary endpoints in response to comments from a health authority. |
| **Protocol amendment 3** | | |
| **Change** | | **Rationale for Change** |
| In order to mitigate the potential risk for epistaxis, 3 exclusion criteria were added:   - #18 Patients who are on concomitant antiplatelet therapy (eg, clopidogrel), anti‑coagulants (eg, warfarin, heparin, factor Xa inhibitor, or thrombin inhibitors) in the last 30 days or within 5 half-lives of the therapy, whichever is longer. Low dose acetylsalicylic acid (aspirin) is acceptable. - #19 Patients with a history of severe, non-traumatic bleeding requiring transfusion or hospitalization for hemodynamic compromise - #20 Patients with a known pre-existing medical history of a bleeding diathesis (eg, hemophilia A, von Willebrand’s Factor deficiency, platelet count ≤20x10^9^/L).   As an additional mitigation strategy, investigators should discourage the use of anti‑inflammatory drugs that inhibit platelet function or are otherwise associated with an increased risk of bleeding, to the extent possible.  Clarified the low dose acetylsalicylic acid (aspirin; [≤100 mg/day]) | | The purpose of this amendment is to revise the protocol to reflect new safety information relating to a potential risk for epistaxis. |
| Added baseline exploratory research sample to explore mechanism of epistaxis and REGN2477 mechanism of action  Added antiplatelet therapy, anticoagulants, and herbal supplements (that inhibit platelet function or are associated with increased risk of bleeding) to the list of prohibited medications  To increase early detection of epistaxis or bleeding events, the following were added as adverse events of special interest (AESIs):   - Moderate to severe episodes of non-traumatic bleeding - Moderate epistaxis (defined as any episode lasting longer than 30 minutes or requiring professional medical intervention) - Severe epistaxis (based on definition of a severe AE as per protocol) | | To aid in the exclusion of patients who may have existing propensity for bleeding and to assess the effect of REGN2477 administration, baseline and post-treatment laboratory measures of coagulation parameters and platelet effector function were added. |
| **Protocol amendment 2** | | |
| **Change** | | **Rationale for Change** |
| To support interpretation of the study results, all patients will have their ACVR1 gene sequenced during the study (mandatory).  The primary statistical analyses for the study will be based on the original study design, not the expanded study population. | | The purpose of this amendment is to remove the requirement that patients have the specific classic ACVR1[R206H] mutation. This change is based on in vitro studies which demonstrated that different mutations in ACVR1 receptors transduced bone morphogenic protein signaling when stimulated with Activin A. These results indicate that REGN2477 may be effective for all FOP mutations. This change expands the scope of the study to include patients with a clinical diagnosis of FOP who may have different ACVR1 mutations. |
| Use a hierarchical testing procedure instead of the Hochberg procedure. Revised Statistical Plan  Specify hierarchical testing of the 4 efficacy primary endpoints  Specify that the baseline active HO analysis set includes all randomized patients who had active HO lesion at baseline  Specify that patients will be randomized to REGN2477 or placebo in a 1:1 ratio by ACVR1 mutation (classic [R206H], other) in addition to gender and presence/absence of baseline active HO lesions.  Specify that results are analyzed first in patients with the ACVR1[R206H] mutation and secondly in patients with any ACVR1 mutation.  Make minor corrections to descriptions and tables | | The statistical analysis was revised to address feedback from regulatory agencies, as well as addressing the changes related to inclusion of patients with ACVR1 mutations. |
| The duration of time for patients to maintain highly effective contraception was extended to 30 weeks after the last dose of study drug to address feedback from a regulatory agency.  Added an exclusion criterion “Previous history or diagnosis of cancer” and updated the SAE definition to note that “a new diagnosis or progression of a malignancy in patients enrolled in the study will also be considered a SAE” to address feedback from a regulatory agency.  Deleted ‘futility’ as a reason for premature termination of the study  Added individual level dose modification criteria, clarify the study level dose modification, and specify that a pregnancy will be tracked until delivery along with a 3-month postnatal follow-up period for the infant, in response to a health authority request. | | To address feedback from a regulatory agency: |
| Added the week 76 time point to the additional analyses of safety and efficacy data to be performed following the open-label treatment period for consistency with the planned analyses. | |  |
| Added a secondary objective and secondary endpoints for assessment of the effect of REGN2477, between week 28 and week 56, on the number, activity, and volume of HO lesions identified by ^18^F-NaF PET or by CT in patients who switch from placebo to REGN2477 at week 28. | |  |
| Added assessment of hs-CRP as a biomarker  Added assessment of magnesium to the clinical chemistry panel  Added an endpoint for FEV1 of spirometry  Added measurement of height at baseline | |  |
| Provided more detail about the frequency of IDMC meetings and clarified the monitoring for potential AEs in male patients. | |  |
| Clarified the rationale for use of SC route of administration.  Provided details on how the safety of each dose will be reviewed prior to next dose. | |  |
| Clarified the PET/CT procedures and the exploratory endpoints assessed by PET/CT. | |  |
| Clarified that treatment with imatinib or isotretinoin is also exclusionary and are prohibited medications | |  |
| Added or updated footnotes to indicate:  that patients can be rescreened for study participation  that measurement of a patient’s height may not be precise or not possible to be assessed  clarifying the urinalysis procedure | | Provide more detail about the clinical endpoint measures and study assessments |
| Removed futility as a reason for study termination | |  |
| Minor edits for clarification of study terminology and correction:   - Clarified the description of the sample size   Correct an error in description of the full analysis set in the synopsis | |  |
| Minor edits for clarification of study terminology, corrections, and minor changes:  Period 1, Period 2, and Period 3 instead of “treatment period 1”, “treatment period 2”, or “treatment period 3”  “diary” has been replaced with “e-diary”.  “FOP disease diary” has been replaced with “FOP disease activity diary”.  “Menstrual History” instead of “Menstrual Questionnaire”  “FOP Independent Activities of Daily Living” instead of “Activity of Daily Living”  “FCV” to “FVC”  Subjects changed to patients  Adding “Doppler” to scrotum ultrasound | |  |
| **Protocol amendment 1** | | |
| **Change** | | **Rationale for Change** |
| Changed “the 24-week follow-up period” to the “follow-up treatment period” (Period 3). Each patient will continue to receive REGN2477 every 4 weeks during this period until they complete the week 76 visit, and all data have been collected and validated through the time when the last patient randomized into the study completes the week 28 visit (Treatment Period 1), and results of the primary analyses of safety and efficacy are available to the sponsor. If at the time patients reach the week 76 visit data from the last patient randomized into the study through the week 28 visit have not been collected and validated, and results of the primary analyses of safety and efficacy are not yet available to the sponsor, patients will continue to receive REGN2477 every 4 weeks, beyond week 76.  Removed footnote #17 from Schedule of Events, Table 3, as this footnote addressed the transition from on-treatment to off- treatment which is no longer relevant to the study.  The deletion of this footnote caused footnote #19 for Daily Pain NRS and Daily FOP disease activity diary from Schedule of Events, Table 1 to be changed to footnote#18.  Removed the post-treatment period as the third observation period, because treatment with REGN2477 will continue during Treatment Follow-up (Period 3) through the end of the study. | | Treatment with REGN2477 will continue provided that no safety signals are identified during continuous monitoring of the study by the investigator, medical monitor and IDMC. In addition, all patients will be required to maintain contraception during this period. |
| “End of Treatment” was removed from visit 17 of Period 2 (Open Label Treatment Period), as the end of study will occur after Period 3 (Follow-up Treatment Period). | |  |
| Schedule of Events Table 3: Addition of a new column entitled: “End of Study/Week 80” to denote the new end of study and to indicate the procedures that will be conducted during this period in the event that data from the last patient randomized into the study through the week 28 visit (Treatment Period 1) have not been collected and validated, and results of the primary analyses of safety and efficacy are not yet available to the sponsor.  A new row entitled “Administer REGN2477” was added. | | Reduced visit windows in Period 3 from ±14 days to ±7 days, as patients will continue to be treated with REGN2477 during this period. |
| Removed the sentence regarding patients entering the off-drug treatment phase after early termination as the off-drug phase was removed and the statement is no longer applicable | |  |
| Clarified that PET/CT scans will be read and analyzed using a blinded central reading center during the study, however at baseline, added the provision that the investigator will be allowed to independently read and analyze PET/CT images. Baseline PET/CT images will be performed before each patient is randomized into the study. As these baseline images do not contain any unblinding data, the PI may utilize these images as part of standard of care resulting in not having to duplicate the imaging assessments and thereby reducing the patients’ total annual radiation exposure. | |  |
| Added the requirement that any incidental findings identified through PET/CT scans will be reported to the investigator and medical monitor, and that the investigator will communicate any findings to the patient.  Added the requirement that any incidental findings identified through the optional DNA analysis will be reported to the investigator and/or the VuMC Unsolicited Finding Committee (UFC) for appropriate follow-up with the patient. Also clarified that by signing the informed consent for the optional DNA sub-study, the patient agrees to be informed of any incidental findings. | |  |
| Added whole body ^18^F-NaF PET and whole body low dose CT imaging at week 76 (day 533 [± 7 days]) for a total of 5 scans during the study. This will allow for the evaluation of longer term effects of REGN2477 (ie, an additional 20 weeks of treatment) in FOP patients. As a result of the additional scan, the total ED range approximation was changed from 21 mSv to 25 mSv. | |  |
| Added that ECG and symptom-directed physical examination may be performed during the follow-up treatment period (Period 3, after week 56) with REGN2477 at the investigator’s discretion, and that urine pregnancy testing will be performed prior to each REGN2477 dose administration, and a serum pregnancy test will be performed at the patient’s end of study visit. PK/ADA sampling will continue to be performed at all visits after week 56. | |  |
| Removed the post-treatment period from the definitions of Adverse Events, as the post-treatment period is no longer relevant. | |  |
| Added the week 76 time point to the additional analyses of safety and efficacy data to be performed following the open-label treatment period for consistency with the planned analyses in the global study. | |  |

#### Table S2. Key inclusion/exclusion criteria

**Key Inclusion Criteria**

Men and women 18–60 years of age at screening

Clinical diagnosis of FOP (based on findings of congenital malformation of the great toes, episodic soft tissue swelling, and/or progressive HO)

Confirmation of FOP diagnosis with documentation of any *ACVR1* mutation

FOP disease activity within 1 year of screening visit. FOP disease activity is defined as pain, swelling, stiffness, and other signs/symptoms associated with FOP flare-ups; or worsening of joint function, or radiographic progression of HO (increase in site or number of HO lesions) with/without being associated with flare-up episodes

Willing and able to undergo PET and CT imaging procedures and other procedures as defined in this study

**Key Exclusion Criteria**

Significant concomitant illness or history of significant illness such as, but not limited to cardiac, renal, rheumatologic, neurologic, psychiatric, endocrine, metabolic, or lymphatic disease,

Previous history or diagnosis of cancer

Use of bisphosphonate within 1 year of screening

Concurrent participation in another interventional clinical study, or a non-interventional study with radiographic measures or invasive procedures (eg, collection of blood or tissue samples). Participation in the FOP Connection Registry or other studies in which participants complete study questionnaires are allowed

Treatment with another investigational drug, denosumab, imatinib, or isotretinoin in the last 30 days or within 5 half-lives of the investigational drug, whichever is longer

Pregnant or breastfeeding women

Male and women of childbearing potential participants who are unwilling to practice highly effective contraception

*Patients who are on concomitant antiplatelet therapy or anti-coagulants in the last 30 days or within 5 half-lives of therapy (low-dose acetylsalicylic acid is acceptable).

*Patients with a history of severe, non-traumatic bleeding requiring transfusion or hospitalization for hemodynamic compromise

*Patients with a known pre-existing medical history of a bleeding diathesis

*Additional exclusion criteria were added to the protocol through an amendment, based upon the occurrence of one severe epistaxis event, and to mitigate the potential risk for epistaxis. Half of the patients were already included when the protocol was amended. CT=computed tomography; FOP= fibrodysplasia ossificans progressiva; HO=heterotopic ossification; PET=positron emission tomography.
Table S3. List of Key Primary and Secondary Outcomes for Period_1, Period_2 and open label extension (Per Protocol)

**Period_1**

**Primary Safety Endpoint: Period_1**

Incidence and severity of TEAEs through the end of the Period 1 [time frame: Baseline to week 28]

**Primary Outcome Measures: Period_1**

Time-weighted average (standardized AUC) of the percent change in TLA by ^18^F-NaF PET using AHO [time frame: Baseline to week 28]

Percent change in the total volume of HO lesions as assessed by CT using AHO [time frame: Baseline to week 28]

Time-weighted average (standardized AUC) of the percent change in TLA by ^18^F-NaF PET using AHOC [time frame: Baseline to week 28]

Percent change in the total volume of HO lesions as assessed by CT using AHOC [time frame: Baseline to week 28]

**Key Secondary Outcome Measures: Period_1**

Time-weighted average (standardized AUC) of the change in daily pain due to FOP, as measured using the daily NRS using AHO [time frame: Baseline to week 28]

Time-weighted average (standardized AUC) of the change in daily pain due to FOP, as measured using the daily NRS using AHOC [time frame: Baseline to week 28]

**Other Secondary Outcome Measures: Period_1**

Percent change in ^18^F-NaF SUVmax of individual active HO site(s) by PET using AHOC [time frame: Baseline to week 8]

Percent change in ^18^F-NaF SUVmax of individual active HO site(s) by PET using AHO [time frame: Baseline to week 8]

Change in number of HO lesions as assessed by ^18^F-NaF PET using AHOC [time frame: Baseline to week 28]

Change in number of HO lesions as assessed by ^18^F-NaF PET using AHO [time frame: Baseline to week 28]

Change in number of HO lesions as assessed by ^18^F-NaF PET using FAS [time frame: Baseline to week 28]

Change in number of HO lesions detectable by CT using AHOC [time frame: Baseline to week 28]

Change in number of HO lesions detectable by CT using AHO [time frame: Baseline to week 28]

Change in number of HO lesions detectable by CT using FAS [time frame: Baseline to week 28]

Time-weighted average (standardized AUC) of the change from baseline in daily pain due to FOP, as measured using the daily NRS using FAS [time frame: Baseline to week 28]

Time-weighted average (standardized AUC) of the percent change from baseline in biomarkers of bone formation levels (including Total Procollagen Type 1 N-Terminal Propeptide, bone specific alkaline phosphatase, and total alkaline phosphatase) in serum over 28 weeks using FAS [time frame: up to week 28]

**Exploratory Outcome Measures: Period_1**

Change from baseline in CAJIS total score to week 28 using AHO [time frame: Baseline to week 28]

Change from baseline in EQ-5D-3L total score to week 28 using AHO [time frame: Baseline to week 28]

Patient-reported assessment of activities of daily living by FOP I-ADL [time frame: Baseline to week 28]

Number and duration of flare-ups by patient e-diary (defined as experiencing ≥2 of the following: new onset of pain, swelling, joint stiffness, decrease in movement, or detection of HO) [time frame: Baseline to week 28]

Location and severity of FOP disease signs and symptoms by patient e-diary including 1) pain, 2) swelling, 3) joint stiffness, 4) decreased movement, each scored using a 4-point scale (0: no symptom; 1: mild; 2: moderate; 3: severe) [time frame: Baseline to week 28]

Total dosage of glucocorticoids use over time

Number of patients with new HO lesions as assessed by ^18^F-NaF PET using AHO [time frame: Baseline to week 28]

Change in number of lesions that are only ^18^F-NaF PET detectable at baseline to CT detectable lesions at week 28 [time frame: Baseline to week 28]

Change from baseline in the FEV1 of spirometry at week 28 [time frame: Baseline to week 28]

Time-weighted average (standardized AUC) change from baseline in the levels of hs-CRP [time frame: Baseline to week 28]

**Period_2**

**Primary Safety Endpoint: Period_2**

Incidence and severity of TEAEs through the end of the Period 2 [time frame: Baseline to week 56]

**Primary Outcome Measures: Period_2**

Number of new HO lesions as assessed by CT at week 56 relative to week 28 using AHO (in patients switching from placebo to garetosmab after Period 1) [time frame: week 28 to week 56]

**Key Secondary Outcome Measures: Period_2**

Total volume of new HO lesions in patients switching from placebo to garetosmab after Period 1 as assessed by CT using AHO [time frame: week 28 to week 56]

Change in number of HO lesions by ^18^F-NaF PET versus the same patients between baseline and week 28 using AHO [time frame: week 28 to week 56]

TLA by ^18^F-NaF PET in new HO lesions in patients switching from placebo to garetosmab after Period 1 using AHO [time frame: week 28 to week 56]

Percentage of patients switching from placebo to garetosmab after Period 1 with new HO lesions as assessed by CT using AHO [time frame: week 28 to week 56]

Percentage of patients switching from placebo to garetosmab after Period 1 with new HO lesions as assessed by ^18^F-NaF PET using AHO [time frame: week 28 to week 56]

**Other Secondary Outcome Measures: Period_2**

Number of new HO lesions in patients switching from placebo to garetosmab after Period 1 as assessed by CT using AHO [time frame: week 28 to week 56]

Total volume of new HO lesions as assessed by CT in patients switching from placebo to garetosmab after Period_1 using AHO [time frame: week 28 to week 56]

Percent of patients with new HO lesions as assessed by CT in patients switching from placebo to garetosmab after Period_1 using AHO [time frame: week 28 to week 56]

Percentage of patients switching from placebo to garetosmab after Period 1 with investigator-assessed flare-ups using AHO [time frame: week 28 to week 56]

Percentage of patients switching from placebo to garetosmab after Period 1 with flare-ups assessed by patient e-diary using AHO [time frame: week 28 to week 56]

Number of new HO lesions in patients who continue garetosmab after Period 1 as assessed by CT using AHO [time frame: Baseline to week 56]

Number of new HO lesions as assessed by CT in patients who continue garetosmab after Period 1 using AHO [time frame: baseline to week 56]

Number of new HO lesions as assessed by CT in patients who continue garetosmab after Period 1 using AHO [time frame: week 28 to week 56]

Total volume in new HO lesions as assessed by CT in patients who continue garetosmab after Period 1 using AHO [time frame: Baseline to week 56]

TLA in new HO lesions as assessed by ^18^F-NaF PET in patients who continue garetosmab after Period 1 using AHO [time frame: Baseline to week 56]

Percent of patients with new HO lesions as assessed by ^18^F-NaF PET in patients who continue garetosmab after Period 1 using AHO [time frame: baseline to week 56]

Percent change from week 28 in SUVmax to week 56 in patients switching from placebo to garetosmab after Period 1 using AHO [time frame: week 28 to week 56]

Percent change from baseline in SUVmax to week 56 in patients who continue garetosmab after Period 1 using AHO [time frame: Baseline to week 56]

Percent change from baseline in TLA by ^18^F-NaF PET using AHO [time frame: Baseline to week 56]

Percent change in the total volume of HO lesions in patients switching from placebo to garetosmab after Period 1 versus the same patients between baseline and week 28 as assessed by CT using AHO [time frame: Baseline to week 56]

Percent change from baseline in the total volume of HO lesions as assessed by CT using AHO [time frame: Baseline to week 56]

Change in number of HO lesions in patients switching from placebo to garetosmab after Period 1 as assessed by ^18^F-NaF PET from week 28 to week 56 versus the same patients between baseline and week 28 using AHO [time frame: Baseline to week 28; week 28 to week 56]

Change in number of HO lesions in patients switching from placebo to garetosmab after Period 1 as assessed by CT scan from week 28 to week 56 versus the same patients between baseline and week 28 using AHO [time frame: Baseline to week 28; week 28 to week 56]

**Exploratory Outcome Measures: Period_2**

Change in the total volume of new HO lesions as assessed by CT in patients switching from placebo to garetosmab after Period 1 using AHO [time frame: week 28 to week 56]

Joint function assessment by physician at week 56 by the CAJIS using AHO [time frame: week 28 to week 56]

Patient-reported quality of life by EQ-5D-3L at week 56 using AHO [time frame: week 28 to week 56]

Patient-reported assessment of activities of daily living by FOP I-ADL at week 56 using AHO [time frame: week 28 to week 56]

Number and duration of flare-ups by patient e-diary (defined as experiencing ≥2 of the following: new onset of pain, swelling, joint stiffness, decrease in movement, or detection of HO) [time frame: week 28 to week 56]

Location and severity of FOP disease signs and symptoms by patient e-diary including 1) pain, 2) swelling, 3) joint stiffness, 4) decreased movement, each scored using a 4-point scale (0: no symptom; 1: mild; 2: moderate; 3: severe) [time frame: week 28 to week 56]

Number of new HO lesions as assessed by ^18^F-NaF PET in patients who continue garetosmab after Period 1 using AHO [time frame: baseline to week 56]

Change in number of HO lesions by CT using AHO [time frame: baseline to week 56]

Number of patients with new HO lesions as assessed by ^18^F-NaF PET using AHO [time frame: baseline to week 56]

Percent change from baseline of mean SUVmean of selected normal bones (e.g., lumbar spine and femoral heads) as assessed by ^18^F-NaF PET/CT using AHO [time frame: baseline to week 56]

Percent change from baseline in venous plasma clearance ^18^F-NaF SUV using AHO [time frame: baseline to week 56]

Percent change from baseline in ^18^F-NaF incorporation rate (Ki) in individual active HO lesion(s) using AHO [time frame: baseline to week 56]

Percent change from baseline of ratio of SUVmax of individual active HO lesion(s) to venous plasma SUV during PET scan using AHO [time frame: baseline to week 56]

Change from baseline in the FEV1 of spirometry using AHO [time frame: baseline to week 56]

Change from baseline in the FEV1 of spirometry using AHO [time frame: baseline to week 28]

Endpoints were ordered per protocol. Period_2 endpoints- new hypothesis were refined based on period 1 ^18^F-NaF PET=fluorine-18-labelled sodium fluoride positron emission tomography; ADA=anti-drug antibody; AHO=active heterotopic ossification analysis set; AHOC=active heterotopic ossification classic mutation analysis set; AUC=area under the curve; CAJIS=Cumulative Analog Joint Involvement Scale; CT=computed tomography; EQ-5D-3L=EuroQol 5 dimensions questionnaire with a 3-level scale; FAS=full analysis set; FOP=fibrodysplasia ossificans progressiva; HO=heterotopic ossification; NRS=numeric rating scale; PK=pharmacokinetic; SUVmax=maximum standardized uptake value; TEAE=treatment-emergent adverse event; TLA=total lesion activity.

#### Table S4. Baseline demographics and disease characteristics*

|  | **Placebo (n=24)** | **Garetosmab 10 mg/kg Q4W (n=20)** | **Total (N=44)** |
| --- | --- | --- | --- |
| **Demographics** |  |  |  |
| Age, years, mean (SD) | 27.8 (8.5) | 27.3 (8.7) | 27.6 (8.5) |
| Ethnicity, n (%) |  |  |  |
| Not Hispanic or Latino | 24 (100) | 20 (100) | 44 (100) |
| Race, n (%) |  |  |  |
| White | 22 (91.7) | 17 (85.0) | 39 (88.6) |
| Black or African American | 1 (4.2) | 0 | 1 (2.3) |
| Asian | 1 (4.2) | 2 (10.0) | 3 (6.8) |
| Other | 0 | 1 (5.0) | 1 (2.3) |
| Sex, n (%) |  |  |  |
| Female | 14 (58.3) | 11 (55.0) | 25 (56.8) |
| Weight, kg, mean (SD) | 64.0 (23.4) | 57.4 (10.1) | 61.0 (18.7) |
| Region, n (%) |  |  |  |
| North America | 6 (25.0) | 4 (20.0) | 10 (22.7) |
| Europe | 18 (75.0) | 16 (80.0) | 34 (77.3) |
| **Clinical Characteristics** |  |  |  |
| Age at FOP diagnosis, years, mean (SD) | 8.1 (7.5) | 9.1 (5.5) | 8.5 (6.6) |
| Duration of FOP disease, years, mean (SD) | 19.9 (10.0) | 18.3 (11.0) | 19.2 (10.4) |
| FOP genetic mutation, n (%) |  |  |  |
| R206H (classic) | 22 (91.7) | 20 (100) | 42 (95.5) |
| Other | 2 (8.3) | 0 | 2 (4.5) |
| FEV1, L, mean (SD)† | 1.9 (0.8) | 1.8 (0.6) | 1.9 (0.7) |
| Percent predicted FEV1, %, mean (SD)† | 53.2 (19.1) | 53.0 (15.8) | 53.1 (17.4) |
| FVC, L, mean (SD)† | 2.1 (0.9) | 2.0 (0.7) | 2.1 (0.8) |
| P1NP, ug/L, mean (SD) | 138.1 (100.4) | 182.9 (168.5) | 158.5 (135.8) |
| BSAP, U/L, mean (SD) | 32.8 (14.8) | 32.4 (15.9) | 32.7 (15.1) |
| tAP, IU/L, mean (SD) | 76.8 (25.0) | 72.9 (29.0) | 75.0 (26.7) |
| **Patient- and Physician-Reported Outcomes** |  |  |  |
| NRS, mean (SD)ǂ |  |  |  |
| Average daily pain | 1.96 (2.175) | 2.04 (2.071) | 1.99 (2.104) |
| Average daily pain over 7 days | 2.17 (2.246) | 2.01 (1.805) | 2.10 (2.041) |
| Patients with flare-ups in previous 12 months, n (%)§ | 20 (83.3) | 19 (95.0) | 39 (88.6) |
| Total Joint Function Score (CAJIS), mean (SD) ǁ | 15.7 (6.2) | 15.8 (7.3) | 15.7 (6.6) |
| EQ-5D-3L Total Score, mean (SD) | 9.4 (1.5) | 9.2 (2.1) | 9.3 (1.8) |
| FOP I-ADL, mean (SD) | 69.3 (26.3) | 77.9 (35.9) | 73.2 (30.9) |
| **Imaging Characteristics** |  |  |  |
| Presence of active HO at baseline, n (%)¶ | 24 (100) | 20 (100) | 44 (100) |
| Number of active HO lesions by ^18^F-NaF PET, n (%) |  |  |  |
| 1 | 0 | 0 | 0 |
| 2 | 3 (12.5) | 1 (5.0) | 4 (9.1) |
| 3 | 1 (4.2) | 1 (5.0) | 2 (4.5) |
| 4 | 2 (8.3) | 2 (10.0) | 4 (9.1) |
| 5 | 1 (4.2) | 2 (10.0) | 3 (6.8) |
| 6 | 4 (16.7) | 1 (5.0) | 5 (11.4) |
| 7 | 13 (54.2) | 13 (65.0) | 26 (59.1) |
| Total lesion activity by ^18^F-NaF PET, mean (SD) | 473.4 (348.4) | 418.2 (372.8) | 448.3 (356.5) |
| Number of HO lesions by CT, n (%) |  |  |  |
| 1 | 0 | 0 | 0 |
| 2 | 3 (12.5) | 1 (5.0) | 4 (9.1) |
| 3 | 2 (8.3) | 2 (10.0) | 4 (9.1) |
| 4 | 1 (4.2) | 1 (5.0) | 2 (4.5) |
| 5 | 1 (4.2) | 2 (10.0) | 3 (6.8) |
| 6 | 6 (25.0) | 1 (5.0) | 7 (15.9) |
| 7 | 11 (45.8) | 13 (65.0) | 24 (54.5) |
| Total volume of HO lesions by CT, cm^3^, mean (SD) | 235.8 (253.3) | 251.4 (327.9) | 242.9 (286.2) |

^18^F-NaF PET=fluorine-18-labelled sodium fluoride positron emission tomography; AHO=baseline active HO analysis set; BSAP=bone specific alkaline phosphatase; CAJIAS=Cumulative Analog Joint Involvement Scale; CT=computed tomography; EQ-5D-3L=EuroQol 5 dimensions questionnaire with a 3-level scale; FEV1=forced expiratory volume in 1 second; FOP=fibrodysplasia ossificans progressiva; FVC=forced vital capacity; HO=heterotopic ossification; I-ADL=Instrumental Activities of Daily Living; L=liter; NRS=Numeric Rating Scale; P1NP=procollagen type 1 N-terminal propeptide; Q4W=every 4 weeks; ROI=region of interest; SD=standard deviation; SUVmax=maximal standardized uptake value; SUVmean=mean standardized uptake value; tAP=total alkaline phosphatase. *AHO. †N’s for spirometry are as follows: n=22 (placebo), n=19 (garetosmab 10 mg/kg Q4W), and n=41 (total). ǂN’s for NRS are as follows: n=23 (placebo), n=18 (garetosmab 10 mg/kg Q4W), and n=41 (total). § Patient e-diary. ǁPhysician assessment. ¶Defined as ≥1 lesion with a SUVmax that is ≥3 times the SUVmean for the supra-acetabular ROI.

#### Table S5. Summary of primary, and secondary endpoints for Period_1 and Period_2

**A. Period_1 Results**

| **Objectives**  **multiplicity controlled** | **Endpoint (multiplicity controlled)*** | **Population** | **Model**^†,‡^ | **Garetosmab**  **LSM (SE)** | **Placebo**  **LSM (SE)** | **LSM difference**  **(95% CI)** | **p-value** |
| --- | --- | --- | --- | --- | --- | --- | --- |
| Primary- PET | Time-weighted average of percent change from baseline in TLA over 28 weeks | AHO | ANCOVA | -8.1 (9.93) | 16.6 (9.11) | -24.6 (-51.8, 2.5) | 0.0741 |
| Primary-CT | Percent change from Baseline in total volume of HO lesions at Week 28 | AHO | MMRM | 7.1 (20.43) | 32.0 (18.66) | -24.9 (-80.8, 30.9) | 0.3726 |
| Primary-PET | Time-weighted average of percent change from baseline in TLA over 28 weeks | AHOC | ANCOVA | -8.0 (10.14) | 17.6 (9.73) | -25.6 (-53.9, 2.8) | 0.0756 |
| Primary-CT | Percent change from baseline in total volume of HO lesions at Week 28 | AHOC | MMRM | 7.0 (20.87) | 34.9 (19.90) | -27.8 (-86.1, 30.5) | 0.3407 |
| Key Secondary – Pain NRS | Time-weighted average of change from baseline in daily average pain over 28 weeks | AHO | ANCOVA | -0.51 (0.231) | -0.17 (0.21) | -0.34 (-0.96, 0.27) | 0.2656 |
|  |  | AHOC | ANCOVA | -0.48 (0.237) | -0.12 (0.22) | -0.36 (-1.01, 0.29) | 0.2651 |
| **Secondary/exploratory objectives** | **Endpoint** | **Population** | **Model**^†,‡^ | **Garetosmab*** | **Placebo*** | **Difference (95% CI)** | **p-value** |
| Secondary – Imaging – SUVmax | Percent change from baseline in mean SUVmax at Week 8 | AHO | ANCOVA | -22.3 (4.02) | -6.6 (3.70) | -15.7 (-26.8, -4.6) | 0.0065 |
|  |  | AHOC | ANCOVA | -22.3 (3.69) | -5.3 (3.54) | -17.0 (-27.4, -6.7) | 0.0019 |
| Secondary – Imaging - # PET Lesion | Change from baseline in number of HO lesions by PET at Week 28 | AHO | Ranked ANCOVA | -2 (-7, 1) | -2 (-5, 8) | -1 (-2, 0) | 0.1768 |
| Secondary – Imaging - # PET Lesion | Change from baseline in number of HO lesions by PET at Week 28 | AHOC | Ranked ANCOVA | -2 (-7, 1) | -2 (-5, 8) | -1 (-2, 1) | 0.1746 |
| Secondary – Imaging - # CT Lesion | Change from baseline in number of HO lesions detectable by CT at Week 28 | AHO | Ranked ANCOVA | 0 (-4, 1) | 1 (0, 9) | -1 (-1, 0) | 0.0192 |
| Secondary – Imaging - # CT Lesion | Change from baseline in number of HO lesions detectable by CT at Week 28 | AHOC | Ranked ANCOVA | 0 (-4, 1) | 1 (0, 9) | -1 (-1, 0) | 0.0153 |
| Secondary – P1NP | Time-weighted average of percent change from baseline over 28 weeks | AHO | ANCOVA | -25.16 (7.062) | 1.01 (6.468) | -26.17 (-45.57, -6.76) | 0.0095 |
|  |  | AHOC | ANCOVA | -25.41 (7.023) | 3.43 (6.730) | -28.84 (-48.55, -9.13) | 0.0052 |
| Secondary – BSAP | Time-weighted average of percent change from baseline over 28 weeks | AHO | ANCOVA | -12.36 (4.853) | 9.76 (4.456) | -22.12 (-35.38, -8.86) | 0.0017 |
|  |  | AHOC | ANCOVA | -12.32 (4.873) | 11.08 (4.676) | -23.40 (-37.01, -9.80) | 0.0013 |
| Secondary – tAP | Time-weighted average of percent change from baseline over 28 weeks | AHO | ANCOVA | -1.77 (5.055) | 10.89 (4.649) | -12.66 (-26.51, 1.18) | 0.0719 |
| Secondary – tAP | Time-weighted average of percent change from baseline over 28 weeks | AHOC | ANCOVA | -1.77 (5.101) | 11.12 (4.901) | -12.89 (-27.15, 1.38) | 0.0753 |

^†^For ANCOVA or MMRM, LS mean (SE) and LS mean differences (95% CI) are shown. For Fisher’s exact test, number and proportion of patients shown.

^‡^For Ranked ANCOVA, median (mix, max) within group and median differences shown. For Ranked ANCOVA model, median treatment difference and 95% CI was derived from the Hodges-Lehmann estimate and Moses distribution-free CI, respectively. P-value was estimated from ANCOVA on ranked responses.

AHO, patients with at least 1 active heterotopic ossification lesion at baseline; AHOC, patients with at least 1 active heterotopic ossification lesion at baseline and with classic ACVR1[R206H] mutation; ANCOVA, analysis of covariance; BSAP, bone specific alkaline phosphatase; CAJIS, Cumulative Analog Joint Involvement Scale; CI, confidence interval; CRP, C-reactive protein; CT, computerized tomography; EQ5D, Health status questionnaire by EuroQol Group; HO, heterotopic ossification; IADL, independent activity daily living questionnaire; LSM, least-squares mean; MMRM, mixed model with repeated measures; NRS, numeric rating scale; PET, positron emission tomography; P1NP, procollagen type 1 N-Terminal propeptide; SE, standard error; tAP, total alkaline phosphatase; TLA, total lesion activity.

**B. Period_2 Results**

| Objectives  Multiplicity Controlled | Endpoint (Multiplicity Controlled) | Arm | Analysis Set | Model | Period 1 | Period 2 | Comparison | p-Value |
| --- | --- | --- | --- | --- | --- | --- | --- | --- |
| Primary - CT | Number of new HO lesions as assessed by CT at week 56 relative to week 28 scan | Placebo/garetosmab | COVID-19 mITT  (n=22) | Descriptive + Wilcoxon | Observed Rate = 1  Total # of lesions = 22 | Observed Rate = 0  Total # of lesions = 0 | Observed Rate Reduction = 100% | 0.0039 |
|  |  | Garetosmab/garetosmab | COVID-19 mITT  (n=18) | Descriptive | Observed Rate = 0.11  Total # of lesions = 2 | Observed Rate = 0  Total # of lesions = 0 |  |  |
| Key Secondary - CT | Total volume of new HO lesions as assessed by CT at week 56 relative to week 28 scan | Placebo/garetosmab | COVID-19 Mitt  (n=22) | MMRM+ Wilcoxon | LS Mean:  9.29 cm3 | LS Mean:  0.05 cm3 | LS Mean Difference:  -9.24 (-17.96, -0.52) | 0.0039 |
|  |  | Garetosmab/garetosmab | COVID-19 mITT  (n=18) | Descriptive | Mean Volume:  1 cm3 | Mean Volume:  0 cm3 |  |  |
| Key Secondary - PET | Number of new HO lesions as assessed by 18F-NaF PET at week 56 relative to week 28 scan | Placebo/garetosmab | COVID-19 mITT  (n=22) | GEE + Wilcoxon | Adjusted Rate:  0.93 (0.54, 1.62)  Total # of lesions = 23 | Adjusted Rate:  0.04 (0.01, 0.31)  Total # of lesions = 1 | Adjusted Rate Ratio:  0.05 (0.01, 0.33)  Rate Reduction= 95% | 0.0039 |
|  |  | Garetosmab/garetosmab | COVID-19 mITT  (n=18) | Descriptive | Observed Rate = 0.06 (Q1=0, Q2 = 0, Q3 = 0, max=1)  Total # of lesions = 1 | Observed Rate = 0  Total # of lesions = 1 |  |  |
| Key Secondary - PET | Total lesion activity by 18F-NaF PET in new lesions at week 56 relative to week 28 scan | Placebo/garetosmab | COVID-19 mITT  (n=22) | MMRM+ Wilcoxon | LS Mean:  204.45 | LS Mean:  13.20 | LS Mean Difference:  -191.25 (-390.80, 8.29) | 0.0273 |
|  |  | Garetosmab/garetosmab | COVID-19 mITT  (n=18) | Descriptive | Mean TLA:  4.7 | Mean TLA:  0 |  |  |
| Key Secondary - CT | Percent of patients with new HO lesions as assessed by CT at week 56 relative to week 28 | Placebo/garetosmab | COVID-19 mITT  (n=22) | Descriptive + McNemar | 40.9% (9/22) | 0 (0/22) | Observed Relative Risk Reduction = 100% | 0.0027 |
|  |  | Garetosmab/garetosmab | COVID-19 mITT  (n=18) | Descriptive | 11.1% (2/18) | 0 (0/18) |  |  |
| Key Secondary - PET | Percent of patients with new HO lesions as assessed by PET at week 56 relative to week 28 | Placebo/garetosmab | COVID-19 mITT  (n=22) | GEE + McNemar | 40.9% (9/22) | 4.5% (1/22) | Adjusted Relative Risk Reduction = 89%  Adjusted Odds Ratio:  0.07 (0.01, 0.48) | 0.0047 |
|  |  | Garetosmab/garetosmab | COVID-19 mITT  (n=18) | Descriptive | 5.6% (1/18) | 0 (0/18) |  |  |
| Secondary – Investigator Flare-ups | Percent of patients with new investigator-assessed flare-ups in Period 2 | Placebo/garetosmab | COVID-19 mITT  (n=22) | McNemar | 45.5% (10/22)  (Total # flare ups: 22) | 13.6% (3/22)  (Total # flare ups: 4) | Observed Relative Risk Reduction = 70% | 0.0196 |
|  |  | Garetosmab/garetosmab | COVID-19 mITT  (n=18) | Descriptive | 11.1% (2/18)  (Total # flare ups: 2) | 5.6% (1/18)  (Total # flare ups: 1) | ------------------- |  |
| Secondary – Patient Reported Flare-ups | Percent of patients with new flare-ups (using patient diary) in Period 2 | Placebo/garetosmab | COVID-19 mITT (n=22) | McNemar | 68.2% (15/22)  (Total # flare ups: 31) | 13.6% (3/22)  (Total # flare ups: 11) | Observed Relative Risk Reduction = 80% | 0.0005 |
|  |  | Garetosmab/garetosmab | COVID-19 mITT  (n=18) | Descriptive | 33.3% (6/18)  (Total # flare ups: 12) | 22.2% (4/18)  (Total # flare ups: 6) | ------------------- |  |
| Secondary-Imaging-CT-New Relative to Week 28 | Number of new HO lesions as assessed by CT Only at week 56 relative to week 28 scan | Placebo/garetosmab | COVID-19 mITT  (n=22) | Descriptive + Wilcoxon | Observed Rate = 1.27  Total # of lesions = 28 | Observed Rate = 0.13  Total # of lesions = 3 | ------------------- |  |
| Secondary-Imaging-CT-New Relative to Baseline | Number of new HO lesions as assessed by CT at week 56 relative to baseline | Garetosmab/garetosmab | COVID-19 mITT | Descriptive  (Bootstrap CI) | Mean:  0.1 (0.0, 0.3) | Mean:  0.1 (0.0, 0.3) | ------------------- |  |
| Secondary-Imaging-CT-New Relative to Baseline | Total volume in new HO lesions as assessed by CT at week 56 relative to baseline | Garetosmab/garetosmab | COVID-19 mITT | Descriptive  (Bootstrap CI) | Mean:  1.0 (0.0, 2.9) | Mean:  0.4 (0.0, 1.1) | ------------------- |  |
| Secondary-Imaging-CT-New Relative to Baseline | Percent of patients with new HO lesions as assessed by CT at week 56 relative to baseline | Garetosmab/garetosmab | COVID-19 mITT | Descriptive | 11.1% (2/18) | 11.1% (2/18) | ------------------- |  |
| Secondary-Imaging-PET-New Relative to Baseline | Number of new lesions as assessed by 18F-NaF PET at week 56 relative to baseline | Garetosmab/garetosmab | COVID-19 mITT | Descriptive  (Bootstrap CI) | Mean:  0.1 (0.0, 0.2) | Mean:  0.1 (0.0, 0.2) | ------------------- |  |
| Secondary-Imaging-PET-New Relative to Baseline | Total lesion activity in new HO lesions as assessed by 18F-NaF PET at week 56 relative to baseline | Garetosmab/garetosmab | COVID-19 mITT | Descriptive  (Bootstrap CI) | 4.7 (0.0, 12.7) | 0.7 (0.0, 1.8) | ------------------- |  |
| Secondary-Imaging-PET-New Relative to Baseline | Percent of patients with new HO lesions as assessed by 18F-NaF PET at week 56 relative to baseline | Garetosmab/garetosmab | COVID-19 mITT | Descriptive | 5.6% (1/18) | 5.6% (1/18) | ------------------- |  |
| Secondary – PET - SUVmax | Percent change from week 28 in SUVmax to week 56 | Placebo/garetosmab | COVID-19 mITT | Bootstrap + Wilcoxon | -16.8%  (-24.6%, -8.7%) | -30.3%  (-36.6%, -24.1%) | -13.8%  (-24.6%, -2.6%) |  |
|  |  | Garetosmab/garetosmab | COVID-19 mITT | Bootstrap | -32.1%  (-41.7%, -22.9%) | -16.6%  (-29.0, -6.5%) | 15.8%  (3.0%, 27.5%) |  |
| Secondary – PET - SUVmax | Percent change from baseline in SUVmax to week 56 | Garetosmab/garetosmab | COVID-19 mITT | Descriptive | -32.1%  (-41.7%, -22.9%) | -41.9%  (-55.1%, -28.8%) | ------------------- |  |
| Secondary – PET - TLA | Percent change from week 28 in total lesion activity by 18F-NaF PET to week 56 | Placebo/garetosmab | COVID-19 mITT | Bootstrap + Wilcoxon | 48.2%  (0.4%, 120.2%) | -16.4%  (-30.6%, -2.5%) | -66.9%  (-154.6%, -6.4%) |  |
|  |  | Garetosmab/garetosmab | COVID-19 mITT | Bootstrap | -15.9%  (-34.5%, 3.5%) | -3.6%  (-16.8%, 7.9%) | 14.0%  (-4.1%, 33.7%) |  |
| Secondary – PET - TLA | Percent change from baseline in total lesion activity by 18F-NaF PET to week 56 | Garetosmab/garetosmab | COVID-19 mITT | Bootstrap | -15.9%  (-34.5%, 3.5%) | -16.4%  (-39.8%, 8.8%) | ------------------- |  |
| Secondary – CT – Total Volume | Percent change from week 28 in the total volume of HO lesions as assessed by CT to week 56 | Placebo/garetosmab | COVID-19 mITT | Bootstrap + Wilcoxon | 34.0%  (1.7%, 91.9%) | 4.5%  (-1.7%, 11.7%) | -29.5%  (-91.2%, 7.0%) |  |
|  |  | Garetosmab/garetosmab | COVID-19 mITT | Bootstrap | 7.5%  (2.0%, 13.8%) | -3.5%  (-9.6%, 1.4%) | -11.1%  (-22.8%, -1.3%) |  |
| Secondary – CT – Total Volume | Percent change from baseline in the total volume of HO lesions as assessed by CT to week 56 | Garetosmab/garetosmab | COVID-19 mITT | Bootstrap | 7.5%  (2.0%, 13.8%) | 2.6%  (-2.0%, 7.1%) | ------------------- |  |
| Secondary – PET – Number of Lesions | Change from week 28 in number of HO lesions by 18F-NaF PET to week 56 | Placebo/garetosmab | COVID-19 mITT | Bootstrap + Wilcoxon | -1.0  (-2.0, 0.1) | -2.1  (-2.9, -1.4) | -1.1  (-3.0, 0.4) |  |
|  |  | Garetosmab/garetosmab | COVID-19 mITT | Bootstrap | -2.4  (-3.5, -1.4) | -1.4  (-2.1, -0.7) | 1.3  (-0.2, 2.8) |  |
| Secondary – PET – Number of Lesions | Change from baseline in number of HO lesions by 18F-NaF PET at week 56 | Garetosmab/garetosmab | COVID-19 mITT | Bootstrap | -2.4  (-3.5, -1.4) | -4.0  (-4.9, -3.2) | ------------------- |  |
| Secondary – CT – Number of Lesions | Change from week 28 in number of HO lesions by CT to week 56 | Placebo/garetosmab | COVID-19 mITT | Bootstrap + Wilcoxon | 1.1  (0.5, 2.0) | -0.2  (-0.5, 0.1) | -1.3  (-2.4, -0.5) |  |
|  |  | Garetosmab/garetosmab | COVID-19 mITT | Bootstrap | -0.4  (-1.1, 0.2) | -0.1  (-0.2, 0.0) | 0.3  (-0.2, 0.9) |  |
| Secondary – CT – Number of Lesions | Change from baseline in number of HO lesions by CT to week 56 | Garetosmab/garetosmab | COVID-19 mITT | Bootstrap | -0.4  (-1.1, 0.2) | -0.4  (-1.2, 0.2) | ------------------- |  |
| Secondary – NRS | Daily pain due to FOP as measured using the daily NRS | Placebo/garetosmab | COVID-19 mITT | Bootstrap + Wilcoxon | 1.79  (1.0, 2.7) | 1.60  (0.9, 2.4) | -0.19  (-0.6, 0.1) |  |
|  |  | Garetosmab/garetosmab | COVID-19 mITT | Bootstrap | 1.31  (0.7, 2.1) | 1.15  (0.5, 2.0) | -0.16  (-0.4, 0.1) |  |
| Secondary - Glucocorticoids | Total dosage of glucocorticoids use over time | Placebo/garetosmab | COVID-19 mITT | Bootstrap + Wilcoxon | 4186.4  (2198.6, 6451.8) | 1099.1  (411.6, 1925.0) | -3087.3  (-5514.1, -982.3) |  |
|  |  | Garetosmab/garetosmab | COVID-19 mITT | Bootstrap | 691.7  (222.2, 1260.3) | 493.9  (47.5, 1080.6) | -197.8  (-2230, 2000) |  |

CT, computerized tomography; FOP, fibrodysplasia ossificans progressiva; GEE, general estimating equation; HO, heterotopic ossification; LS, least-squares; mITT, modified intention to treat; MMRM, mixed model with repeated measures; NRS, numeric rating scale; PET, positron emission tomography; SUVmax, maximum standardized uptake value; TLA, total lesion activity.

#### Table S6. Key prespecified secondary, exploratory, and post-hoc analyses for Period_1*

| **Assessment**  **(Type)** | **Outcome** | **Garetosmab** | **Placebo** | **Reduction** | ***P*-Value¶** |
| --- | --- | --- | --- | --- | --- |
| New ^18^F-NaF PET lesions (post-hoc) | Total lesion activity in new lesions per patient by ^18^F-NaF PET at Week 28 (mean) | 5.22 g | 205.99 g | 97% relative reduction | 0.009 |
| New CT lesions  (post-hoc) | Total volume of new lesions per patient by CT at Week 28 (mean) | 1.06 cm^3^ | 10.21 cm^3^ | 90% relative reduction | 0.017 |
| ^18^F-NaF PET lesions (prespecified exploratory) | Percent of patients with new bone lesions measured by ^18^F-NaF PET over 28 weeks | 15.0% (3/20) | 45.8% (11/24) | 67.4% reduction in risk (relative risk=0.33) | 0.05 |
| New CT lesions (post-hoc) | Percent of patients with new bone lesions measured by CT over 28 weeks | 15.0% (3/20) | 45.8% (11/24) | 67.4% reduction in risk (relative risk=0.33) | 0.05 |
| New ^18^F-NaF PET lesions (post-hoc) | Number of new bone lesions measured by ^18^F-NaF PET over 28 weeks | Rate: 0.15 Count: 3 LA/pt: 34.8 g† | Rate: 1.2 Count: 29 LA/pt:  449.4 g† | 87.4% decrease (0.13 rate ratio) | 0.006 |
| New CT lesions (post-hoc) | Number of new bone lesions per patient measured by CT over 28 weeks† | Rate: 0.15 Count: 3 Vol/pt: 7.1 cm^3^† | Rate: 1.1 Count: 27 Vol/pt:  22.3 cm^3^† | 86.7% decrease (0.13 rate ratio) | 0.009 |
| ^18^F-NaF PET lesions (prespecified secondary) | Number of HO lesions  (95% CI) by ^18^F-NaF PET per scan over 28 weeks† | 3.5 (2.8, 4.3) | 4.8 (4.0, 5.7) | 27% decrease (0.73 rate ratio) | 0.03 |
| Imaging – SUVmax (prespecified secondary) | Percent change from baseline in SUVmax at week 8, LS mean (SE)# | -22.3 (4.0) | -6.6 (3.7) | -15.7 (-26.8, -4.6) | 0.007 |
| Imaging – SUVmax  (post-hoc) | Percent change from baseline in SUVmax at week 28, LS mean (SE)# | -34.0 (3.7) | -11.6 (3.4) | -22.4 (-32.6, -12.2) | <0.001 |
| Imaging – number of  CT lesions (prespecified secondary) | Number of HO lesion/scan (95% CI) through 28 weeks† | 5.3 (4.7, 6.1) | 6.3 (5.6, 7.0) | 16% decrease  (0.84 rate ratio) | 0.07 |

^18^F-NaF PET=fluorine-18-labelled sodium fluoride positron emission tomography; ANCOVA=analysis of covariance; CI=confidence interval; CT=computed tomography; HO=heterotopic ossification; LS=least squares; Q4W=every 4 weeks; SUVmax=maximal standardized uptake value; SUVmean=mean standardized uptake value; SE=standard error. *Active HO analysis set. †LA/pt, Vol/pt: lesion activity per patient and volume per patient are calculated by averaging new lesion activities/volume among patients who had new lesions. †Negative binomial model. #ANCOVA. ¶Nominal *P*-value.

#### Table S7. TEAEs that occurred in ≥10% of the garetosmab 10 mg/kg Q4W group in the double-blind period of the study (Period_1)*^,†^

| **Primary System Organ Class Preferred Term** | **Placebo (n=24)** | **Garetosmab 10 mg/kg Q4W**  **(n=20)** |
| --- | --- | --- |
| Number of TEAEs | 246 | 307 |
| Patients with ≥1 TEAE, n (%) | 24 (100) | 20 (100) |
| Skin and subcutaneous tissue disorders, n (%) | 11 (45.8) | 18 (90.0) |
| Acne | 3 (12.5) | 6 (30.0) |
| Madarosis | 0 | 6 (30.0) |
| Dry skin | 1 (4.2) | 3 (15.0) |
| Pruritus | 1 (4.2) | 3 (15.0) |
| Alopecia | 1 (4.2) | 2 (10.0) |
| Blister | 0 | 2 (10.0) |
| Eczema | 0 | 2 (10.0) |
| Rash | 4 (16.7) | 2 (10.0) |
| Rash papular | 0 | 2 (10.0) |
| Infections and infestations, n (%) | 10 (41.7) | 14 (70.0) |
| Rhinitis | 0 | 4 (20.0) |
| Folliculitis | 0 | 3 (15.0) |
| Gastroenteritis | 1 (4.2) | 3 (15.0) |
| Nasopharyngitis | 5 (20.8) | 3 (15.0) |
| Rash pustular | 0 | 2 (10.0) |
| Abscess limb | 0 | 2 (10.0) |
| Respiratory, thoracic, and mediastinal disorders, n (%) | 9 (37.5) | 14 (70.0) |
| Epistaxis | 4 (16.7) | 10 (50.0) |
| Cough | 2 (8.3) | 3 (15.0) |
| Gastrointestinal disorders, n (%) | 9 (37.5) | 13 (65.0) |
| Diarrhea | 6 (25.0) | 5 (25.0) |
| Toothache | 0 | 4 (20.0) |
| Nausea | 2 (8.3) | 3 (15.0) |
| Abdominal pain | 1 (4.2) | 2 (10.0) |
| Abdominal pain upper | 1 (4.2) | 2 (10.0) |
| Mouth ulceration | 1 (4.2) | 2 (10.0) |
| Musculoskeletal and connective tissue disorders, n (%) | 20 (83.3) | 13 (65.0) |
| Arthralgia | 7 (29.2) | 5 (25.0) |
| Pain in extremity | 9 (37.5) | 5 (25.0) |
| Back pain | 1 (4.2) | 4 (20.0) |
| Musculoskeletal pain | 2 (8.3) | 4 (20.0) |
| Neck pain | 3 (12.5) | 4 (20.0) |
| Joint swelling | 2 (8.3) | 3 (15.0) |
| Muscle spasms | 0 | 2 (10.0) |
| Myalgia | 3 (12.5) | 2 (10.0) |
| Nervous system disorders, n (%) | 11 (45.8) | 12 (60.0) |
| Headache | 7 (29.2) | 10 (50.0) |
| Dizziness | 2 (8.3) | 4 (20.0) |
| General disorders and administration site conditions, n (%) | 9 (37.5) | 11 (55.0) |
| Pyrexia | 0 | 3 (15.0) |
| Chest discomfort | 0 | 2 (10.0) |
| Fatigue | 2 (8.3) | 2 (10.0) |
| Infusion site pain | 0 | 2 (10.0) |
| Blood and lymphatic system disorders, n (%) | 1 (4.2) | 4 (20.0) |
| Anemia | 1 (4.2) | 2 (10.0) |

Q4W=every 4 weeks; TEAE=treatment-emergent adverse event. *Safety analysis set. †A patient with multiple TEAEs is counted once for the same preferred term or system organ class. This table is sorted by descending order of frequency of system organ class and preferred term for the treatment group.

#### Table S8. Skin and soft tissue infection TEAEs*^,†^

| **Period_1** | | |  |  |
| --- | --- | --- | --- | --- |
|  | | | **Placebo**  **(n=24)** | **Garetosmab 10 mg/kg Q4W**  **(n=20)** |
| **Skin and subcutaneous infections** | | | 3 (12.5%) | 12 (60%) |
|  | Acne | | 3 (12.5%) | 6 (30.0%) |
|  | Folliculitis | | 0 | 3 (15%) |
|  | Abscess limb | | 0 | 2 (10.0%) |
|  | Anal abscess | | 0 | 1 (5.0%) |
|  | Anorectal cellulitis | | 0 | 1 (5.0%) |
| Pustule | | | 0 | 1 (5.0%) |
| **Period_2 and open-label extension** | | | | |
|  | | **Total**  **(N=43)** | | |
| **Skin and subcutaneous infections** | | 29 (67.4%) | | |
|  | Acne | 16 (37.2%) | | |
|  | Abscess limb | 7 (16.3%) | | |
|  | Anal abscess | 5 (11.6%) | | |
|  | Folliculitis | 4 (9.3%) | | |
|  | Furuncle | 4 (9.3%) | | |
| Abscess | | 3 (7.0%) | | |
| Subcutaneous abscess | | 3 (7.0%) | | |
|  | Abdominal abscess | 2 (4.7%) | | |
|  | Groin abscess | 2 (4.7%) | | |
| Perineal abscess | | 2 (4.7%) | | |
| Tooth abscess | | 2 (4.7%) | | |
| Vulval abscess | | 2 (4.7%) | | |
| Abscess oral | | 1 (2.3%) | | |
| Abscess rupture | | 1 (2.3%) | | |
| Acne cystic | | 1 (2.3%) | | |
|  | Acne pustular | 1 (2.3%) | | |
|  | Carbuncle | 1 (2.3%) | | |
|  | Cellulitis | 1 (2.3%) | | |
| Cellulitis orbital | | 1 (2.3%) | | |
|  | Perirectal abscess | 1 (2.3%) | | |
|  | Pustule | 1 (2.3%) | | |
|  | Skin infection | 1 (2.3%) | | |

Q4W=every 4 weeks; TEAE=treatment-emergent adverse event. *Safety analysis set. †A patient with multiple TEAEs is counted once for the same preferred term or system organ class. This table is sorted by descending order of frequency of system organ class and preferred term for the treatment group.

#### Table S9. Serious adverse events that occurred in the double-blind period of the study (Period_1)*^,†^

| **Primary System Organ Class Preferred Term** | **Placebo (n=24)** | **Garetosmab 10 mg/kg Q4W (n=20)** |
| --- | --- | --- |
| Number of SAEs | 2 | 7 |
| Patients with ≥1 SAE, n (%) | 2 (8.3) | 4 (20.0) |
| Infections and infestations, n (%) | 1 (4.2) | 3 (15.0) |
| Gastroenteritis | 0 | 1 (5.0) |
| Pneumonia | 0 | 1 (5.0) |
| Sepsis | 0 | 1 (5.0) |
| Urinary tract infection | 0 | 1 (5.0) |
| Gastroenteritis viral | 1 (4.2) | 0 |
| Gastrointestinal disorders, n (%) | 0 | 1 (5.0) |
| Intestinal obstruction | 0 | 1 (5.0) |
| Respiratory, thoracic, and mediastinal disorders, n (%) | 0 | 1 (5.0) |
| Epistaxis | 0 | 1 (5.0) |
| Musculoskeletal and connective tissue disorders, n (%) | 1 (4.2) | 0 |
| Joint swelling | 1 (4.2) | 0 |

Q4W=every 4 weeks; SAE=serious adverse event. *Safety analysis set. †A patient with multiple SAEs is counted once for the same preferred term or system organ class. This table is sorted by descending order of frequency of system organ class and preferred term for the treatment group.

#### Table S10. Bleeding TEAEs

| **Period_1** | | | |  |  |
| --- | --- | --- | --- | --- | --- |
|  | | | | **Placebo**  **(n=24)** | **Garetosmab 10 mg/kg Q4W**  **(n=20)** |
| **Any Bleeding event** | | | | 9 (37.5) | 13 (65.0) |
|  | | Epistaxis | | 4 (16.7) | 10 (50.0) |
|  | | Petechiae | | 0 | 1 (5.0) |
|  | | Hemoptysis | | 0 | 1 (5.0) |
|  | | Increased tendency to bruise | | 0 | 1 (5.0) |
|  | | Hematuria | | 3 (12.5) | 0 |
|  | | Metrorrhagia | | 1 (4.2) | 0 |
|  | | Vaginal hemorrhage | | 1 (4.2) | 0 |
|  | | Hematoma | | 1 (4.2) | 0 |
|  | | Chronic pigmented purpura | | 1 (4.2) | 0 |
|  | | Menorrhagia | | 1 (4.2) | 0 |
| **Period_2 and open-label extension** | | | | | |
|  | | | **Total**  **(N=43)** | | |
| **Any Bleeding event** | | | 18 (41.9) | | |
|  | Epistaxis | | 15 (34.9) | | |
|  | Hematuria | | 2 (4.7) | | |
|  | Hematochezia | | 2 (4.7) | | |
|  | Ecchymosis | | 2 (4.7) | | |
|  | Petechiae | | 1 (2.3) | | |
|  | Nipple exudate bloody | | 1 (2.3) | | |
|  | Vaginal hemorrhage | | 1 (2.3) | | |
|  | Nail bed bleeding | | 1 (2.3) | | |

#### Table S11. TEAEs that occurred in ≥10% in Period_2 and the open-label extension period until end of study*^,†^

| **Primary System Organ Class Preferred Term** | **Total (N=43)** |
| --- | --- |
| Number of TEAEs | 865 |
| Patients with ≥1 TEAE, n (%) | 43 (100%) |
| Infections and infestations, n (%) | 37 (86.0) |
| Nasopharyngitis | 14 (32.6) |
| Abscess limb | 6 (14.0) |
| Rhinitis | 5 (11.6) |
| Anal abscess | 4 (9.3) |
| Folliculitis | 4 (9.3) |
| Furuncle | 4 (9.3) |
| Gastroenteritis | 4 (9.3) |
| Paronychia | 4 (9.3) |
| Abdominal abscess | 2 (4.7) |
| Groin abscess | 2 (4.7) |
| Hordeolum | 2 (4.7) |
| Subcutaneous abscess | 2 (4.7) |
| Vulval abscess | 2 (4.7) |
| Skin and subcutaneous tissue disorders, n (%) | 35 (81.4) |
| Madarosis | 20 (46.5) |
| Acne | 14 (32.6) |
| Alopecia | 8 (18.6) |
| Rash | 6 (14.0) |
| Hirsutism | 4 (9.3) |
| Erythema | 3 (7.0) |
| Hypertrichosis | 3 (7.0) |
| Decubitus ulcer | 2 (4.7) |
| Musculoskeletal and connective tissue disorders, n (%) | 25 (58.1) |
| Arthralgia | 15 (34.9) |
| Pain in extremity | 12 (27.9) |
| Back pain | 10 (23.3) |
| Musculoskeletal pain | 5 (11.6) |
| Neck pain | 5 (11.6) |
| Spinal pain | 5 (11.6) |
| Myalgia | 3 (7.0) |
| Muscular weakness | 2 (4.7) |
| Gastrointestinal disorders, n (%) | 24 (55.8) |
| Diarrhea | 7 (16.3) |
| Nausea | 7 (16.3) |
| Vomiting | 4 (9.3) |
| Aphthous ulcer | 3 (7.0) |
| Mouth ulceration | 3 (7.0) |
| Injury, poisoning and procedural complications, n (%) | 22 (51.2) |
| Post-traumatic pain | 6 (14.0) |
| Contusion | 5 (11.6) |
| Joint injury | 4 (9.3) |
| Skin laceration | 3 (7.0) |
| Respiratory, thoracic, and mediastinal disorders, n (%) | 22 (51.2) |
| Epistaxis | 15 (34.9) |
| Cough | 5 (11.6) |
| Oropharyngeal pain | 3 (7.0) |
| Rhinorrhea | 3 (7.0) |
| General disorders and administration site conditions, n (%) | 19 (44.2) |
| Pyrexia | 11 (25.6) |
| Pain | 3 (7.0) |
| Swelling | 2 (4.7) |
| Nervous system disorders, n (%) | 19 (44.2) |
| Headache | 13 (30.2) |
| Dizziness | 4 (9.3) |
| Reproductive system and breast disorders, n (%) | 9 (20.9) |
| Ovarian cyst | 3 (7.0) |
| Ear and labyrinth disorders, n (%) | 6 (14.0) |
| Hypoacusis | 3 (7.0) |
| Ear discomfort | 2 (4.7) |

Q4W=every 4 weeks; TEAE=treatment-emergent adverse event. *Safety analysis set. †A patient with multiple TEAEs is counted once for the same preferred term or system organ class. This table is sorted by descending order of frequency of system organ class and preferred term for the treatment group.

#### Table S12. Serious adverse events that occurred in the open-label period*^,†^

| **Primary System Organ Class Preferred Term** | **Total (N=43)** |
| --- | --- |
| Number of SAEs | 20 |
| Patients with ≥1 SAE, n (%) | 13 (30.2) |
| Infections and infestations, n (%) | 7 (16.3) |
| Abscess | 1 (2.3) |
| Abscess limb | 1 (2.3) |
| Perineal abscess | 1 (2.3) |
| Perirectal abscess | 1 (2.3) |
| Pneumonia | 1 (2.3) |
| Respiratory tract infection viral | 1 (2.3) |
| Subcutaneous abscess | 1 (2.3) |
| General disorders and administration site conditions, n (%) | 3 (7.0) |
| Pyrexia | 2 (4.7) |
| Sudden death | 1 (2.3) |
| Injury, poisoning and procedural complications, n (%) | 3 (7.0) |
| Head injury | 1 (2.3) |
| Skull fracture | 1 (2.3) |
| Splenic rupture | 1 (2.3) |
| Gastrointestinal disorders, n (%) | 2 (4.7) |
| Crohn's disease | 1 (2.3) |
| Intestinal obstruction | 1 (2.3) |
| Nervous system disorders, n (%) | 1 (2.3) |
| Cerebrovascular accident | 1 (2.3) |
| Renal and urinary disorders, n (%) | 1 (2.3) |
| Renal colic | 1 (2.3) |
| Respiratory, thoracic, and mediastinal disorders, n (%) | 1 (2.3) |
| Acute respiratory failure | 1 (2.3) |

Q4W=every 4 weeks; SAE=serious adverse event. *Safety analysis set. †A patient with multiple SAEs is counted once for the same preferred term or system organ class. The table is sorted by descending order of frequency of system organ class and preferred term for the total group. Included are SAEs that were reported on or before the end of study date of October 30, 2020.

#### Table S13. Case summaries of patients who died

| Case 1 | A 26 to 30-year-old with a CAJIS score of 16/30 with ankylosis of hips, knee, and shoulders, had a SAE of severe head injury (resulting from a fall) in the open-label extension, which was immediately followed by death. The patient had been randomized to garetosmab in Period 1 and received 16 doses of garetosmab by the time of the event. Prior to this event, the patient had severe gait impairment and required aid to perform daily activities. An autopsy showed major head trauma, with a laceration of the scalp in the vertex region and periorbital hematoma on an external examination. Internal examination revealed a diastatic skull fracture together with diffuse subdural and subarachnoid hemorrhages, as well as multiple fractured ribs.^1^  The cause of the death was reported as being due to severe head and brain trauma due to a fall, in the setting of a patient with diffuse rigidity and walking disability. |
| --- | --- |
| Case 2 | A 41 to 45-year-old with a CAJIS score of 28/30 commensurate with complete ankylosis in the spine, shoulders, hips, knee joints, and locked jaw, experienced a hemorrhagic stroke in the open-label extension. The patient had been randomized to garetosmab in Period 1 and received 16 doses of garetosmab by the time of the event. During hospitalization, the patient’s condition deteriorated, and the patient died. The cause of death was reported as a massive hemorrhagic stroke to the deep structures of the brain leading to acute respiratory failure. No autopsy was performed on patient’s family demand. This patient had poorly controlled arterial hypertension and had experienced 2 AEs of worsening hypertension during the study. Review of the head CT images (captured on the day of the hospitalization and a couple of days after the event), later performed by 2 independent radiologists reported findings suggestive of chronic hypertensive effects and agreed that the hemorrhage was typical for a hypertensive bleed and not due to a ruptured cerebral vascular malformation. |
| Case 3 | A 36 to 40-year-old patient with a CAJIS score of 30/30 with complete ankylosis of the spine, shoulder, hip, and knee joints bilaterally as well as significant jaw stiffness, experienced a fatal SAE of intestinal obstruction during Period_2. The patient had been randomized to garetosmab in Period 1 and received 14 doses of garetosmab by the time of the event. The patient was hospitalized for constipation, non-specific abdominal pain, vomiting, and fever. Imaging studies confirmed a mechanical obstruction and laboratory tests showed elevated inflammatory markers. They were treated with enemas, nasogastric tube, and broad-spectrum antibiotics. The patient’s condition initially improved, but rapidly deteriorated following discharge to a nursing home facility resulting in death. An autopsy was not performed. The patient had experienced an episode of intestinal obstruction during study Period_1 which also had led to a SAE (hospitalization) and resolved with clinical procedures, without the need of surgical intervention. |
| Case 4 | A 31 to 35-year-old patient with a CAJIS score of 26/30 with almost complete ankylosis of the neck, thoracic and lumbar spine, shoulders, elbows, hips, and knees and with movement only in wrists and ankles experienced a fatal SAE of traumatic splenic rupture following a fall in the open-label extension. The patient had been randomized to placebo (7 doses) in Period 1 and received 8 doses of garetosmab in the open-label period by the time of the event. The patient was "frozen" on a flat horizontal position, though with assistance they could switch to a vertical position, with limited balance. A surgical procedure was started, but after a difficult intubation the patient experienced cardiac arrest and died. An autopsy was not performed, and the investigator assessed this SAE as more likely related to the severity of the underlying disease and the trauma. |
| Case 5 | A 36 to 40-year-old patient with a CAJIS score of 19/30 experienced sudden death in the open-label extension. The patient had been randomized to placebo (7 doses) in Period 1 and received 15 doses of garetosmab in the open-label period by the time of the event. The patient's past medical history was significant for hypothyroidism, brain cavernoma hemorrhage, mild renal insufficiency, and an uncommon skin condition known as pigmented purpuric dermatosis. A preliminary macroscopic post-mortem exam noted gross right lung hemorrhage, but this was later determined to be incorrect. A full autopsy report ruled out any lung hemorrhage or acute bleeding event and determined that the most likely cause of death was aspiration pneumonia. After consultation with a second pathologist, the final cause of death was considered sudden cardiac death, with additional findings of extensive granulomatous inflammation most likely attributable to chronic on-going aspiration of foreign material. |

#### Table S14. Exploratory analysis of percent change of total volume (cm^3^) of target lesions by CT at week 8 and week 28 versus baseline

|  | **Placebo**  **(n=24)** | **Garetosmab 10 mg/kg Q4W (n=20)** |
| --- | --- | --- |
| **Baseline** |  |  |
| Number of patients included | 24 | 20 |
| Total number of target lesions | 134 | 119 |
| Total volume of target HO lesions (cm^3^) |  |  |
| n | 24 | 20 |
| Mean (SD) | 235.8 (253.3) | 251.4 (327.9) |
| Median | 159.7 | 112.3 |
| Q1 : Q3 | 113.6 : 267.3 | 46.5 : 242.9 |
| Min : Max | 17.3 : 1173.3 | 5.2 : 1046.6 |
| **Week 8** |  |  |
| Number of patients included | 24 | 20 |
| Total number of target lesions | 136 | 114 |
| Total volume of target HO lesions (cm^3^) |  |  |
| n | 24 | 20 |
| Mean (SD) | 245 (264.8) | 258.3 (330.3) |
| Median | 174.9 | 112 |
| Q1 : Q3 | 120.5 : 261.3 | 54.8 : 258.7 |
| Min : Max | 28:57.8 | 7.8 : 1076.8 |
| Percent change from baseline |  |  |
| n | 24 | 20 |
| Mean (SD) | -0.2% (18.0%) | 11.2% (18.1%) |
| Median | 1.4% | 5.6% |
| Q1 : Q3 | -2.8% : 8.1% | -0.3% : 12.4% |
| Min : Max | -55.1% : 37.7% | -7.8% : 59.1% |
| Median % difference*: Week 8 vs. baseline (95% CI) | -1.61% (6.37%, 3.79%) | -6.86% (-19.7%, -1.93%) |
| Nominal *P* value vs. baseline | 0.509 | 0.006 |
| Adjusted nominal *P* value vs. baseline | 0.509 | 0.024 |
| **Week 28** |  |  |
| Number of patients included | 24 | 20 |
| Total number of target lesions | 136 | 110 |
| Total volume of target HO lesions (cm^3^) |  |  |
| n | 24 | 20 |
| Mean (SD) | 242.2 (244.5) | 256.9 (332.5) |
| Median | 194.1 | 116.4 |
| Q1 : Q3 | 117.6 : 270.9 | 53.6 : 264.8 |
| Min : Max | 15.3 : 1147 | 6.9 : 1077.2 |
| Percent change from baseline |  |  |
| n | 24 | 20 |
| Mean (SD) | 11.1% (40.7%) | 6.4% (13.4%) |
| Median | 1.0% | 4.0% |
| Q1 : Q3 | -1.2% : 8.1% | -2.8% : 12.4% |
| Min : Max | -36.0% : 189.0% | -12.7% : 40.6% |
| Median % difference*: Week 28 vs. baseline (95% CI) | -2.65% (-8.44%, 0.59%) | -4.75% (-12.1%, 0.33%) |
| Nominal *P* value vs. baseline | 0.128 | 0.070 |
| Adjusted nominal *P* value vs. baseline | 0.256 | 0.210 |

*Median difference from baseline and 95% CI was derived from the Hodges-Lehmann estimate and Moses distribution-free CI, respectively. The *P* value was obtained from a paired Wilcoxon signed rank test to compare the percent change of total volume of target lesions at week 8 and week 28 vs baseline.

The adjusted *P* value accounts for multiple comparisons using the Holm method.

### Study Sites and Investigators

**Hôpital Cochin, Service de Rhumatologie, Paris, France:** Jacques Fechtenbaum, Karine Briot

**Hôpital Lariboisiere, Hospitalier Universitaire Nord, Paris, France:** Jeremy Ora

**Department of Pediatrics, Unit of Rare Diseases, IRCCS Istituto Giannina Gaslini, Genoa, Italy:** Gioacchino A. Rotulo, Marta Bertamino, Sara Signa, Marcello Mariani

**Amsterdam UMC, Vrije Universiteit, Amsterdam, The Netherlands:** Bernard Smilde, Coen Netelenbos, Sanne Treurniet, Silvia Storoni, Ruben D. De Ruiter

**Institute of Medical Sciences, Medical College of Rzeszów University, Rzeszów University, Rzeszów, Podkarpackie, Poland:** Arthur Mazur, Katarzyna Wiacek

**Department of Rheumatology, Hospital Universitario Ramón y Cajal, Madrid, Spain:** Jesus L. Martos, Monica Vazquez-Diaz, Sandra Garrote-Corral

**Centre for Metabolic Bone Disease Royal National Orthopaedic Hospital NHS Trust, London, UK:** Vera Choida, Jonathan Moses, Tiara Gill

**The Perelman School of Medicine - The University of Pennsylvania, Philadelphia, PA, US:** Staci Kallish, Thomas Guzzo

**Department of Medicine, Mayo Clinic, Rochester, MN, US:** Lance Mynderse, Matthew Drake, Michael Brown, Stephen Broski, Landon Trost, Timothy Curry, Joshua Savage, Thomas Stewart, Julia Lehman, Alexander Meves

**Vanderbilt University Medical Center, Program for Metabolic Bone Disorders Nashville, TN, US:** Daniel Tilden, Kevin Niswender, Laura Bryant

### Supplementary section: Methods

#### Inclusion and exclusion criteria

The study population consisted of male and female patients aged 18–60 years with a clinical diagnosis of FOP and a history of FOP disease activity within 1 year of screening, and documentation of *ACVR1* mutation (Table S1). In addition, patients had to be willing and able to attend and comply with study visits and to undergo PET and CT imaging procedures. Patients were excluded if they used bisphosphonate therapies within 1 year of screening, as these medications alter bone metabolism and would confound the primary efficacy analysis.

To address a potential risk of embryotoxicity or male reproductive organ toxicity, the protocol excluded pregnant or breastfeeding women, as well as women of child-bearing potential and men who were unwilling to practice highly effective contraception.

- To address a potential risk of epistaxis identified during the study in Period_1, the following exclusion criteria were added (protocol amendment): Patients on concomitant antiplatelet therapy (eg, clopidogrel), anti-coagulants (eg, warfarin, heparin, factor Xa inhibitor, or thrombin inhibitors) in the last 30 days or within 5 half-lives of the therapy, whichever was longer. Low-dose acetylsalicylic acid (aspirin) was acceptable.
- Patients with a history of severe, non-traumatic bleeding requiring transfusion or hospitalization for hemodynamic compromise
- Patients with a known pre-existing medical history of a bleeding diathesis (eg, hemophilia A, von Willebrand’s Factor deficiency, platelet count ≤20x109/L)
- Detailed inclusion and exclusion criteria are provided in the full **protocol** available online.

#### Trial oversight

LUMINA-1 (NCT03188666) was conducted at 11 sites in 8 countries. The full **protocol** is available online. Patient safety and welfare were monitored by an Independent Data Monitoring Committee. This study was conducted in accordance with the 2013 Declaration of Helsinki and the International Council for Harmonization guidelines for GCP. All patients provided written, informed consent.

The trial was approved by the following Institutional Review Boards: University Health Network 700 University Ave.10th Floor, Suite 1056, Toronto Ontario, M5G1Z5, Canada; Comité de Protection des Personnes (CPP) Ile-de-F, 78 rue du Général Leclerc, Le Kremlin Bicentre, Paris France, 94275; Comitato Etico Regione Liguria, IRCCS Ospedale Policlinico, San Martino, Largo Rosanna Benzi, 10, Genova Italy, 16132; Science committee AMS, Attn. Dr. R.T.de Jongh, VUmc, internal medicine, room 4A35, De Boelelaan 1117, 1081 HV Amsterdam, The Netherlands; METC VUmc BS7, Kamer H-443, Postbus 7057, Amsterdam Netherlands, 1007 MD; Komisja Bioetyczna

Uniwersytetu Rzeszowskiego ul. Warszawska 26A, 35-205, Rzeszow Poland; Comité de Ética de la Investigación con medicamentos del Hospital Universitario Ramón y Cajal. Ctra. Colmenar, km. 9,100, Madrid Spain, 28034; London - Central Research Ethics Committee, 3rd Floor, Barlow House, 4 Minshull Street, Manchester UK, M13DZ; University of Pennsylvania, Office of Regulatory, 3624 Market Street, Suite 301 S, Philadelphia PA, 19104, United States; Mayo Clinic Institutional Review Board, 200 First Street SW, Rochester Minnesota, 55905, United States; Vanderbilt University

1313 21st Ave., South, Suite 505, Nashville Tennessee, 37232, United States.

#### Trial design

The COVID-19 pandemic caused delays in dose administration, PET/CT scan collection, and PET tracer availability (N=4). To mitigate confounding effects on study outcomes, week 56 and 76 analyses were based on a COVID-19 modified intent-to-treat (mITT) analysis, defined as all patients with active HO at baseline who received treatment in Period_2, and for whom at least one post-week 28 scan was collected with the period between consecutive garetosmab doses being <9 weeks before the first post-week 28 scan. Details of the protocol amendments are included in **Table S1**.

#### Imaging rationale

Imaging by ^18^F-NaF PET identifies new bone formation and mineralization through the accumulation of ^18^F as it substitutes hydroxyl groups in newly formed hydroxyapatite.^2^ Food and Drug Administration-approved imaging by ^18^F-NaF PET has been widely used to detect and quantify changes in abnormal osteogenic activity in several bone pathologies such as Paget’s disease, ankylosing spondylitis, and osteoblastic bone metastases.^3^ In patients with FOP, ^18^F-NaF PET has been used to identify HO lesions with a high PET signal, which also showed growth by CT over a period ranging from 5–20 months, while lesions with a low PET signal, consistent with bone remodeling in the normotopic skeleton, showed no growth by CT over the same period.^4^ CT allows identification and differentiation of HO lesions from normal skeletal bone and quantification of the volume of heterotopic bone. Whole-body volumetric measurement of HO by CT is recommended as a clinical endpoint in FOP studies by the International Clinical Council on FOP.^.^^5^

In LUMINA-1, ^18^F-NaF PET/CT was used to identify pre-existing target lesions at baseline, identify new HO lesions, measure osteogenic activity of bone lesions, differentiate mineralizing lesions from mature HO, and measure volume changes in pre-existing target and new HO lesions. Baseline imaging with PET/CT was performed at most 7 days prior to study drug administration and at subsequent timepoints (Figure S2).

A prior study demonstrated the utility of PET in FOP patients to identify active HO lesions by demonstrating that over a period of 5–20 months only those lesions with high PET signal showed growth by CT, while lesions with low PET signal (equivalent to that of the normotopic skeleton) showed no growth by CT.^19^ Furthermore, at the time of LUMINA-1’s design, it was unknown whether a sufficient number of new HO lesions would arise over the 28-week interval (Period_1) to rely on them alone for assessing the efficacy of garetosmab. Therefore, it was hypothesized that PET presented the most sensitive imaging modality to quantify total change in HO activity, whereas CT would enable detection of changes in volume of any HO lesion during Period_1.

#### Imaging acquisition and read procedures

Whole-body PET/CT scans were acquired. PET/CT acquisition and reconstruction parameters within pre-specified ranges were defined for each patient at the baseline scan and kept constant throughout the study. All PET/CT scans were transferred to a contract research organization for centralized quality control and review by 2 independent readers and an adjudicator; all 3 were blinded to treatment assignment. We hypothesized that HO lesions that showed the highest uptake of ^18^F-NaF on a baseline PET image would be most likely to show rapid growth when untreated and would be inhibited from growing by garetosmab treatment. HO lesions showing high 18F-NaF uptake were defined as being active, with a SUVmax ≥3 times that of the SUVmean of a normotopic reference region in the supra-acetabular area of the pelvis). The *SUV_mean_* (mean standardized uptake value) is defined as the mean decay-corrected activity concentration (r) within a given region of interest of a PET image divided by the total injected radioactivity dose (A_0_) normalized by body mass (M_0_) (${SUV}_{mean}=\frac{r}{\frac{A_{0}}{M_{0}}}$). *SUV_max_* (maximal *SUV*) is the is the *SUV* value of the of the most intense voxel within a region of interest (e.g. an HO lesion) in a PET image. Metabolic volume of an HO lesion is the sum volumes of voxels with and SUV above a threshold defined as 40% of the SUVmax of that lesion. Readers were instructed to manually exclude normotopic bone or other regions they deemed not appropriate to include within the metabolic volume of an HO lesion.

From each patient’s baseline PET/CT images, readers selected up to 7 candidate active HO pre-existing target lesions. Subsequently, an adjudicator chose up to 7 lesions from this initial pool as the set of pre-existing target lesions to be followed and quantified by both readers, beginning with the lesion with the greatest SUVmax and continuing in a descending order of signal magnitude. Pre-existing target lesions thus identified were followed throughout the study and included in the analysis of inhibitory effect on HO formation by garetosmab versus placebo. Decrease in volumetric growth was measured by CT in active pre-existing target lesions.

Appearance of new lesions was assessed by readers post-baseline based on PET/CT scans. When new lesions were identified by both readers, the adjudicator registered the decision as to which reader was more correct. Readers were blinded to the adjudicator’s assessment of new lesions. New lesions developing post-baseline as identified by PET were required to be active. New lesions identified by CT alone, consistent with HO location and morphology, were required to have a density >200 HU with a volume ≥1 cm^3^.

Changes in volumetric growth were assessed by CT in active, pre-existing target lesions. Readers independently assessed PET maximal standardized uptake value (SUVmax), mean SUV (SUVmean), SUV peak (SUVpeak), and total metabolic volume (MV) of each HO lesion. Total Lesion Activity (TLA), a measure of patient-level overall burden of growing and actively mineralizing HO lesions, was calculated as the sum of the product of target and new HO lesion’s SUVmean and metabolic volume at each timepoint.

#### Sample size

The study populations included a baseline-active HO analysis set (AHO) of all randomized patients with ≥1 active HO lesion at baseline. Patients with active HO lesions (ie, lesions with active mineralization) were defined as those patients at baseline that had at least one HO lesion demonstrating uptake of ^18^F-NaF PET of at least three times that of normal reference bone (ie, supracetabular bone) as assessed by central review. All randomized patients had active disease at baseline, so the AHO and full analysis set are identical. The Statistical Analysis Plan (**Supplementary Appendix**) also specified a baseline-active HO classic *ACVR1^R206H^* mutation analysis set (AHOC); only 2 randomized patients had atypical *ACVR1* mutations. The safety analysis set included all randomized patients who received any study drug.

The study was powered to provide 80% power at a 2-sided 0.05 significance with 12 patients per group, allowing detection of an observed treatment difference in the order of 57%, 65%, and 40% reduction in TLA, total lesion volume, and PET maximum standardized uptake value (SUVmax), respectively, based on preclinical, animal studies.^6-8^ Testing of primary and key secondary efficacy outcomes followed a pre-specified hierarchical testing procedure to address multiplicity at an overall 2-sided alpha=0.05 significance. Other secondary outcomes were tested at the nominal 2-sided alpha=0.05 significance without multiplicity adjustment. Safety outcomes were analyzed using descriptive statistics.

**BMP-9 methods**

Total soluble BMP-9 concentrations in human serum were measured using an ELISA, which uses a mouse anti-human BMP-9 monoclonal antibody as the capture reagent and recombinant BMP-9 as the standard. Captured soluble BMP-9 is detected using a biotinylated goat anti-human BMP-9 polyclonal antibody followed by Streptavidin conjugated with horseradish peroxidase (Poly-HRP Streptavidin). The LLOQ of the assay is 31.3 pg/mL in neat human serum.

#### Statistical analyses

The study was powered to provide 80% power at a 2-sided 0.05 significance with 12 patients per group, allowing detection of an observed treatment difference in the order of 57%, 65%, and 40% reduction in TLA, total lesion volume, and PET maximum standardized uptake value (SUVmax), respectively, based on other bone diseases and modeling in FOP mice. Testing of primary and key secondary efficacy outcomes followed a pre-specified hierarchical testing procedure to address multiplicity at an overall 2-sided alpha=0.05 significance for Period_1. Other secondary outcomes were tested at the nominal 2-sided alpha=0.05 significance without multiplicity adjustment. In Period_2, a 2-sided 10% significant level was applied for the primary and key secondary endpoints. Safety outcomes were analyzed using descriptive statistics.

TWA percent change in HO TLA from baseline was analyzed using an analysis of covariance (ANCOVA) model including treatment, gender, and baseline TLA as covariates. Percent change from baseline HO volume was analyzed using a mixed-effect model for repeated measures containing treatment, gender, visit, baseline total volume, and treatment-by-visit interaction as fixed effects.

The COVID-19 pandemic caused delays in dose administration, PET/CT scan collection, and PET tracer availability for a few patients (N=4). To mitigate confounding effects on study outcomes, Period_2 analyses were based on a COVID-19 modified intent-to-treat (mITT) analysis, defined as all patients with active HO at baseline who received treatment in Periods_2, and for whom at least one post-week 28 scan was collected with the period between consecutive garetosmab doses being <9 weeks before the first post-week 28 scan. Statistical methodology amendments are detailed in **Table S1**.

### Supplementary section: Results

Efficacy in period_1**:** Impact of garetosmab on pre-existing target lesions

Overall, 257 (PET) and 253 (CT) pre-existing target lesions were identified at baseline. Garetosmab reduced SUVmax of ^18^F-NaF in pre-existing target lesions by 22.6% (week 8) and 33.2% (week 28), versus reductions of 6.4% and 20.2%, respectively, with placebo (nominal *P*=0.005 and *P*=0.02, respectively; Figure S9). Garetosmab had no effect on the time-weighted average percent change from baseline versus placebo for TLA and total HO volume of pre-existing target lesions (TLA: least squares [LS] mean difference=0.4; 95% CI: -14.3–15.2; *P*=0.95; HO volume: LS mean difference=-4.8; 95% CI: -23.6–14.1; *P*=0.61; Figure S10). Nonetheless, garetosmab treatment resulted in resolution of 9 pre-existing target lesions by week 28 by CT; 1 pre-existing target lesion was resolved with placebo.

Efficacy in period_2**:** Patients remaining on garetosmab

Efficacy was maintained in Period_2 relative to Period_1 (0 new lesions by CT and PET versus 2 by CT and 1 by PET, respectively; Figure 2). Although the 2 lesions identified by CT in Period_1 were still detectable in Period_2, the volume remained stable (5.19 cm^3^ to 5.00 cm^3^) or substantially decreased (13.69 cm^3^ to 2.37 cm^3^). The percentage of patients with new lesions was 0% for Period_2 relative to Period_1 by PET and CT versus 5.6% by PET and 11.1% by CT for Period_1 relative to baseline (Figure 3). New lesion volume was 0 cm^3^ for Period_2 versus 1 cm^3^ for Period_1, and TLA was 0 versus 4.7, respectively (Figure 5). The proportion of patients with new flare-ups by patient diary was 22.2% in Period_2 versus 33.3% in Period_1, and by investigator was 5.6% versus 11.1%, respectively (Figure 4).
